# Associations of dementia polyexposure scores to Alzheimer’s disease endophenotypes in diverse populations

**DOI:** 10.64898/2026.01.10.26343864

**Authors:** Meri Okorie, Xiaqing Jiang, Kristine Yaffe, Jennifer S. Yokoyama, Shea J. Andrews, the Health and Aging Brain Study – Health Disparities

## Abstract

**INTRODUCTION:** Dementia clinical risk scores (CRS) provide accessible tools for identifying individuals at risk for Alzheimer’s disease (AD), yet their performance across diverse populations and relationships to AD endophenotypes remains unclear.

**METHODS:** We evaluated four CRS, mCAIDE, WHICAP, LIBRA, and CogDRisk, in relation to cognitive impairment diagnoses and endophenotypes for AD, including plasma biomarkers, neuroimaging measures, and cognitive composite scores. Logistic and linear regression models stratified by self-reported race/ethnicity were used to assess the associations of CRS with diagnosis and their predictive performance, and associations with endophenotypes.

**RESULTS:** Higher CRS were consistently associated with increased odds of dementia across all races/ethnicities. CogDRisk showed the strongest and most consistent performance across diagnostic and endophenotypic outcomes. The other three CRS performed similarly, with mCAIDE performing the worst and lacking associations with plasma biomarkers.

**CONCLUSIONS:** CRS capture AD-related risk across diverse populations and modestly reflect underlying biological endophenotypes, supporting their utility in community-based risk assessment.

## 1. BACKGROUND

Alzheimer’s disease (AD) presents substantial clinical and economic challenges, yet early identification of individuals at risk remains limited in diverse, community-based settings. Current AD risk and diagnostic assessment involve genetic and cognitive screening, cerebrospinal fluid (CSF) and plasma biomarkers of AD pathology, and neuroimaging; however, these approaches, though valuable, remain costly, invasive, and limited to those with access to specialized care in developed countries^1–3^. Cognitive testing and CSF and plasma biomarkers also only capture risks once pathology has begun, even if clinical symptoms of AD have not yet manifested^4–6^. In addition, many older adults, especially those from underserved populations, are evaluated in primary care contexts where advanced diagnostic resources are unavailable^7–9^. Given these limitations and the growing prevalence and burden of AD, there is a pressing need for effective risk assessment tools that can identify at-risk individuals earlier and support timely preventive interventions. The availability of such tools may also raise awareness of modifiable risk factors related to AD, supporting clinical actionability and potentially quicker diagnosis.

Clinical risk scores (CRS) have been developed as accessible tools that estimate AD and dementia risks based on demographics, lifestyle, and clinical profiles^10^. Many CRS can be readily implemented in community and primary care settings, often using self-reported information, and can help identify individuals who may benefit from further confirmatory testing with biomarkers or genetic screening^11,12^. CRS are designed to capture modifiable midlife-to-late-life risk factors, enabling individualized prevention strategies and risk stratification. These tools are particularly relevant for individuals with a family history of dementia or known genetic susceptibility, as they provide actionable targets for risk reduction and support precision health approaches. Many CRS have been proposed^13^ and validated in mid- and late-life populations across diverse settings, including the modified Cardiovascular Risk Factors, Aging, and Incidence of Dementia (mCAIDE) score^14^, the Washington Heights–Inwood Columbia Aging Project (WHICAP) score^15^, the Lifestyle for Brain Health (LIBRA) index^16^, and the Cognitive Dementia Risk (CogDRisk) score^17^. Among them, mCAIDE and WHICAP were independently developed using U.S.-based cohorts, while LIBRA and CogDRisk were developed based on systematic reviews and meta-analyses. Each integrates slightly different risk domains such as vascular health, lifestyle, psychosocial, and cognitive engagement factors, reflecting distinct methodological approaches and population contexts in which they were developed^14–20^.

External validation studies of CRS have reported variable performance across cohorts and settings. LIBRA and CogDRisk have been validated in multiple independent studies, with area under the curve (AUC) values ranging from approximately 0.53 to 0.77^21–26^. However, most validation studies were conducted in populations of predominantly European descent, limiting their generalizability to other racial and ethnic groups. The WHICAP score, which uniquely incorporates ethnicity-specific weighting, has been evaluated in racially and ethnically diverse urban cohorts, though small subgroup sample sizes have constrained statistical power^27^. The mCAIDE score has been compared with the original CAIDE index in large, multiethnic samples^28^, but few studies have independently validated mCAIDE itself.

Despite their potential, CRS have not been adequately validated across different racial and ethnic groups within the US population, raising questions about their generalizability and accuracy in diverse populations. Furthermore, most CRS were developed to aid in risk prediction of dementia diagnosis rather than the underlying disease pathology, and none of the validation studies have examined associations between these four CRS and AD biomarkers. Because abnormalities in AD biomarkers precede clinical syndromes of dementia^29–31^, the growing emphasis on AD pathophysiology highlights the need to validate CRS in detecting early abnormalities in biological endophenotypes. The present study examines how four widely used CRS relate to dementia diagnosis and to AD endophenotypes within a multiethnic cohort, the Health and Aging Brain Study - Health Disparities (HABS-HD). By investigating the utility of CRS across racial/ethnic groups and their sensitivity to early changes in AD endophenotypes, we aim to clarify the extent to which CRS can serve as equitable and biologically informative tools for AD and dementia risk assessment in real-world, community-based settings.

## 2. METHODS

### 2.1 Health & Aging Brain Study - Health Disparities

#### 2.1.1 Study Population

This study uses cross-sectional data from the HABS-HD cohort. HABS-HD is an ongoing community-based AD study that comprises participants from self-reported Black (AA), Latinx (LA) (Mexican American, Puerto Rican, Dominican Republic, etc.), and non-Hispanic White (NHW) populations recruited at the University of North Texas Health Science Center, Fort Worth, Texas, USA. The study collects data on demographics, AD biomarkers, neuroimaging, clinical history, and genomics, providing a comprehensive resource for AD and related dementia research. The study recruited generally healthy individuals free of serious mental illnesses and medical conditions, and we further excluded participants younger than 55 years old and those without *APOE* genotype information. The visit 1 baseline measurements for all outcomes of interest were used for all downstream analyses. All HABS-HD participants (and/or their legal guardians) signed written informed consent to participate in the study.

#### 2.1.2 Endophenotype Standardization

*Plasma biomarkers*: Levels of plasma biomarkers were quantified using commercially available kits, Quanterix, for all the participants of the HABS-HD. Plasma biomarkers (i.e., Aβ_42_, Aβ_40_, pTau_181_, Tau, NfL) were log-transformed to reduce the skewness, followed by z-score normalization (mean = 0, standard deviation (SD) = 1) for all participants to standardize and remove any batch effects. Aβ_42_/Aβ_40_ ratio was calculated by dividing the raw values of Aβ_42_ and Aβ_40_ prior to transformation and normalization^32^. We excluded participants with plasma biomarkers identified as outliers using the interquartile range methodology^33^. *Brain morphometry*: Cortical thickness was defined as the surface area-weighted average of cortical thickness in the right and left entorhinal cortex, fusiform, and inferior and middle temporal cortex. These regions of interest correspond to brain areas that are especially vulnerable to AD–related neurodegeneration. Hippocampal volume was determined by taking the mean volume of the right and left hippocampal volumes determined on a T1-weighted volume scan^34^. *Cognition*: The memory domain was assessed using immediate and delayed recall from the Wechsler Memory Scale-III (WMS-III) Logical Memory^35^ and the Spanish-English Verbal Learning Test (SEVLT)^36^. The verbal ability domain was assessed using Letter Fluency and Animal Naming tests^37^. The executive function domain was assessed using the WMS-III Digit Span^35^ and the Trail Making Test, Parts A and B^37^. All cognitive test scores were standardized using z-score transformation (mean = 0, SD = 1), and these scores for each domain (memory, verbal ability, and executive function) were averaged to create composite scores. In addition, the Mini-Mental State Examination (MMSE)^38^ and Clinical Dementia Rating (CDR) scale^39^ were administered to all participants as part of the neuropsychological assessment.

### 2.2 Modifiable Risk Factors

Education: Years of education were coded as a numeric variable, with values ranging from 0 (kindergarten) to the maximum value of 20 (post-graduate).

Body Mass Index (BMI): BMI at baseline was calculated as weight divided by height squared, with units converted to the standard metric of kg/m². Obesity status of participants was classified based on their BMI (kg/m²) as underweight (<18.5 kg/m²), normal weight (18.5–24.9 kg/m²), overweight (25–29.9 kg/m²), or obese (≥30 kg/m²).

Hypertension: All participants underwent a series of anthropometric measurements to assess blood pressure. Hypertension was defined as a past medical history of the condition or consistently elevated blood pressure, with at least two measurements of systolic blood pressure ≥140 mmHg or diastolic blood pressure ≥90 mmHg at the baseline visit.

Dyslipidemia/High Cholesterol: Hyperlipidemia was defined as meeting any of the following criteria: LDL cholesterol ≥120 mg/dL, total cholesterol ≥240 mg/dL, triglycerides ≥200 mg/dL, or a documented history of high cholesterol.

Diabetes: All participants had measures of hemoglobin A1C from fasting blood samples. Diabetes was defined as an A1C level ≥6.5% at the baseline visit or a documented history of the condition.

Depression: Participants were considered to have depression if they had a past medical history of the condition or scored ≥10 on the 30-item Geriatric Depression Scale (GDS).

Stroke: Participants self-reported (i.e., yes or no) a history of stroke or indicated if they had ever been told by a healthcare professional that they had experienced a stroke.

Traumatic Brain Injury (TBI): TBI with or without loss of consciousness information was collected through self-report data.

Renal Dysfunction: Estimated glomerular filtration rate (eGFR) was calculated using the 2021 Chronic Kidney Disease Epidemiology (CKD-EPI) creatinine equation, which estimates kidney function based on serum creatinine, age, and sex. eGFR values are reported in mL/min/1.73 m², and participants with eGFR <60 mL/min/1.73 m² were classified as having renal dysfunction, consistent with standard clinical definitions of chronic kidney disease.

Alcohol Intake: Alcohol intake was assessed using a single item from the Alcohol Use Disorders Identification Test (AUDIT): ‘How often do you have a drink containing alcohol?’ Response options included never, monthly or less, 2–4 times per month, 2–3 times per week, and 4 or more times per week.

Smoking: Smoking was classified as self-reported current smokers, indicating yes or no. Social Support: Loneliness was approximated using the social support questionnaires in the HABS-HD. The questionnaire consisted of 12 items assessing perceived availability of emotional, neighbourhood-level, and instrumental support.

Physical Activity: Physical activity was assessed using the Rapid Assessment of Physical Activity (RAPA), a questionnaire designed to provide a simple and accessible measure of physical activity among adults aged 50 years and older^40^. RAPA scores were derived according to established guidelines, with higher scores indicating greater levels of physical activity.

### 2.3 Clinical risk scores

For a detailed description of variables for each CRS, refer to Table S1.

#### 2.3.1 Modified Cardiovascular Risk Factors, Aging, and Dementia (mCAIDE)

mCAIDE was derived from the CAIDE score developed in Finland, but modified to better reflect U.S. cohorts and mid-to-late-life risk using two U.S.-based cohorts^14^. mCAIDE incorporates age, sex, education, BMI, systolic blood pressure (SBP), self-reported high cholesterol status, and physical activity measured by the mini Physical Performance Testing (miniPPT). The RAPA was used in place of mPPT to replace the physical activity measure. Participants whose total scores were in the lowest tertile were classified as ‘inactive,’ and those above this threshold as ‘active.’

#### 2.3.2 Washington Heights-Inwood Columbia Aging Project (WHICAP)

WHICAP is a CRS that was developed in an independent cohort of the Washington Heights and Inwood Community Aging project in Manhattan, NY, to create a risk index for late-onset AD^15^. It includes the variables sex, age, education, race/ethnicity, *APOE*\*ɛ4 carrier status, BMI, low high-density lipoprotein cholesterol (HDL-C), diabetes, hypertension, and current smoker. Among the four CRS considered in our analysis, WHICAP was the only score that explicitly incorporates race/ethnicity as a predictor. Therefore, we selected WHICAP to assess its transportability within HABS-HD, a multiethnic cohort from a different geographic region of the US. HDL-C was categorized as low or normal based on sex-specific cutoffs: males with HDL-C <50 mg/dL and females with HDL-C <40 mg/dL were classified as low, while values above these thresholds were classified as normal. *APOE\**ɛ4 carrier status was excluded from the scoring to prevent inflation of WHICAP relative to other CRS that do not incorporate *APOE\**ɛ4 information. Additionally, since genetic information is not always readily available in primary clinical settings, CRS that do not incorporate genetic factors may be more practical and more readily adopted.

#### 2.3.3. Lifestyle for Brain Health (LIBRA)

The LIBRA score was developed using an approach that combined a systematic review, meta-analysis, and Delphi consensus studies to identify and weight modifiable lifestyle, cardiovascular, and metabolic risk factors for all-cause dementia^16,19,41^. LIBRA was specifically designed to support primary prevention by focusing purely on modifiable risk factors in mid-to-late life and does not include age, sex, or education. The LIBRA score includes a healthy diet (i.e. Mediterranean diet), physical inactivity, cognitive activity, low to moderate alcohol consumption, current smoker, heart diseases, diabetes, high cholesterol, BMI, hypertension, renal dysfunction, and depression. Participants who reported drinking 2–3 times per week on the AUDIT questionnaire were classified as light to moderate consumers, corresponding to >0 and <14 units of alcohol per week. For the physical activity measure, participants who answered ‘yes’ to item 1 or item 2 on the RAPA questionnaire were classified as physically inactive. Healthy diet and cognitive activity measures were omitted from the scoring due to the lack of data in the HABS-HD study.

#### 2.3.4. Cognitive Health and Dementia Risk Reduction (CogDRisk)

CogDRisk was developed through a systematic review and meta-analysis of studies identified from international databases to quantify the impact of modifiable factors on cognitive decline and dementia^17,20^. The CogDRisk score includes age stratified by sex, education, midlife (≤65 yrs) obesity, dyslipidemia, diabetes stratified by sex, history of stroke, history of TBI, hypertension, atrial fibrillation, clinical diagnosis of insomnia, depression, physical inactivity, cognitive engagement, social engagement, diet, and current smoker. Participants who scored at least 1 SD below the mean on the social support questionnaire score were classified as ‘lonely,’ and those above this threshold were classified as ‘not lonely’ for the CogDrisk scoring. For physical activity measures, participants who answered ‘yes’ to item 6 or item 7 on the RAPA questionnaire were classified as physically active, reflecting an activity level of >150 minutes per week of moderate-to-vigorous activity. Baseline BMI at visit 1 was used in place of midlife obesity/BMI, which was not available in this cross-sectional study. Insomnia, atrial fibrillation, cognitive engagement, and diet measures were excluded from the scoring due to unavailability in the HABS-HD study.

### 2.3 Missingness

Missing data for variables used in the construction of the CRS were imputed using the *missForest* algorithm implemented in R^42^ for both continuous and categorical variables. The imputation dataset included relevant demographics, clinical variables, biomarkers, and predictors. Variables with high substantial missingness (> 40%), such as myocardial and heart disease, were excluded from imputation. To verify that imputation did not introduce bias, all analyses were compared against analyses using complete case data.

### 2.3 Mild Cognitive Impairment and Dementia Diagnosis

Cognitive status was assessed using self- and informant-reports, the Clinical Dementia Rating (CDR scale), and a neuropsychological battery covering memory, executive function, and verbal ability (animal naming, FAS, SEVLT, WMS-III Logical Memory, Digit Span, Trail Making, and Digit Symbol Substitution) as previously described^43,44^. Participants without any complaints of cognitive change, either reported by the individual or their informants, with a CDR sum of boxes (CDR-SOB) of 0, were classified as cognitively unimpaired. If a participant has an isolated poor neuropsychological test performance, in the absence of any cognitive complaints and functional decline, they were assigned as cognitively unimpaired. Mild cognitive impairment (MCI) was classified as participants with complaints of cognitive change, either reported by the individual or their informants, with a CDR-SOB score of 0.5-2.0 and performance at or below 1.5 SD below z-score adjusted norms on at least one cognitive test. Dementia diagnosis was made based on CDR sum of boxes score >=2.5, and cognitive test score at or below 2 SD below the mean on tests in two or more domains.

### 2.4 Statistical analysis

#### 2.4.1 Cognitive Impairment

Logistic regression models were used to assess the association of CRS with (i) MCI, (ii) dementia, and (iii) MCI + dementia, using cognitively normal participants as the reference group. Age, sex, *APOE* genotypes, and years of education were adjusted for in LIBRA, and *APOE* genotype was included as a covariate for mCAIDE, WHICAP, and CogDRisk. The *APOE* status was encoded as a categorical variable, categorized as *APOE*\*ε2+ carriers (ε2/ε2, ε2/ε3) and ε4+ carriers (ε2/ε4, ε3/ε4, or ε4/ε4 ), with the ε3/ε3 allele serving as the reference group. Model performance was evaluated by calculating the AUC values to assess model discrimination, and the Nagelkerke R-squared (R^2^) was calculated to determine the goodness-of-fit of the models. Self-reported race/ethnicity was used for stratified analysis. P-values were adjusted for multiple hypothesis testing by applying the false discovery rate (FDR) at 5%.

#### 2.4.2 AD Endophenotypes

Linear regression models were used to estimate the effects of CRS on AD endophenotypes, including (i) plasma biomarkers (Aβ_42_/Aβ_40_ ratio, pTau_181_, total Tau, and NfL), (ii) neuroimaging measures (cortical thickness and hippocampal volume), and (iii) clinical function (cognitive severity assessed by MMSE and clinical severity assessed by CDR, and memory, verbal ability, and executive function cognitive domains assessed by composite scores). For associations with plasma biomarkers, models for mCAIDE and WHICAP were adjusted for *APOE* genotype and eGFR, models for LIBRA were adjusted for age, sex, BMI, and *APOE* genotype, and models for CogDRisk were adjusted for *APOE* genotype, eGFR, and BMI. For neuroimaging associations, models for mCAIDE, WHICAP, and CogDRisk were adjusted for *APOE* genotype and intracranial volume. In contrast, models for LIBRA were adjusted for age, sex, *APOE* genotype, and intracranial volume. For cognitive function associations, models for mCAIDE, WHICAP, and CogDRisk were adjusted for interview language and *APOE* genotype, while models for LIBRA were adjusted for age, sex, *APOE* genotype, and interview language. P-values were adjusted for multiple hypothesis testing by applying the false discovery rate (FDR) at 5%.

For both logistic and linear regression analyses, we specified two baseline models: (i) a demographics model including age, sex, and years of education, and (ii) a demographics plus *APOE* genotype model. These baseline models were used to evaluate how much each CRS improved prediction beyond demographics alone and beyond demographics combined with *APOE* genotype.

#### 2.4.3 Racial/Ethnic stratified analysis

To evaluate whether the associations between each CRS and outcomes differed across racial/ethnic groups, we conducted pairwise z-tests comparing regression coefficients between subpopulations. For each score, the difference between standardized coefficients (β_1_–β_2_) was divided by the square root of the sum of their squared standard errors to calculate a z-statistic and two-sided p-value. Comparisons were performed for all pairwise combinations of racial/ethnic groups, excluding the overall population, with significant differences highlighted at an FDR-adjusted p < 0.05.

#### 2.4.4 Sensitivity analysis

Given that some demographic components of the CRS (e.g., age, sex, and years of education) are non-modifiable and less clinically actionable, we excluded these variables from mCAIDE, WHICAP, and CogDRisk. We then conducted regression analyses to assess their associations with endophenotypes and to evaluate the predictive accuracy of the models. For all linear and logistic regression models, age, sex, and years of education were included as covariates. P-values were adjusted for multiple hypothesis testing by applying the FDR at 5%.

## 3. RESULTS

### 3.1 Participant characteristics

Of the 2,627 participants, 1,907 were cognitively normal, and 720 were diagnosed with MCI or dementia (Table 1). Female participants (61.4%) comprised the majority in HABS-HD. Non-Hispanic White (NHW) individuals (N = 1,062, 40.4%) were the largest racial/ethnic group, followed by Latinx (LA) (N = 948, 36.1%) and then Black/African American (AA) participants (N = 617, 23.5%). Mean CRS values ranged from 0 to 0.1 in cognitively normal and MCI participants, increasing to 0.3 to 0.5 in those with dementia (Table 1).

**Table 1.** Characteristics of the HABS-HD cohort, including descriptive statistics of demographics, clinical risk scores (CRS), and AD endophenotypes. .

| Characteristics<sup>a</sup> | Cognitively Normal | Mild Cognitive Impairment | Dementia |
| --- | --- | --- | --- |
| Sample Size | 1907 | 540 | 180 |
| **Demographics** |  |  |  |
| Age | 66.7 (± 7.3) | 66.9 (± 8.1) | 70.3 (± 9.0) |
| Female | 1232 (64.6%) | 287 (53.1%) | 93 (51.7%) |
| NHW<sup>b</sup> | 865 (45.4%) | 139 (25.7%) | 58 (32.2%) |
| LA<sup>b</sup> | 680 (35.7%) | 195 (36.1%) | 73 (40.6%) |
| AA<sup>b</sup> | 362 (19.0%) | 206 (38.1%) | 49 (27.2%) |
| APOE-ε4 Carrier | 420 (25.9%) | 133 (30.5%) | 44 (29.7%) |
| **Clinical Risk Score** |  |  |  |
| mCAIDE | -0.070 (± 1.01) | 0.137 (± 0.97) | 0.335 (± 0.90) |
| WHICAP | -0.090 (± 0.97) | 0.158 (± 0.93) | 0.476 (± 1.30) |
| LIBRA | -0.088 (± 0.98) | 0.194 (± 1.00) | 0.354 (± 1.05) |
| CogDRisk | -0.067 (± 0.96) | 0.051 (± 1.03) | 0.555 (± 1.19) |
| **Cognition** |  |  |  |
| MMSE | 28.17 (± 2.15) | 26.60 (± 2.89) | 21.38 (± 5.64) |
| CDR | 0.0000 (± 0.00) | 1.2417 (± 0.86) | 4.5917 (± 2.93) |
| Executive Function | 0.26 (± 0.64) | -0.52 (± 0.77) | -1.21 (± 0.91) |
| Verbal Ability | 0.23 (± 0.75) | -0.40 (± 0.74) | -1.27 (± 0.74) |
| Memory | 0.32 (± 0.66) | -0.62 (± 0.67) | -1.56 (± 0.65) |
| **Neuroimaging** |  |  |  |
| Cortical Thickness | 2.75 (± 0.13) | 2.73 (± 0.14) | 2.58 (± 0.22) |
| Hippocampal Volume | 0.12 (± 0.92) | -0.14 (± 0.98) | -0.95 (± 1.35) |
| **Plasma Biomarkers** |  |  |  |
| Aβ<sub>42</sub>/Aβ<sub>40</sub> | 0.011 (± 0.99) | 0.053 (± 1.04) | -0.273 (± 0.90) |
| Total Tau | -0.023 (± 0.97) | 0.009 (± 1.03) | 0.243 (± 1.20) |
| pTau<sub>181</sub> | -0.10 (± 0.90) | 0.10 (± 1.09) | 0.77 (± 1.33) |
| NfL | -0.078 (± 0.92) | 0.061 (± 1.11) | 0.656 (± 1.26) |
| <sup>a</sup>Mean (SD), n (%), <sup>b</sup>Percentages calculated using the total cohort as the denominator for a given diagnosis |  |  |  |

Across all cognitive statuses, LA participants generally exhibited higher CRS compared to AA and NHW participants, particularly for mCAIDE and WHICAP, two cardiometabolic-centred CRS. The racial difference in WHICAP was most pronounced for dementia, with NHW participants having a mean of −0.40, AA participants with a mean of 0.1, and LA participants with a mean of 1.20. For LIBRA, mean scores were similar between LA and AA participants for MCI (0.30 and 0.20, respectively) and for dementia (0.51 and 0.50, respectively), while NHW participants had a mean of means of 0 for dementia and MCI diagnosis. For CogDRisk, LA and NHW participants had higher mean scores for MCI (0.19 and 0.33, respectively) compared to AA group with a mean of -0.27. For dementia diagnosis, LA and NHW participants had means of 0.63 and 0.65, respectively, compared to the mean of 0.33 for AA participants (Table 1).

### 3.2 Higher clinical risk burdens are associated with increased odds of cognitive impairment across racial/ethnic groups

Across all CRS, higher scores were consistently associated with greater odds of developing MCI and dementia in the total cohort. For MCI diagnosis, each SD increase in CRS corresponded to a 21-32% higher risk, with the strongest associations observed for LIBRA (32%), followed by

WHICAP (30%) and mCAIDE (21%). In contrast, CogDRisk was not significantly associated with MCI risk. Associations were stronger for dementia risk, with each SD increase in CRS corresponding to a 35–73% increase in odds. WHICAP demonstrated the largest effect (73%), followed by CogDRisk (68%) and LIBRA (61%). When MCI and dementia were combined, effect sizes were attenuated across all CRS relative to the dementia-only group, due to a weaker association of CRS with MCI (Fig. 1, Table S2).

**Figure 1.**
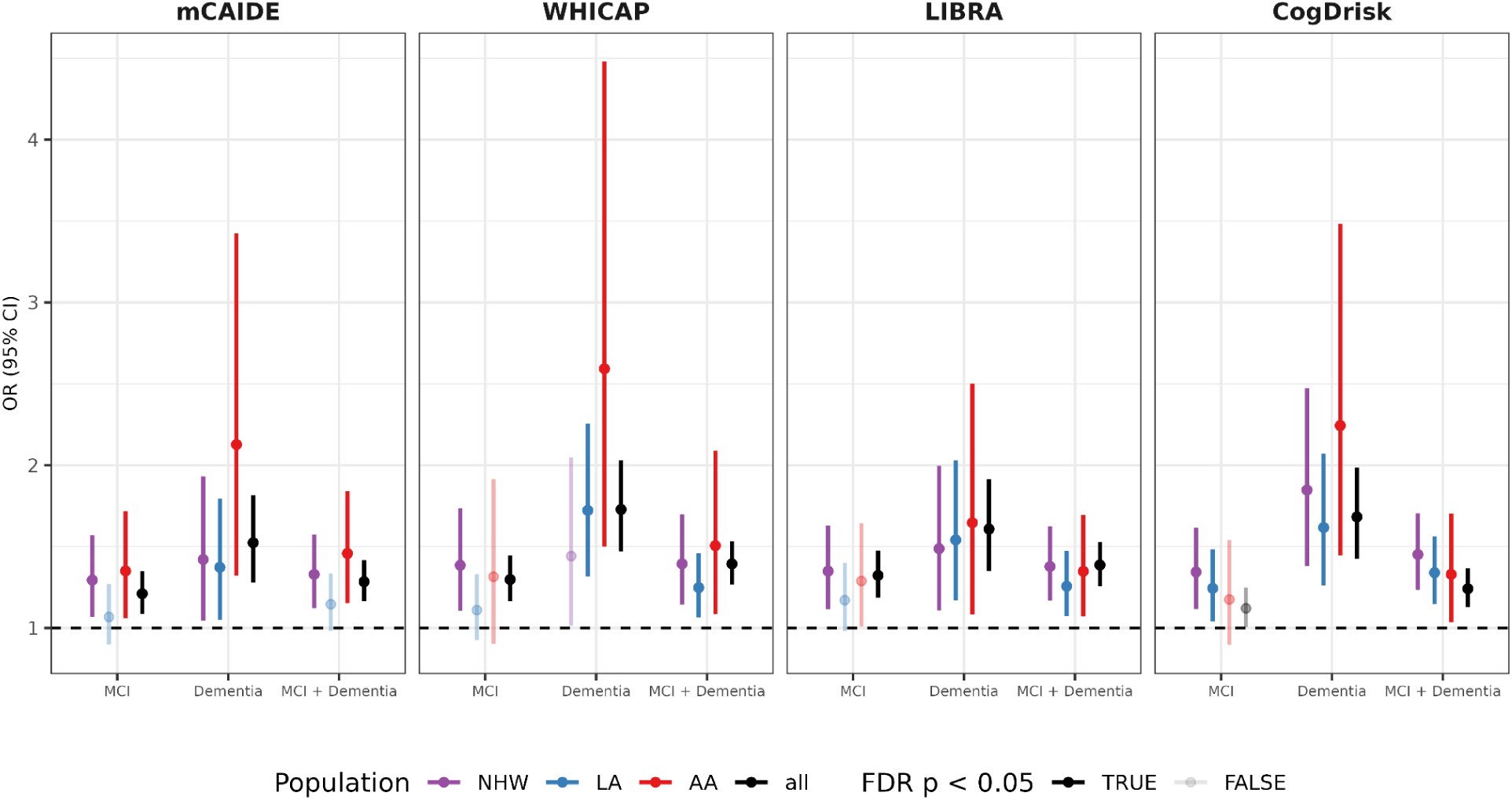
Associations of all CRS with MCI, dementia, and MCI + dementia diagnosis, with cognitively unimpaired participants as the reference group. Logistic regression analysis stratified by self-reported race/ethnicity.

These associations translated into only modest predictive performance. For dementia diagnosis, the demographics model had the highest AUC of 0.70 (R² = 0.070), which was comparable to the highest AUC achieved by CogDRisk (AUC = 0.65, R² = 0.046). All CRS had marginal differences in AUC, with mCAIDE having the lowest performance (AUC = 0.61, R² = 0.020) (Fig. 2, Table S3).

**Figure 2.**
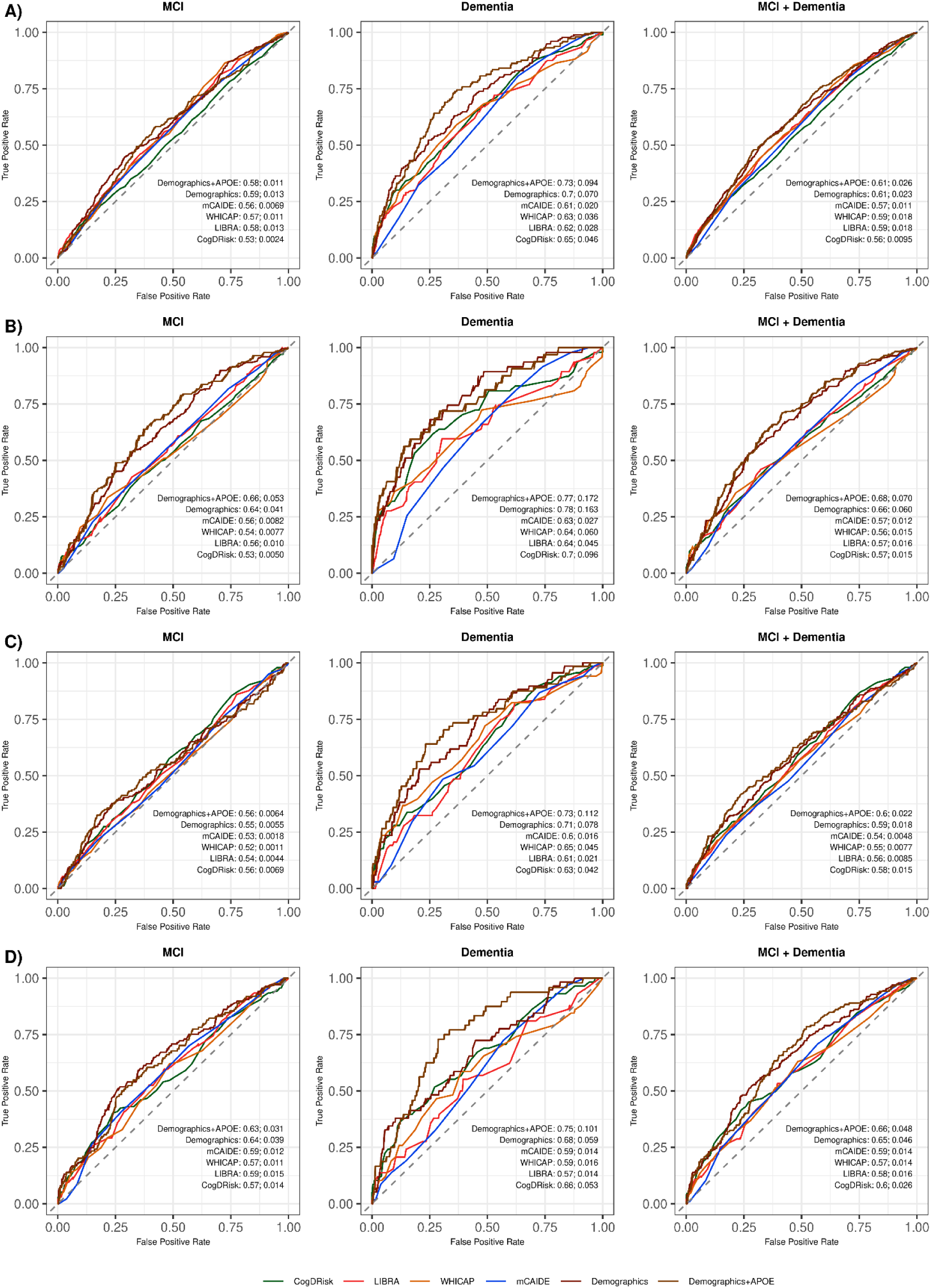
ROCAUC curve for CRS, demographics, and demographic + *APOE* model, stratified by self-reported race/ethnicity. The first number next to the model indicates the AUC value, followed by Nagelkerke R^2^.

In race/ethnicity-stratified analyses, higher CRS were associated with the highest odds of developing dementia in AA participants. WHICAP, though with the widest confidence interval, showed the highest association with dementia diagnosis in AA participants, with higher CRS associated with almost 160% higher odds of developing dementia. It was also the only CRS that did not demonstrate an association with odds of developing dementia in NHW participants (Fig 1, Table S2). Higher CogDRisk was associated with a 124% increase in the odds of developing dementia in the AA group, a 62% increase in the LA group, and an 85% increase in the NHW group relative to cognitively normal participants in their corresponding groups. Similarly, a higher LIBRA score was associated with a 65% increase in the odds of developing dementia in the AA group, a 54% increase in the LA group, and a 49% increase in the NHW group. mCAIDE was also associated with substantially higher odds in AA participants (113%) compared to LA (37%) and NHW (42%) participants (Fig. 1, Table S2). However, the z-test did not demonstrate any significant difference in effect size between the two racial/ethnic groups (Table S4).

Consistent with the overall results, model performance was only modest across racial/ethnic groups. The demographic model achieved the highest AUC in both the total cohort and across subgroups. CogDRisk achieved the highest AUC and R^2^ across racial/ethnic groups, ranging from 0.70 in AA participants to 0.66 in NHW participants and 0.63 in LA participants. A general pattern of higher AUC and R^2^ was observed in AA participants across the four CRS compared to NHW participants, who had the lowest AUC and R^2^ (Fig. 2, Table S3).

### 3.3 Higher clinical risk burden was associated with worsening AD endophenotypes

#### 3.3.1 mCAIDE

Higher mCAIDE scores were associated with increased levels of NfL and total Tau and decreased Aβ_42_/Aβ_40_ ratios in the total cohort, but not with pTau_181_ (Fig. 3, Table S2). In race/ethnicity-stratified analyses, associations with higher NfL and lower Aβ_42_/Aβ_40_ were significant only in LA participants (Fig. 3, Table S2). For neuroimaging outcomes, mCAIDE was strongly associated with smaller hippocampal volumes across race/ethnicity groups and decreased cortical thickness in LA and NHW participants, but not in AA participants (Fig. 3, Table S2). The effect sizes for mCAIDE on cortical thickness were statistically significantly different in LA participants compared to NHW participants (z = -3.79, p = 2.10e-3) (Table S4). For cognitive outcomes, higher mCAIDE scores were associated with lower MMSE performance and poorer verbal ability across racial groups (Fig. 3, Table S2). There was a statistically significant difference in the effect size for mCAIDE on memory between LA and NHW participants (z = 2.82, p = 0.039), but no other significant differences were observed (Table S4).

**Figure 3.**
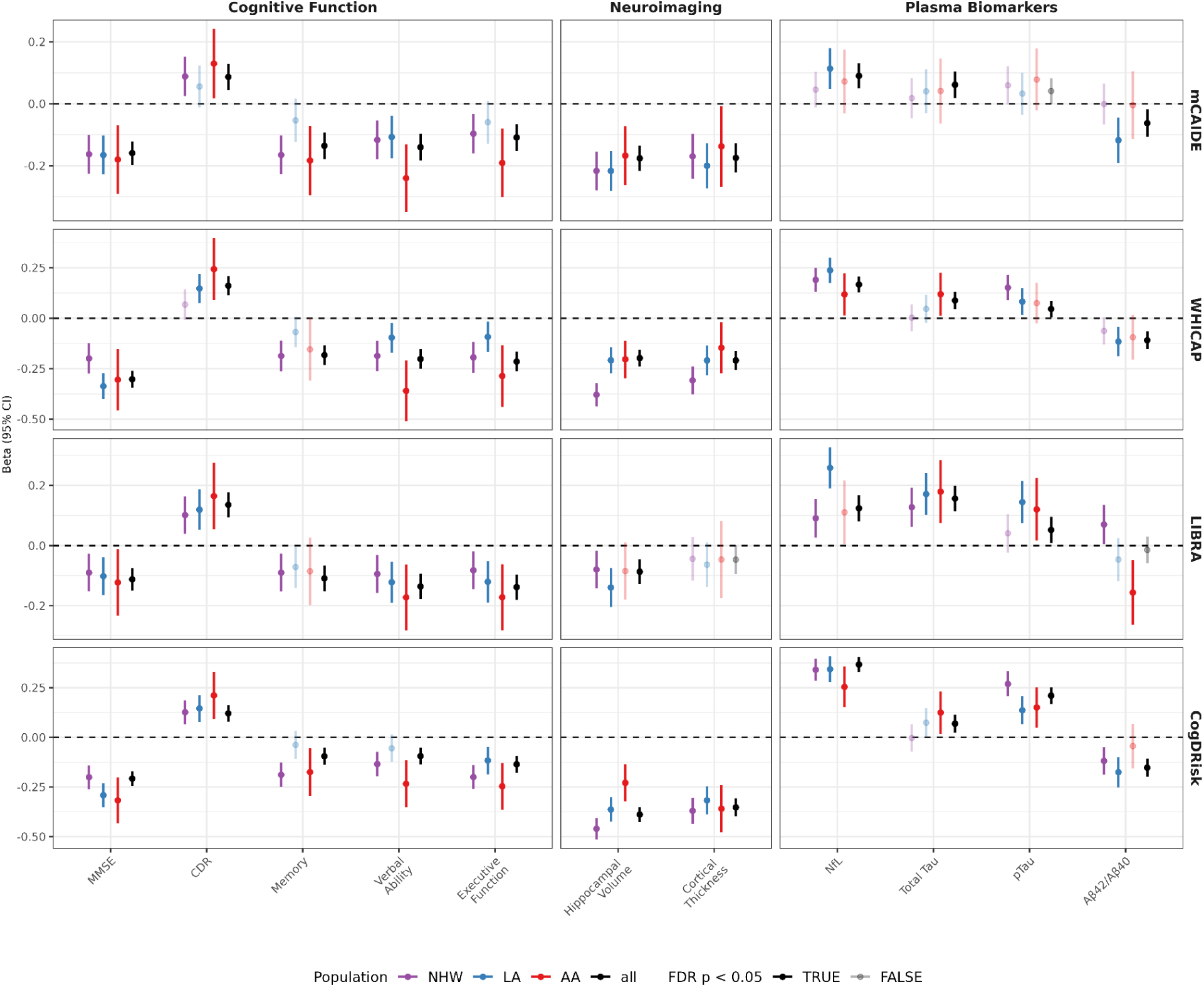
Associations of the four CRS with AD endophenotypes in the total cohort and stratified by self-reported race/ethnicity.

#### 3.3.2 WHICAP

The WHICAP score demonstrated similar trends to mCAIDE, with weaker or absent associations among LA and AA participants, specifically for plasma biomarker outcomes. In the total cohort, higher WHICAP scores were associated with increased NfL, pTau_181_, and total Tau, and decreased Aβ_42_/Aβ_40_. Across racial groups, NfL associations remained consistent, while other biomarkers varied (Fig. 3, Table S2). For neuroimaging outcomes, higher WHICAP was associated with decreased cortical thickness and decreased hippocampal volume (Fig. 3, Table S2). The magnitude of the effect size was larger in NHW compared to AA and LA (z = AA: 11.6, p < 1.0e-5; LA: 11.9, p < 1.0e-5) (Table S4). For cognitive outcomes, WHICAP was associated with verbal ability and MMSE, both in the total cohort and across all racial/ethnic groups (Fig. 3, Table S2).

#### 3.3.3 LIBRA

For plasma biomarkers, the LIBRA score was consistently associated with increased levels of total Tau and pTau_181_ and decreased levels of Aβ_42_/Aβ_40_, in AA participants. The effect of Aβ_42_/Aβ_40_ significantly differed between AA and NHW participants, with a negative association in AA and a positive association in NHW (z = -3.00, p = 2.54e-2) (Fig. 3, Table S2). Additionally, a higher LIBRA score was associated with substantially higher NfL levels in LA participants, with a significantly stronger positive association compared to the NHW participants (Table S4). For neuroimaging outcomes, a higher LIBRA score was associated with lower hippocampal volume in LA and NHW participants, but not in AA participants. A higher LIBRA score was not associated with either cortical thickness or WMH levels in any population (Fig. 3, Table S2). For clinical outcomes, a higher LIBRA score was associated with higher CDR and decreased executive function across racial/ethnic groups (Fig. 3, Table S2).

#### 3.3.4 CogDRisk

CogDRisk was the only CRS showing consistent associations across all AD endophenotypes, including plasma, neuroimaging, and cognitive domains, in the total cohort. In the total cohort, higher CogDRisk scores were significantly associated with reduced hippocampal volume and cortical thickness, higher levels of plasma biomarkers including total Tau, pTau_181_, and a lower Aβ_42_/Aβ_40_ ratio. Higher CogDRisk scores were also associated with poorer performance across all cognitive domains, including memory, verbal ability, and executive function, as well as with higher CDR scores and lower MMSE scores. (Fig. 3, Table S2).

For plasma biomarkers, a higher CogDRisk score was associated with increased levels of pTau_181_ in LA participants (Fig. 3, Table S2). This effect size was significantly greater in LA than in NHW participants (z = -2.91, p = 3.16e-2) (Table S4). Higher CogDRisk was not associated with levels of pTau_181_ or Aβ_42_/Aβ_40_ in AA participants. There was a significant difference in the effect size of CogDRisk on hippocampal volume between AA and NHW participants (z = 3.96, p = 1.23e-3), with higher CogDRisk showing larger effects in AA than in NHW participants, a pattern not observed for cortical thickness (Table S4). For cognitive outcomes, a higher CogDRisk score was consistently associated with decreased MMSE, increased CDR, and decreased executive function across racial/ethnic groups (Fig. 3, Table S2).

### 3.4 Model performance from sensitivity analysis

For the sensitivity analysis, we excluded demographic information (i.e., age, sex, and years of education) from mCAIDE, WHICAP, and CogDRisk scores to assess their contribution to model performance. For diagnostic outcomes, higher CRS was associated with increased odds of developing dementia in the total cohort across all diagnoses and all CRS, but this association diminished in race/ethnicity-stratified analysis (Table S5). This translated into reduced AUC and R^2^ values for all CRS (mCAIDE: AUC = 0.55, R^2^ = 0.0037; WHICAP: AUC = 0.53, R^2^ = 0.032; CogDRisk: AUC = 0.60, R^2^ = 0.015) (Table S6).

For endophenotypic outcomes, substantial decreases in the associations of all CRS were observed after excluding demographic information. In the total cohort, significant associations were observed specifically for cognitive outcome measurements, but did not translate consistently into neuroimaging and plasma biomarker outcomes. Higher CogDRisk was associated with increased levels of total Tau across all races/ethnicities, but all other associations with endophenotypes for all CRS diminished in sensitivity analysis (Table S5). Consistent with the overall results, the sensitivity analysis showcased decreased AUC and R^2^ across races/ethnicities. CogDRisk had the highest AUC for all races/ethnicities (AA: 0.63, LA: 0.57, NHW: 0.59) compared to WHICAP and mCAIDE, in which AUC ranged from 0.51 to 0.54 across races/ethnicities (Table S6).

## 4. DISCUSSION

In this study, we evaluated the associations of mCAIDE, WHICAP, LIBRA, and CogDRisk with cognitive impairment and AD endophenotypes in a cross-sectional, multiethnic cohort. Across the total cohort, higher CRS were associated with increased odds of dementia, although effect sizes varied by race/ethnicity. Among the scores, a higher CogDRisk score was consistently associated with higher odds of cognitive impairment and worse AD endophenotypes across racial groups. LIBRA also demonstrated consistent associations with AD endophenotypes, though with less consistency across races/ethnicities, which may be partly explained by the absence of demographic information in its scoring. WHICAP and mCAIDE performed similarly overall, with WHICAP having stronger and more consistent associations with plasma biomarkers. The lack of association between mCAIDE and dementia risk in NHW participants may reflect differences in the relevance of cardiometabolic risk factors captured by mCAIDE within this subgroup

Across AD endophenotypes, higher CRS scores were generally associated with more severe abnormalities, particularly in AA participants. This pattern may reflect underlying risks driven by unmeasured social determinants of health (SDoH) (e.g. socioeconomic status, neighbourhood and built environment, healthcare access) in the population. The wide confidence intervals in this population also suggest model instability due to a smaller sample size. Additionally, many significant z-test findings arose from comparisons between NHW and LA participants, which are likely attributable to larger standard errors and the lack of SDoH adjustment among AA participants. Overall, the variability in CRS performance across racial groups highlights the need to recalibrate risk models to better capture risk in historically underrepresented and disproportionately affected populations.

AUC and R^2^ for logistic regression were highest in the AA participants and lowest in the NHW participants, which was unanticipated given that CRS were largely developed in NHW/European descent populations. This pattern may be driven by the small sample size and the imbalance of case/control distributions in the AA group. Although the AA group included fewer total participants, it had a more balanced distribution of cases and controls compared to NHW participants, which can inflate apparent model performance in smaller samples. Because model performance metrics such as AUC and R^2^ are sensitive to outcome prevalence and class balance, groups with higher disease prevalence may exhibit more optimistic estimates despite smaller total sample sizes. Consistent with this, the prevalence of cognitive impairment was higher among AA participants (41.7%) than among LA (28.3%) or NHW participants (18.6%). In settings with limited sample sizes and more balanced outcome distributions, these conditions can lead to inflated performance estimates that may not generalize to broader populations. Together, these findings underscore the need for larger, more representative clinical datasets for underrepresented populations to ensure sufficient statistical power and disease prevalence distributions that mirror real-world patterns.

Consistent with previous studies, mCAIDE was the lowest-performing model among all, exhibiting weaker discriminatory power and associations^21,22,25^. CogDRisk score, on the other hand, consistently emerged as the best-performing model across outcomes, followed closely by LIBRA, which was also observed previously^21,24,25^. Superior performance of CogDRisk and LIBRA score is likely reflected by its broader incorporation of mid- to late-life cardiometabolic, lifestyle, and psychosocial risk and protective factors that are strongly linked to dementia risk (e.g., cognitive and social engagement, depression, diet) and amenable to intervention^45^. Therefore, this broader risk profiling enhances not only predictive performance but also informs targeted, multidomain prevention strategies to improve individuals’ clinical risk profiles. The LIBRA score has also shown comparable AUC and R^2^ to other CRS in sensitivity analysis, supporting its utility as a modifiable risk index. The consistent reporting on the performance of CogDRisk and LIBRA scores suggests that CRS integrating a wider range of clinical and lifestyle modifiable risk factors may offer improved model performance to demographic information alone.

Several limitations should be acknowledged. First, only four CRS were evaluated, which does not capture the full spectrum of both clinically relevant and community-based CRS that have been developed over the years^13^. Even among the four CRS that were evaluated, some CRS components had to be modified due to unavailable information in HABS-HD (e.g. diet, cognitive activity), though it is unlikely that the results would change significantly. Second, the cross-sectional study used in this paper limits causal inference (e.g. whether abnormalities in CRS affect plasma biomarkers or vice versa) and the ability to assess the temporal relationships of CRS and endophenotypes. Future studies should examine the longitudinal trajectories of biomarker changes associated with high baseline CRS to strengthen evidence that elevated CRS corresponds to abnormalities in AD pathology. Lastly, this study did not include participants from Native American/Alaska Native, Asian, or Native Hawaiian/Other Pacific Islander groups. Future work should extend these evaluations to cohorts that include these populations that reflect the broader racial and socioeconomic diversity of the US population.

Despite these limitations, this study contributes novel evidence to the development of clinical risk assessment tools. By incorporating racially diverse participants, we were able to evaluate the generalizability of CRS across populations that are often underrepresented in AD research. More importantly, this work advances the field by linking CRS to endophenotypes, particularly plasma biomarkers, which are minimally invasive and increasingly investigated for clinical implementation^6,46–49^.

In conclusion, the current study found that the CogDRisk score outperformed other CRS for both associations and model performance metrics compared to LIBRA, WHICAP, and mCAIDE, in which all three showed similar performances across different analyses. However, none of the CRS models outperformed the demographics or demographics + *APOE* genotype models, which is expected given that age, sex, years of education, and *APOE*\*ɛ4 allele are the strongest predictors. predictors of dementia risk. This underscores that CRS are most useful as low-cost, widely accessible tools for preliminary risk screening, rather than as standalone predictors, and highlights the potential value of combining them with other risk measures such as genetic screenings or biomarker assessments to create a multimodal approach that may improve overall accuracy and reliability. Taken together, these findings highlight both the promises and limitations of CRS and clarify the role of CRS as a supportive tool to complement established demographic and genetic predictors in dementia risk assessments.

## Data Availability

The primary data used in this study were obtained from the Health & Aging Brain Study - Health Disparities (HABSHD) through a formal data request process. These data are not publicly available but may be accessed through a data request submitted to the governing body of the University of North Texas Health Fort Worth (https://apps.unthsc.edu/itr/our
).

## ACKNOWLEDGEMENTS

The authors would like to thank all the participants, staff, researcher teams, and partners of the Health & Aging Brain Study - Health Disparities (HABS-HD). The HABS-HD is supported by the National Institute on Aging of the National Institutes of Health under Award Numbers R01AG054073, R01AG058533, R01AG070862, P41EB015922, and U19AG078109. The content is solely the responsibility of the authors and does not necessarily represent the official views of the National Institutes of Health. We gratefully acknowledge the contributions of our study partners and their families, whose help and participation made this work possible.

## CODE AVAILABILITY

All codes developed and used are available at https://github.com/AndrewsLabUCSF/CRS-analysis. Information regarding all the software and reference datasets used in the analysis can also be found in the repository.

## CONSENT STATEMENT

All aspects of the HABS-HD study protocol are managed by the North Texas Regional IRB. All HABS-HD partners (and/or legal guardians) undergo informed consent and provide written informed authorization to engage in the research study.

## REFERENCES

1. Kerwin, D. et al. Alzheimer’s disease diagnosis and management: Perspectives from around the world. Alzheimers Dement. Diagn. Assess. Dis. Monit. 14, e12334 (2022).

2. Seeher, K., Cataldi, R., Dua, T. & Kestel, D. Inequitable Access to Dementia Diagnosis and Care in Low-Resource Settings – A Global Perspective. Clin. Gerontol. 46, 133–137 (2023).

3. Global status report on the public health response to dementia. https://www.who.int/publications/i/item/9789240033245.

4. Jack, C. R. et al. Hypothetical model of dynamic biomarkers of the Alzheimer’s pathological cascade. Lancet Neurol. 9, 119–128 (2010).

5. Jack Jr., C. R., et al. Revised criteria for diagnosis and staging of Alzheimer’s disease: Alzheimer’s Association Workgroup. Alzheimers Dement. 20, 5143–5169 (2024).

6. Jack Jr., C. R., et al. NIA-AA Research Framework: Toward a biological definition of Alzheimer’s disease. Alzheimers Dement. 14, 535–562 (2018).

7. 2025 Alzheimer’s disease facts and figures - 2025 - Alzheimer’s & Dementia - Wiley Online Library. https://alz-journals.onlinelibrary.wiley.com/doi/10.1002/alz.70235.

8. Hinton, L., Tran, D., Peak, K., Meyer, O. L. & Quiñones, A. R. Mapping racial and ethnic healthcare disparities for persons living with dementia: A scoping review. Alzheimers Dement. 20, 3000–3020 (2024).

9. Chin, A. L., Negash, S. & Hamilton, R. Diversity and Disparity in Dementia: The Impact of Ethnoracial Differences in Alzheimer Disease. Alzheimer Dis. Assoc. Disord. 25, 187 (2011).

10. Anstey, K. J. et al. Dementia Risk Scores and Their Role in the Implementation of Risk Reduction Guidelines. Front. Neurol. 12, 765454 (2022).

11. Global action plan on the public health response to dementia 2017 - 2025. https://www.who.int/publications/i/item/global-action-plan-on-the-public-health-response-to-d ementia-2017---2025.

12. Risk Reduction of Cognitive Decline and Dementia: WHO Guidelines. (World Health Organization, Geneva, 2019).

13. Huque, M. H. et al. A critical review and classification of dementia risk assessment tools to inform dementia risk reduction. J. Prev. Alzheimers Dis. 12, 100333 (2025).

14. Tolea, M. I., Heo, J., Chrisphonte, S. & Galvin, J. E. A modified CAIDE (mCAIDE) risk score as a screening tool for cognitive impairment in older adults. J. Alzheimers Dis. JAD 82, 1755–1768 (2021).

15. Reitz, C. et al. A Summary Risk Score for the Prediction of Alzheimer Disease in Elderly Persons. Arch. Neurol. 67, 835–841 (2010).

16. Schiepers, O. J. G. et al. Lifestyle for Brain Health (LIBRA): a new model for dementia prevention. Int. J. Geriatr. Psychiatry 33, 167–175 (2018).

17. Anstey, K. J., Kootar, S., Huque, M. H., Eramudugolla, R. & Peters, R. Development of the CogDrisk tool to assess risk factors for dementia. Alzheimers Dement. Diagn. Assess. Dis. Monit. 14, e12336 (2022).

18. Kivipelto, M. et al. Risk score for the prediction of dementia risk in 20 years among middle aged people: a longitudinal, population-based study. Lancet Neurol. 5, 735–741 (2006).

19. Deckers, K. et al. Target risk factors for dementia prevention: a systematic review and Delphi consensus study on the evidence from observational studies. Int. J. Geriatr. Psychiatry 30, 234–246 (2015).

20. Anstey, K. J., Ee, N., Eramudugolla, R., Jagger, C. & Peters, R. A Systematic Review of Meta-Analyses that Evaluate Risk Factors for Dementia to Evaluate the Quantity, Quality, and Global Representativeness of Evidence. J. Alzheimer’s Dis. 70, S165–S186 (2019).

21. Huque, M. H. et al. CogDrisk, ANU-ADRI, CAIDE, and LIBRA Risk Scores for Estimating Dementia Risk. *JAMA Netw*. Open 6, e2331460 (2023).

22. Geethadevi, G. M. et al. Validity of three risk prediction models for dementia or cognitive impairment in Australia. Age Ageing 51, afac307 (2022).

23. Deckers, K. et al. Long-term dementia risk prediction by the LIBRA score: A 30-year follow-up of the CAIDE study. Int. J. Geriatr. Psychiatry 35, 195–203 (2020).

24. Huque, M. H. & Anstey, K. J. Assessment of dementia risk scores in predicting mild cognitive impairment: A comparison of CogDrisk, CAIDE, LIBRA, and ANU-ADRI. J. Prev. Alzheimers Dis. 12, 100324 (2025).

25. Stubs, J. et al. Dementia risk prediction: A comparative analysis of the ANU-ADRI, CAIDE, CogDrisk, LIBRA, and LIBRA2 indices in the HUNT study. J. Prev. Alzheimers Dis. 12, 100326 (2025).

26. Kootar, S. et al. Validation of the CogDrisk Instrument as Predictive of Dementia in Four General Community-Dwelling Populations. J. Prev. Alzheimers Dis. 10, 478–487 (2023).

27. Torres, S., Alexander, A., O’Bryant, S. & Medina, L. D. Cognition and the Predictive Utility of Three Risk Scores in an Ethnically Diverse Sample. J. Alzheimers Dis. JAD 75, 1049–1059 (2020).

28. Andrews, S. J. et al. Dementia risk scores, apolipoprotein E, and risk of Alzheimer’s disease: One size does not fit all. Alzheimers Dement. 20, 8595–8604 (2024).

29. Zetterberg, H. & Bendlin, B. B. Biomarkers for Alzheimer’s disease—preparing for a new era of disease-modifying therapies. Mol. Psychiatry 26, 296–308 (2021).

30. Jack, C. R. & Holtzman, D. M. Biomarker Modeling of Alzheimer’s Disease. Neuron 80, 1347–1358 (2013).

31. Jia, J. et al. Biomarker Changes during 20 Years Preceding Alzheimer’s Disease. N. Engl. J. Med. 390, 712–722 (2024).

32. Petersen, M. E. et al. Characterization of plasma AT(N) biomarkers among a racial and ethnically diverse community-based cohort: an HABS-HD study. Alzheimers Dement. Transl. Res. Clin. Interv. 11, e70045 (2025).

33. Timsina, J. et al. Harmonization of CSF and imaging biomarkers in Alzheimer’s disease: Need and practical applications for genetics studies and preclinical classification. Neurobiol. Dis. 190, 106373 (2024).

34. O’Bryant, S. E. et al. Neurodegeneration from the AT(N) framework is different among Mexican Americans compared to non-Hispanic Whites: A Health & Aging Brain among Latino Elders (HABLE) Study. Alzheimers Dement. Diagn. Assess. Dis. Monit. 14, e12267 (2022).

35. Wechsler, D. Wechsler Memory Scale (WMS-IV) Technical and Interpretive Manual ( 4th Ed.). (Pearson, San Antonio, TX, 2009).

36. González, H. M., Mungas, D. & Haan, M. N. A verbal learning and memory test for English-and Spanish-speaking older Mexican-American adults. Clin. Neuropsychol. 16, 439–451 (2002).

37. Lezak, M. D. Neuropsychological Assessment. (Oxford University Press, 2004).

38. Folstein, M. F., Folstein, S. E. & McHugh, P. R. “Mini-mental state”: A practical method for grading the cognitive state of patients for the clinician. J. Psychiatr. Res. 12, 189–198 (1975).

39. Berg, L. Clinical Dementia Rating (CDR). Psychopharmacol. Bull. 24, 637–639 (1988).

40. Topolski, T. D. et al. The Rapid Assessment of Physical Activity (RAPA) Among Older Adults. Prev. Chronic. Dis. 3, A118 (2006).

41. Anstey, K. J., Cherbuin, N. & Herath, P. M. Development of a New Method for Assessing Global Risk of Alzheimer’s Disease for Use in Population Health Approaches to Prevention. Prev. Sci. 14, 411–421 (2013).

42. Heymans, M. W. & Twisk, J. W. R. Handling missing data in clinical research. J. Clin. Epidemiol. 151, 185–188 (2022).

43. O’Bryant, S. E. et al. The Health & Aging Brain among Latino Elders (HABLE) study methods and participant characteristics. Alzheimers Dement. Diagn. Assess. Dis. Monit. 13, e12202 (2021).

44. Petersen, M. E. et al. Health and Aging Brain Study–Health Disparities (HABS-HD) methods and partner characteristics. Alzheimers Dement. Transl. Res. Clin. Interv. 11, e70140 (2025).

45. Mostert, C. M. et al. Broadening dementia risk models: building on the 2024 Lancet Commission report for a more inclusive global framework. eBioMedicine 120, (2025).

46. Alcolea, D. et al. Use of plasma biomarkers for AT(N) classification of neurodegenerative dementias. J. Neurol. Neurosurg. Psychiatry 92, 1206–1214 (2021).

47. Commissioner, O. of the. FDA Clears First Blood Test Used in Diagnosing Alzheimer’s Disease. FDA https://www.fda.gov/news-events/press-announcements/fda-clears-first-blood-test-used-diagnosing-alzheimers-disease (2025).

48. Karikari, T. K. Blood Tests for Alzheimer’s Disease: Increasing Efforts to Expand and Diversify Research Participation Is Critical for Widespread Validation and Acceptance. J. Alzheimers Dis. 90, 967–974.

49. Schöll, M. et al. Challenges in the practical implementation of blood biomarkers for Alzheimer’s disease. Lancet Healthy Longev. 5, (2024).

