## Supplemental materials for "Associations of dementia polyexposure scores to Alzheimer’s disease endophenotypes in diverse populations"

**Supplemental Content**

Table S1. Description of covariates used in mCAIDE, WHICAP, LIBRA, and CogDRisk scores

Table S2. Regression model estimates for original CRS and sensitivity analysis of CRS across diagnostic and AD endophenotype outcomes.

Table S3. AUC and $R^2$ performance metrics for original CRS and sensitivity analysis of CRS across diagnostic and AD endophenotype outcomes.

Table S4. Z-test comparisons of endophenotype coefficient estimates across racial groups.

Table S5. Regression model estimates for original CRS and sensitivity analysis of CRS across diagnostic and AD endophenotype outcomes using complete cases.

Table S6. AUC and $R^2$ performance metrics for original CRS and sensitivity analysis of CRS across diagnostic and AD endophenotype outcomes using complete cases.

Table S1. Description of covariates in the four CRS scoring.

| CRS | mCAIDE | WHICAP | LIBRA | CogDRisk |
|---|---|---|---|---|
| **Factors** | | | | |
| Age | ≥73 vs. 65-7s vs. <65 yrs | 65-70 vs. >70-75 vs. >75-80 vs. >80-85 vs. >85 yrs | | 60-64 vs. 65-69 vs. 70-74 vs. 75-79 vs. 80-84 vs. 85-89 vs. >90 yrs |
| Sex | Women vs Men | Women vs Men | | Women vs Men |
| Education | <12 vs. 12-16 vs. >16 yrs | 0-6 vs. 7-9 vs. >9 yrs | | <8 vs. 8-11 vs. > 11 yrs |
| Systolic Blood Pressure | ≥140 vs. <140 mm Hg | | | |
| Body Mass Index | >30 vs. ≤30 kg/m$^2$ | ≤25 vs. >25 | <30 vs. ≥30 kg/m$^2$ | |
| Dyslipidemia | Self-reported high cholesterol: Yes vs No | low HDL-C: Yes vs. No | Fasitng blood choletsterol ≥6.2 mmol/L or taking lipid-lowering medication | >6.5 mmol/L vs. < 6.5 mmol/L |
| Physical activitiy | miniPPT: <12 vs. ≥12 | | Recreational walking ≤1 hr per day or having <1 hr of sport or intensive leisure activity per week | Inactive vs. >150 min/week of moderate to vigorous activity |
| Hypertension | | Yes vs. No | ≥ 140/90 mm Hg and/or taking hypertensive medication | All combined high SBP, DBP, and/or diagnosis of hypertension at >65 yrs |
| Heart disease | | | Coronary heart disease | No atrial filbrillation vs. atrial fibrillation without stroke |
| Obesity | | | | Midlife (≤65 yrs) obesity (BMI underweight: < 18.5, normal: 18.5-14.9, overweight: 25-29.9, obese: ≥30 |
| Diabetes | | Yes vs. No | Medical history or current diagnosis of diabetes and/or taking diabetes medication | History of diabetes for women vs men |
| Alcohol | | | >0 and <14 units of alcohol per week | |
| Smoking | | Current smoker: Yes vs. No | Current smoker: Yes vs. No | Current smoker: Yes vs. No |

| | | | | |
|---|---|---|---|---|
| Depression | | | Clinical depressive symptoms: Yes vs. No | Centre for Epidemiological Studies Depression (CES-D) scale: No depression (CES-D ≥20) vs. depression (CES-D >20) |
| Stroke | | | | Hisotry of stroke: Yes vs. No |
| Traumatic brain injury | | | | Hisotry of TBI (with and without loss of conciousness): Yes vs. No |
| Renal dysfunction | | | estimated glomerular filtrate rate <60 mL/min/1.73m$^2$ | |
| Race | | Black vs. Hispanic vs. White | | |
| Healthy diet | | | Mediterranean diet or high unsaturated diet | Fish instake < 1 vs. ≥1 serving per week |
| Cognitivde engagement | | | High vs. low cognitive activity | Low vs. Middle vs. High |
| Social engagement | | | | Lonely vs. Not lonely |
| Insomnia | | | | Clinical diagnosis of insomnia: Yes vs. No |
| *APOE* ε4 | | ≥1 vs. none | | |

Table S2. Regression model estimates for original CRS and CRS sensitivity analysis for mCAIDE, WHICAP, and CogDRisk. For the sensitivity analysis, age, sex, and years of education covariates were excluded from the scoring.

| **Race** | **CRS** | **Outcome** | **P-value** | **P-value, FDR adj.** | **β** | **Lower Conf. Interval** | **Upper Conf. Interval** |
|---|---|---|---|---|---|---|---|
| Black | CogDRisk | Cortical Thickness | 1.05E-08 | 4.38E-08 | -3.60E-01 | -4.79E-01 | -2.40E-01 |
| Black | CogDRisk | MMSE | 1.34E-07 | 4.87E-07 | -3.17E-01 | -4.33E-01 | -2.01E-01 |
| Black | CogDRisk | Executive Function | 4.80E-05 | 1.13E-04 | -2.46E-01 | -3.64E-01 | -1.29E-01 |
| Black | CogDRisk | Verbal Ability | 1.15E-04 | 2.52E-04 | -2.34E-01 | -3.52E-01 | -1.16E-01 |
| Black | CogDRisk | Hippocampal Volume | 2.10E-06 | 6.15E-06 | -2.29E-01 | -3.23E-01 | -1.36E-01 |
| Black | CogDRisk | Memory | 4.40E-03 | 6.85E-03 | -1.75E-01 | -2.95E-01 | -5.50E-02 |
| Black | CogDRisk | Aβ42/Aβ40 | 4.45E-01 | 4.60E-01 | -4.38E-02 | -1.56E-01 | 6.89E-02 |
| Black | CogDRisk | Total Tau | 2.25E-02 | 3.02E-02 | 1.25E-01 | 1.77E-02 | 2.32E-01 |
| Black | CogDRisk | MCI | 2.43E-01 | 2.60E-01 | 1.30E-01 | -8.81E-02 | 3.51E-01 |
| Black | CogDRisk | pTau | 3.90E-03 | 6.11E-03 | 1.50E-01 | 4.86E-02 | 2.52E-01 |
| Black | CogDRisk | CDR | 5.49E-04 | 1.09E-03 | 2.11E-01 | 9.20E-02 | 3.30E-01 |
| Black | CogDRisk | Cog. Impair. | 2.41E-02 | 3.15E-02 | 2.44E-01 | 3.42E-02 | 4.60E-01 |
| Black | CogDRisk | NfL | 1.50E-06 | 4.66E-06 | 2.54E-01 | 1.52E-01 | 3.56E-01 |
| Black | CogDRisk | Dementia | 3.12E-04 | 6.34E-04 | 6.94E-01 | 3.26E-01 | 1.09E+00 |
| Black | LIBRA | Verbal Ability | 2.21E-03 | 3.68E-03 | -1.72E-01 | -2.82E-01 | -6.24E-02 |
| Black | LIBRA | Executive Function | 2.28E-03 | 3.74E-03 | -1.72E-01 | -2.82E-01 | -6.19E-02 |
| Black | LIBRA | Aβ42/Aβ40 | 4.58E-03 | 7.07E-03 | -1.56E-01 | -2.64E-01 | -4.86E-02 |
| Black | LIBRA | MMSE | 3.01E-02 | 3.85E-02 | -1.23E-01 | -2.33E-01 | -1.18E-02 |
| Black | LIBRA | Memory | 1.35E-01 | 1.52E-01 | -8.55E-02 | -1.98E-01 | 2.66E-02 |
| Black | LIBRA | Hippocampal Volume | 8.10E-02 | 9.58E-02 | -8.48E-02 | -1.80E-01 | 1.05E-02 |
| Black | LIBRA | Cortical Thickness | 4.75E-01 | 4.87E-01 | -4.66E-02 | -1.75E-01 | 8.17E-02 |
| Black | LIBRA | NfL | 4.10E-02 | 5.12E-02 | 1.10E-01 | 4.56E-03 | 2.16E-01 |
| Black | LIBRA | pTau | 2.37E-02 | 3.12E-02 | 1.20E-01 | 1.62E-02 | 2.24E-01 |
| Black | LIBRA | CDR | 3.62E-03 | 5.70E-03 | 1.65E-01 | 5.42E-02 | 2.75E-01 |
| Black | LIBRA | Total Tau | 8.16E-04 | 1.53E-03 | 1.79E-01 | 7.49E-02 | 2.84E-01 |
| Black | LIBRA | MCI | 4.14E-02 | 5.16E-02 | 2.31E-01 | 1.06E-02 | 4.55E-01 |
| Black | LIBRA | Cog. Impair. | 1.06E-02 | 1.51E-02 | 2.76E-01 | 6.65E-02 | 4.91E-01 |
| Black | LIBRA | Dementia | 1.92E-02 | 2.60E-02 | 4.70E-01 | 8.42E-02 | 8.76E-01 |
| Black | mCAIDE | Verbal Ability | 2.07E-05 | 5.27E-05 | -2.40E-01 | -3.50E-01 | -1.31E-01 |
| Black | mCAIDE | Executive Function | 7.71E-04 | 1.48E-03 | -1.91E-01 | -3.02E-01 | -8.02E-02 |
| Black | mCAIDE | Memory | 1.39E-03 | 2.47E-03 | -1.83E-01 | -2.95E-01 | -7.14E-02 |
| Black | mCAIDE | MMSE | 1.54E-03 | 2.71E-03 | -1.80E-01 | -2.91E-01 | -6.90E-02 |

| | | | | | | | |
|---|---|---|---|---|---|---|---|
| Black | mCAIDE | Hippocampal Volume | 5.93E-04 | 1.16E-03 | -1.67E-01 | -2.62E-01 | -7.25E-02 |
| Black | mCAIDE | Cortical Thickness | 3.87E-02 | 4.88E-02 | -1.38E-01 | -2.68E-01 | -7.18E-03 |
| Black | mCAIDE | Aβ42/Aβ40 | 9.39E-01 | 9.43E-01 | -4.28E-03 | -1.14E-01 | 1.05E-01 |
| Black | mCAIDE | Total Tau | 4.38E-01 | 4.55E-01 | 4.16E-02 | -6.38E-02 | 1.47E-01 |
| Black | mCAIDE | NfL | 1.70E-01 | 1.89E-01 | 7.21E-02 | -3.11E-02 | 1.75E-01 |
| Black | mCAIDE | pTau | 1.24E-01 | 1.40E-01 | 7.84E-02 | -2.15E-02 | 1.78E-01 |
| Black | mCAIDE | CDR | 2.32E-02 | 3.09E-02 | 1.30E-01 | 1.79E-02 | 2.42E-01 |
| Black | mCAIDE | MCI | 1.44E-02 | 2.00E-02 | 2.79E-01 | 5.76E-02 | 5.06E-01 |
| Black | mCAIDE | Cog. Impair. | 1.56E-03 | 2.71E-03 | 3.45E-01 | 1.34E-01 | 5.62E-01 |
| Black | mCAIDE | Dementia | 1.90E-03 | 3.21E-03 | 6.81E-01 | 2.66E-01 | 1.13E+00 |
| Black | WHICAP | Verbal Ability | 3.65E-06 | 1.05E-05 | -3.60E-01 | -5.10E-01 | -2.09E-01 |
| Black | WHICAP | MMSE | 9.75E-05 | 2.18E-04 | -3.05E-01 | -4.57E-01 | -1.53E-01 |
| Black | WHICAP | Executive Function | 2.56E-04 | 5.27E-04 | -2.86E-01 | -4.39E-01 | -1.34E-01 |
| Black | WHICAP | Hippocampal Volume | 2.25E-05 | 5.66E-05 | -2.04E-01 | -2.97E-01 | -1.11E-01 |
| Black | WHICAP | Memory | 5.19E-02 | 6.34E-02 | -1.54E-01 | -3.10E-01 | 1.30E-03 |
| Black | WHICAP | Cortical Thickness | 2.36E-02 | 3.12E-02 | -1.46E-01 | -2.73E-01 | -1.98E-02 |
| Black | WHICAP | Aβ42/Aβ40 | 9.60E-02 | 1.11E-01 | -9.41E-02 | -2.05E-01 | 1.68E-02 |
| Black | WHICAP | pTau | 1.48E-01 | 1.65E-01 | 7.50E-02 | -2.66E-02 | 1.77E-01 |
| Black | WHICAP | NfL | 2.61E-02 | 3.38E-02 | 1.18E-01 | 1.41E-02 | 2.22E-01 |
| Black | WHICAP | Total Tau | 2.90E-02 | 3.71E-02 | 1.19E-01 | 1.23E-02 | 2.26E-01 |
| Black | WHICAP | MCI | 1.52E-01 | 1.70E-01 | 1.61E-01 | -5.83E-02 | 3.85E-01 |
| Black | WHICAP | CDR | 2.01E-03 | 3.36E-03 | 2.43E-01 | 8.96E-02 | 3.97E-01 |
| Black | WHICAP | Cog. Impair. | 1.41E-02 | 1.97E-02 | 2.73E-01 | 5.91E-02 | 4.96E-01 |
| Black | WHICAP | Dementia | 6.36E-04 | 1.23E-03 | 6.69E-01 | 3.00E-01 | 1.08E+00 |
| Hispanic | CogDRisk | Hippocampal Volume | 5.52E-29 | 1.65E-27 | -3.63E-01 | -4.25E-01 | -3.02E-01 |
| Hispanic | CogDRisk | Cortical Thickness | 1.19E-17 | 1.32E-16 | -3.17E-01 | -3.88E-01 | -2.46E-01 |
| Hispanic | CogDRisk | MMSE | 3.08E-20 | 4.16E-19 | -2.92E-01 | -3.52E-01 | -2.31E-01 |
| Hispanic | CogDRisk | Aβ42/Aβ40 | 6.97E-06 | 1.92E-05 | -1.76E-01 | -2.52E-01 | -9.94E-02 |
| Hispanic | CogDRisk | Executive Function | 9.84E-04 | 1.78E-03 | -1.17E-01 | -1.86E-01 | -4.74E-02 |
| Hispanic | CogDRisk | Verbal Ability | 1.18E-01 | 1.35E-01 | -5.50E-02 | -1.24E-01 | 1.41E-02 |
| Hispanic | CogDRisk | Memory | 2.98E-01 | 3.14E-01 | -3.74E-02 | -1.08E-01 | 3.30E-02 |
| Hispanic | CogDRisk | Total Tau | 4.39E-02 | 5.43E-02 | 7.39E-02 | 2.04E-03 | 1.46E-01 |
| Hispanic | CogDRisk | pTau | 1.31E-04 | 2.81E-04 | 1.36E-01 | 6.67E-02 | 2.06E-01 |
| Hispanic | CogDRisk | CDR | 2.51E-05 | 6.27E-05 | 1.46E-01 | 7.82E-02 | 2.13E-01 |
| Hispanic | CogDRisk | MCI | 1.55E-02 | 2.12E-02 | 2.13E-01 | 4.02E-02 | 3.86E-01 |
| Hispanic | CogDRisk | Cog. Impair. | 1.93E-04 | 3.99E-04 | 2.93E-01 | 1.39E-01 | 4.47E-01 |

| | | | | | | | |
|---|---|---|---|---|---|---|---|
| Hispanic | CogDRisk | NfL | 3.08E-24 | 6.04E-23 | 3.43E-01 | 2.79E-01 | 4.07E-01 |
| Hispanic | CogDRisk | Dementia | 1.35E-04 | 2.87E-04 | 4.86E-01 | 2.36E-01 | 7.37E-01 |
| Hispanic | LIBRA | Hippocampal Volume | 3.03E-05 | 7.49E-05 | -1.39E-01 | -2.05E-01 | -7.42E-02 |
| Hispanic | LIBRA | Verbal Ability | 4.72E-04 | 9.45E-04 | -1.22E-01 | -1.90E-01 | -5.38E-02 |
| Hispanic | LIBRA | Executive Function | 6.09E-04 | 1.18E-03 | -1.20E-01 | -1.89E-01 | -5.17E-02 |
| Hispanic | LIBRA | MMSE | 1.54E-03 | 2.71E-03 | -1.02E-01 | -1.65E-01 | -3.90E-02 |
| Hispanic | LIBRA | Memory | 4.58E-02 | 5.64E-02 | -7.12E-02 | -1.41E-01 | -1.32E-03 |
| Hispanic | LIBRA | Cortical Thickness | 9.23E-02 | 1.08E-01 | -6.35E-02 | -1.37E-01 | 1.05E-02 |
| Hispanic | LIBRA | Aβ42/Aβ40 | 2.00E-01 | 2.17E-01 | -4.66E-02 | -1.18E-01 | 2.47E-02 |
| Hispanic | LIBRA | CDR | 5.12E-04 | 1.02E-03 | 1.20E-01 | 5.23E-02 | 1.87E-01 |
| Hispanic | LIBRA | pTau | 6.19E-05 | 1.43E-04 | 1.44E-01 | 7.41E-02 | 2.15E-01 |
| Hispanic | LIBRA | MCI | 8.74E-02 | 1.03E-01 | 1.53E-01 | -2.23E-02 | 3.28E-01 |
| Hispanic | LIBRA | Total Tau | 1.77E-06 | 5.34E-06 | 1.71E-01 | 1.01E-01 | 2.41E-01 |
| Hispanic | LIBRA | Cog. Impair. | 5.34E-03 | 8.03E-03 | 2.21E-01 | 6.63E-02 | 3.78E-01 |
| Hispanic | LIBRA | NfL | 3.01E-13 | 2.35E-12 | 2.59E-01 | 1.90E-01 | 3.27E-01 |
| Hispanic | LIBRA | Dementia | 2.39E-03 | 3.86E-03 | 4.16E-01 | 1.50E-01 | 6.88E-01 |
| Hispanic | mCAIDE | Hippocampal Volume | 7.57E-11 | 4.12E-10 | -2.17E-01 | -2.81E-01 | -1.52E-01 |
| Hispanic | mCAIDE | Cortical Thickness | 9.86E-08 | 3.68E-07 | -2.00E-01 | -2.73E-01 | -1.27E-01 |
| Hispanic | mCAIDE | MMSE | 2.64E-07 | 9.48E-07 | -1.66E-01 | -2.28E-01 | -1.03E-01 |
| Hispanic | mCAIDE | Aβ42/Aβ40 | 1.84E-03 | 3.14E-03 | -1.18E-01 | -1.92E-01 | -4.38E-02 |
| Hispanic | mCAIDE | Verbal Ability | 2.26E-03 | 3.74E-03 | -1.07E-01 | -1.76E-01 | -3.86E-02 |
| Hispanic | mCAIDE | Executive Function | 9.30E-02 | 1.08E-01 | -5.96E-02 | -1.29E-01 | 9.95E-03 |
| Hispanic | mCAIDE | Memory | 1.35E-01 | 1.52E-01 | -5.36E-02 | -1.24E-01 | 1.67E-02 |
| Hispanic | mCAIDE | pTau | 3.43E-01 | 3.59E-01 | 3.28E-02 | -3.51E-02 | 1.01E-01 |
| Hispanic | mCAIDE | Total Tau | 2.62E-01 | 2.77E-01 | 4.04E-02 | -3.02E-02 | 1.11E-01 |
| Hispanic | mCAIDE | CDR | 1.08E-01 | 1.23E-01 | 5.59E-02 | -1.22E-02 | 1.24E-01 |
| Hispanic | mCAIDE | MCI | 4.54E-01 | 4.67E-01 | 6.70E-02 | -1.08E-01 | 2.43E-01 |
| Hispanic | mCAIDE | NfL | 7.95E-04 | 1.50E-03 | 1.14E-01 | 4.74E-02 | 1.80E-01 |
| Hispanic | mCAIDE | Cog. Impair. | 7.47E-02 | 8.92E-02 | 1.42E-01 | -1.35E-02 | 2.98E-01 |
| Hispanic | mCAIDE | Dementia | 1.58E-02 | 2.15E-02 | 3.32E-01 | 6.56E-02 | 6.07E-01 |
| Hispanic | WHICAP | MMSE | 2.44E-23 | 4.44E-22 | -3.37E-01 | -4.01E-01 | -2.72E-01 |
| Hispanic | WHICAP | Cortical Thickness | 2.74E-08 | 1.07E-07 | -2.09E-01 | -2.82E-01 | -1.36E-01 |
| Hispanic | WHICAP | Hippocampal Volume | 4.48E-10 | 2.05E-09 | -2.08E-01 | -2.73E-01 | -1.43E-01 |
| Hispanic | WHICAP | Aβ42/Aβ40 | 1.68E-03 | 2.88E-03 | -1.16E-01 | -1.88E-01 | -4.36E-02 |
| Hispanic | WHICAP | Verbal Ability | 1.10E-02 | 1.56E-02 | -9.61E-02 | -1.70E-01 | -2.21E-02 |
| Hispanic | WHICAP | Executive Function | 1.53E-02 | 2.10E-02 | -9.23E-02 | -1.67E-01 | -1.78E-02 |

| | | | | | | | |
|---|---|---|---|---|---|---|---|
| Hispanic | WHICAP | Memory | 7.74E-02 | 9.19E-02 | -6.79E-02 | -1.43E-01 | 7.46E-03 |
| Hispanic | WHICAP | Total Tau | 1.88E-01 | 2.06E-01 | 4.63E-02 | -2.27E-02 | 1.15E-01 |
| Hispanic | WHICAP | pTau | 1.47E-02 | 2.04E-02 | 8.22E-02 | 1.62E-02 | 1.48E-01 |
| Hispanic | WHICAP | MCI | 2.58E-01 | 2.74E-01 | 1.01E-01 | -7.40E-02 | 2.76E-01 |
| Hispanic | WHICAP | CDR | 6.91E-05 | 1.57E-04 | 1.48E-01 | 7.52E-02 | 2.20E-01 |
| Hispanic | WHICAP | Cog. Impair. | 5.52E-03 | 8.22E-03 | 2.19E-01 | 6.49E-02 | 3.75E-01 |
| Hispanic | WHICAP | NfL | 4.02E-13 | 3.04E-12 | 2.37E-01 | 1.74E-01 | 3.01E-01 |
| Hispanic | WHICAP | Dementia | 7.28E-05 | 1.64E-04 | 5.40E-01 | 2.76E-01 | 8.11E-01 |
| NHW | CogDRisk | Hippocampal Volume | 1.14E-53 | 8.94E-52 | -4.60E-01 | -5.15E-01 | -4.06E-01 |
| NHW | CogDRisk | Cortical Thickness | 6.59E-26 | 1.43E-24 | -3.70E-01 | -4.36E-01 | -3.04E-01 |
| NHW | CogDRisk | MMSE | 8.61E-11 | 4.60E-10 | -2.01E-01 | -2.61E-01 | -1.41E-01 |
| NHW | CogDRisk | Executive Function | 1.01E-10 | 5.30E-10 | -2.00E-01 | -2.60E-01 | -1.40E-01 |
| NHW | CogDRisk | Memory | 2.01E-09 | 8.69E-09 | -1.89E-01 | -2.50E-01 | -1.28E-01 |
| NHW | CogDRisk | Verbal Ability | 1.53E-05 | 3.97E-05 | -1.35E-01 | -1.96E-01 | -7.39E-02 |
| NHW | CogDRisk | Aβ42/Aβ40 | 7.78E-04 | 1.48E-03 | -1.19E-01 | -1.88E-01 | -4.97E-02 |
| NHW | CogDRisk | Total Tau | 9.22E-01 | 9.33E-01 | -3.40E-03 | -7.17E-02 | 6.49E-02 |
| NHW | CogDRisk | CDR | 4.51E-05 | 1.08E-04 | 1.26E-01 | 6.59E-02 | 1.87E-01 |
| NHW | CogDRisk | pTau | 2.33E-16 | 2.12E-15 | 2.69E-01 | 2.06E-01 | 3.32E-01 |
| NHW | CogDRisk | MCI | 1.65E-03 | 2.85E-03 | 3.01E-01 | 1.14E-01 | 4.89E-01 |
| NHW | CogDRisk | NfL | 2.66E-30 | 9.02E-29 | 3.41E-01 | 2.84E-01 | 3.97E-01 |
| NHW | CogDRisk | Cog. Impair. | 5.37E-06 | 1.50E-05 | 3.86E-01 | 2.20E-01 | 5.53E-01 |
| NHW | CogDRisk | Dementia | 3.66E-05 | 8.88E-05 | 6.23E-01 | 3.29E-01 | 9.22E-01 |
| NHW | LIBRA | Verbal Ability | 3.26E-03 | 5.17E-03 | -9.43E-02 | -1.57E-01 | -3.16E-02 |
| NHW | LIBRA | MMSE | 4.81E-03 | 7.39E-03 | -9.01E-02 | -1.53E-01 | -2.75E-02 |
| NHW | LIBRA | Memory | 4.98E-03 | 7.61E-03 | -9.01E-02 | -1.53E-01 | -2.73E-02 |
| NHW | LIBRA | Executive Function | 1.05E-02 | 1.51E-02 | -8.22E-02 | -1.45E-01 | -1.93E-02 |
| NHW | LIBRA | Hippocampal Volume | 1.29E-02 | 1.82E-02 | -7.96E-02 | -1.42E-01 | -1.69E-02 |
| NHW | LIBRA | Cortical Thickness | 2.33E-01 | 2.50E-01 | -4.40E-02 | -1.16E-01 | 2.83E-02 |
| NHW | LIBRA | pTau | 2.12E-01 | 2.30E-01 | 4.08E-02 | -2.33E-02 | 1.05E-01 |
| NHW | LIBRA | Aβ42/Aβ40 | 3.48E-02 | 4.42E-02 | 7.00E-02 | 5.03E-03 | 1.35E-01 |
| NHW | LIBRA | NfL | 5.83E-03 | 8.59E-03 | 9.12E-02 | 2.64E-02 | 1.56E-01 |
| NHW | LIBRA | CDR | 1.38E-03 | 2.47E-03 | 1.02E-01 | 3.95E-02 | 1.64E-01 |
| NHW | LIBRA | Total Tau | 1.27E-04 | 2.75E-04 | 1.27E-01 | 6.25E-02 | 1.92E-01 |
| NHW | LIBRA | MCI | 1.91E-03 | 3.22E-03 | 3.00E-01 | 1.11E-01 | 4.91E-01 |
| NHW | LIBRA | Cog. Impair. | 1.38E-04 | 2.89E-04 | 3.25E-01 | 1.59E-01 | 4.94E-01 |
| NHW | LIBRA | Dementia | 8.06E-03 | 1.18E-02 | 3.97E-01 | 1.04E-01 | 6.94E-01 |
| NHW | mCAIDE | Hippocampal Volume | 2.15E-11 | 1.36E-10 | -2.17E-01 | -2.79E-01 | -1.54E-01 |

| | | | | | | | |
|---|---|---|---|---|---|---|---|
| NHW | mCAIDE | Cortical Thickness | 4.83E-06 | 1.37E-05 | -1.70E-01 | -2.42E-01 | -9.75E-02 |
| NHW | mCAIDE | Memory | 2.89E-07 | 1.01E-06 | -1.65E-01 | -2.28E-01 | -1.03E-01 |
| NHW | mCAIDE | MMSE | 4.05E-07 | 1.38E-06 | -1.63E-01 | -2.26E-01 | -1.00E-01 |
| NHW | mCAIDE | Verbal Ability | 3.04E-04 | 6.22E-04 | -1.17E-01 | -1.80E-01 | -5.35E-02 |
| NHW | mCAIDE | Executive Function | 2.98E-03 | 4.75E-03 | -9.65E-02 | -1.60E-01 | -3.29E-02 |
| NHW | mCAIDE | Aβ42/Aβ40 | 9.80E-01 | 9.80E-01 | -8.51E-04 | -6.66E-02 | 6.49E-02 |
| NHW | mCAIDE | Total Tau | 5.82E-01 | 5.92E-01 | 1.83E-02 | -4.68E-02 | 8.33E-02 |
| NHW | mCAIDE | NfL | 1.17E-01 | 1.34E-01 | 4.59E-02 | -1.16E-02 | 1.03E-01 |
| NHW | mCAIDE | pTau | 5.78E-02 | 6.99E-02 | 5.98E-02 | -1.99E-03 | 1.22E-01 |
| NHW | mCAIDE | CDR | 5.90E-03 | 8.66E-03 | 8.85E-02 | 2.56E-02 | 1.51E-01 |
| NHW | mCAIDE | MCI | 8.15E-03 | 1.18E-02 | 2.59E-01 | 6.78E-02 | 4.52E-01 |
| NHW | mCAIDE | Cog. Impair. | 9.44E-04 | 1.74E-03 | 2.85E-01 | 1.17E-01 | 4.55E-01 |
| NHW | mCAIDE | Dementia | 2.44E-02 | 3.18E-02 | 3.53E-01 | 4.74E-02 | 6.65E-01 |
| NHW | WHICAP | Hippocampal Volume | 6.78E-35 | 2.75E-33 | -3.79E-01 | -4.37E-01 | -3.21E-01 |
| NHW | WHICAP | Cortical Thickness | 9.94E-18 | 1.16E-16 | -3.08E-01 | -3.77E-01 | -2.39E-01 |
| NHW | WHICAP | MMSE | 2.77E-07 | 9.83E-07 | -1.99E-01 | -2.75E-01 | -1.24E-01 |
| NHW | WHICAP | Executive Function | 5.95E-07 | 1.98E-06 | -1.94E-01 | -2.70E-01 | -1.18E-01 |
| NHW | WHICAP | Memory | 1.99E-06 | 5.91E-06 | -1.87E-01 | -2.64E-01 | -1.10E-01 |
| NHW | WHICAP | Verbal Ability | 1.57E-06 | 4.85E-06 | -1.87E-01 | -2.62E-01 | -1.11E-01 |
| NHW | WHICAP | Aβ42/Aβ40 | 7.02E-02 | 8.45E-02 | -6.26E-02 | -1.30E-01 | 5.18E-03 |
| NHW | WHICAP | Total Tau | 9.39E-01 | 9.43E-01 | 2.63E-03 | -6.45E-02 | 6.98E-02 |
| NHW | WHICAP | CDR | 8.14E-02 | 9.59E-02 | 6.77E-02 | -8.45E-03 | 1.44E-01 |
| NHW | WHICAP | pTau | 2.69E-06 | 7.77E-06 | 1.52E-01 | 8.87E-02 | 2.15E-01 |
| NHW | WHICAP | NfL | 2.46E-10 | 1.22E-09 | 1.90E-01 | 1.32E-01 | 2.49E-01 |
| NHW | WHICAP | MCI | 4.50E-03 | 6.98E-03 | 2.67E-01 | 8.20E-02 | 4.52E-01 |
| NHW | WHICAP | Cog. Impair. | 9.53E-04 | 1.74E-03 | 2.76E-01 | 1.12E-01 | 4.40E-01 |
| NHW | WHICAP | Dementia | 4.09E-02 | 5.12E-02 | 3.01E-01 | 8.89E-03 | 5.87E-01 |
| all | CogDRisk | Hippocampal Volume | 5.76E-83 | 1.45E-80 | -3.89E-01 | -4.27E-01 | -3.51E-01 |
| all | CogDRisk | Cortical Thickness | 5.24E-51 | 3.14E-49 | -3.53E-01 | -3.97E-01 | -3.08E-01 |
| all | CogDRisk | MMSE | 6.50E-28 | 1.74E-26 | -2.08E-01 | -2.44E-01 | -1.71E-01 |
| all | CogDRisk | Aβ42/Aβ40 | 5.81E-11 | 3.36E-10 | -1.53E-01 | -1.98E-01 | -1.07E-01 |
| all | CogDRisk | Executive Function | 4.15E-10 | 1.94E-09 | -1.36E-01 | -1.78E-01 | -9.34E-02 |
| all | CogDRisk | Memory | 1.34E-05 | 3.51E-05 | -9.51E-02 | -1.38E-01 | -5.24E-02 |
| all | CogDRisk | Verbal Ability | 1.33E-05 | 3.51E-05 | -9.45E-02 | -1.37E-01 | -5.21E-02 |
| all | CogDRisk | Total Tau | 2.32E-03 | 3.75E-03 | 6.88E-02 | 2.46E-02 | 1.13E-01 |
| all | CogDRisk | MCI | 3.83E-02 | 4.85E-02 | 1.13E-01 | 5.75E-03 | 2.19E-01 |

| | | | | | | | |
|---|---|---|---|---|---|---|---|
| all | CogDRisk | CDR | 1.83E-08 | 7.37E-08 | 1.21E-01 | 7.88E-02 | 1.63E-01 |
| all | CogDRisk | pTau | 6.67E-23 | 1.12E-21 | 2.10E-01 | 1.69E-01 | 2.51E-01 |
| all | CogDRisk | Cog. Impair. | 7.98E-06 | 2.16E-05 | 2.18E-01 | 1.22E-01 | 3.14E-01 |
| all | CogDRisk | NfL | 2.06E-74 | 2.35E-72 | 3.67E-01 | 3.29E-01 | 4.05E-01 |
| all | CogDRisk | Dementia | 6.94E-10 | 3.14E-09 | 5.22E-01 | 3.56E-01 | 6.89E-01 |
| all | LIBRA | Executive Function | 2.13E-10 | 1.07E-09 | -1.38E-01 | -1.81E-01 | -9.58E-02 |
| all | LIBRA | Verbal Ability | 3.70E-10 | 1.76E-09 | -1.36E-01 | -1.78E-01 | -9.36E-02 |
| all | LIBRA | MMSE | 4.88E-09 | 2.07E-08 | -1.12E-01 | -1.50E-01 | -7.48E-02 |
| all | LIBRA | Memory | 6.05E-07 | 1.99E-06 | -1.09E-01 | -1.52E-01 | -6.63E-02 |
| all | LIBRA | Hippocampal Volume | 3.75E-05 | 9.02E-05 | -8.69E-02 | -1.28E-01 | -4.56E-02 |
| all | LIBRA | Cortical Thickness | 5.34E-02 | 6.49E-02 | -4.69E-02 | -9.45E-02 | 6.93E-04 |
| all | LIBRA | Aβ42/Aβ40 | 5.22E-01 | 5.33E-01 | -1.44E-02 | -5.83E-02 | 2.96E-02 |
| all | LIBRA | pTau | 1.81E-02 | 2.46E-02 | 5.21E-02 | 8.90E-03 | 9.53E-02 |
| all | LIBRA | NfL | 1.95E-08 | 7.74E-08 | 1.24E-01 | 8.10E-02 | 1.67E-01 |
| all | LIBRA | CDR | 2.67E-10 | 1.30E-09 | 1.36E-01 | 9.38E-02 | 1.78E-01 |
| all | LIBRA | Total Tau | 1.59E-12 | 1.08E-11 | 1.56E-01 | 1.13E-01 | 2.00E-01 |
| all | LIBRA | MCI | 4.65E-07 | 1.56E-06 | 2.79E-01 | 1.71E-01 | 3.88E-01 |
| all | LIBRA | Cog. Impair. | 5.91E-11 | 3.36E-10 | 3.29E-01 | 2.31E-01 | 4.28E-01 |
| all | LIBRA | Dementia | 1.03E-07 | 3.80E-07 | 4.75E-01 | 3.01E-01 | 6.51E-01 |
| all | mCAIDE | Hippocampal Volume | 6.12E-17 | 6.30E-16 | -1.76E-01 | -2.17E-01 | -1.35E-01 |
| all | mCAIDE | Cortical Thickness | 5.34E-13 | 3.92E-12 | -1.75E-01 | -2.22E-01 | -1.28E-01 |
| all | mCAIDE | MMSE | 1.78E-16 | 1.76E-15 | -1.59E-01 | -1.97E-01 | -1.22E-01 |
| all | mCAIDE | Verbal Ability | 1.74E-10 | 8.96E-10 | -1.40E-01 | -1.83E-01 | -9.72E-02 |
| all | mCAIDE | Memory | 7.57E-10 | 3.36E-09 | -1.36E-01 | -1.79E-01 | -9.27E-02 |
| all | mCAIDE | Executive Function | 8.21E-07 | 2.62E-06 | -1.09E-01 | -1.52E-01 | -6.57E-02 |
| all | mCAIDE | Aβ42/Aβ40 | 5.70E-03 | 8.44E-03 | -6.25E-02 | -1.07E-01 | -1.82E-02 |
| all | mCAIDE | pTau | 4.89E-02 | 6.00E-02 | 4.11E-02 | 1.89E-04 | 8.20E-02 |
| all | mCAIDE | Total Tau | 5.10E-03 | 7.75E-03 | 6.16E-02 | 1.85E-02 | 1.05E-01 |
| all | mCAIDE | CDR | 6.56E-05 | 1.50E-04 | 8.70E-02 | 4.43E-02 | 1.30E-01 |
| all | mCAIDE | NfL | 9.89E-06 | 2.64E-05 | 9.03E-02 | 5.04E-02 | 1.30E-01 |
| all | mCAIDE | MCI | 4.30E-04 | 8.68E-04 | 1.95E-01 | 8.67E-02 | 3.04E-01 |
| all | mCAIDE | Cog. Impair. | 3.31E-07 | 1.13E-06 | 2.56E-01 | 1.58E-01 | 3.54E-01 |
| all | mCAIDE | Dementia | 1.69E-06 | 5.17E-06 | 4.35E-01 | 2.59E-01 | 6.16E-01 |
| all | WHICAP | MMSE | 3.52E-44 | 1.70E-42 | -3.02E-01 | -3.44E-01 | -2.61E-01 |
| all | WHICAP | Executive Function | 8.25E-18 | 1.06E-16 | -2.15E-01 | -2.63E-01 | -1.66E-01 |
| all | WHICAP | Cortical Thickness | 9.01E-18 | 1.10E-16 | -2.09E-01 | -2.56E-01 | -1.62E-01 |

| | | | | | | | |
|---|---|---|---|---|---|---|---|
| all | WHICAP | Verbal Ability | 5.07E-16 | 4.45E-15 | -2.02E-01 | -2.50E-01 | -1.53E-01 |
| all | WHICAP | Hippocampal Volume | 1.10E-20 | 1.64E-19 | -1.97E-01 | -2.38E-01 | -1.56E-01 |
| all | WHICAP | Memory | 2.44E-13 | 1.96E-12 | -1.83E-01 | -2.31E-01 | -1.34E-01 |
| all | WHICAP | Aβ42/Aβ40 | 1.21E-06 | 3.83E-06 | -1.09E-01 | -1.53E-01 | -6.51E-02 |
| all | WHICAP | pTau | 2.78E-02 | 3.58E-02 | 4.58E-02 | 5.00E-03 | 8.66E-02 |
| all | WHICAP | Total Tau | 5.98E-05 | 1.39E-04 | 8.80E-02 | 4.51E-02 | 1.31E-01 |
| all | WHICAP | CDR | 6.90E-11 | 3.84E-10 | 1.61E-01 | 1.13E-01 | 2.09E-01 |
| all | WHICAP | NfL | 1.86E-16 | 1.77E-15 | 1.67E-01 | 1.28E-01 | 2.06E-01 |
| all | WHICAP | MCI | 1.93E-06 | 5.77E-06 | 2.60E-01 | 1.53E-01 | 3.68E-01 |
| all | WHICAP | Cog. Impair. | 5.98E-12 | 3.89E-11 | 3.42E-01 | 2.45E-01 | 4.40E-01 |
| all | WHICAP | Dementia | 2.50E-11 | 1.55E-10 | 5.72E-01 | 4.05E-01 | 7.41E-01 |
| Black | CogDRisk (sensitivity) | MMSE | 3.35E-01 | 4.66E-01 | -5.19E-02 | -1.58E-01 | 5.38E-02 |
| Black | CogDRisk (sensitivity) | Aβ42/Aβ40 | 1.13E-01 | 2.13E-01 | -8.76E-02 | -1.96E-01 | 2.09E-02 |
| Black | CogDRisk (sensitivity) | | 3.75E-01 | 5.04E-01 | -9.35E-02 | -3.02E-01 | 1.13E-01 |
| Black | CogDRisk (sensitivity) | Verbal Ability | 1.72E-01 | 2.96E-01 | -7.34E-02 | -1.79E-01 | 3.20E-02 |
| Black | CogDRisk (sensitivity) | pTau | 2.51E-01 | 3.77E-01 | -5.83E-02 | -1.58E-01 | 4.15E-02 |
| Black | CogDRisk (sensitivity) | Executive Function | 2.20E-01 | 3.52E-01 | -6.60E-02 | -1.72E-01 | 3.96E-02 |
| Black | CogDRisk (sensitivity) | Memory | 7.38E-01 | 8.45E-01 | -1.82E-02 | -1.25E-01 | 8.85E-02 |
| Black | CogDRisk (sensitivity) | Cortical Thickness | 2.18E-01 | 3.51E-01 | -8.05E-02 | -2.09E-01 | 4.79E-02 |
| Black | CogDRisk (sensitivity) | Hippocampal Volume | 9.33E-01 | 9.54E-01 | -4.12E-03 | -1.01E-01 | 9.28E-02 |
| Black | CogDRisk (sensitivity) | NfL | 5.78E-01 | 7.05E-01 | 2.92E-02 | -7.40E-02 | 1.32E-01 |
| Black | CogDRisk (sensitivity) | MCI | 7.48E-01 | 8.45E-01 | 3.57E-02 | -1.83E-01 | 2.54E-01 |
| Black | CogDRisk (sensitivity) | Cog. Impair. | 3.75E-01 | 5.04E-01 | 9.35E-02 | -1.13E-01 | 3.02E-01 |
| Black | CogDRisk (sensitivity) | Total Tau | 3.21E-03 | 1.36E-02 | 1.56E-01 | 5.27E-02 | 2.60E-01 |
| Black | CogDRisk (sensitivity) | CDR | 2.05E-02 | 5.84E-02 | 1.25E-01 | 1.94E-02 | 2.30E-01 |
| Black | CogDRisk (sensitivity) | Dementia | 6.67E-02 | 1.45E-01 | 3.51E-01 | -2.41E-02 | 7.30E-01 |
| Black | mCAIDE (sensitivity) | | 9.76E-02 | 1.92E-01 | -1.76E-01 | -3.87E-01 | 3.12E-02 |
| Black | mCAIDE (sensitivity) | Verbal Ability | 7.67E-03 | 2.77E-02 | -1.47E-01 | -2.54E-01 | -3.91E-02 |
| Black | mCAIDE (sensitivity) | Executive Function | 3.86E-02 | 9.65E-02 | -1.14E-01 | -2.22E-01 | -6.00E-03 |
| Black | mCAIDE (sensitivity) | MMSE | 5.04E-01 | 6.35E-01 | -3.70E-02 | -1.46E-01 | 7.18E-02 |
| Black | mCAIDE (sensitivity) | Hippocampal Volume | 2.41E-01 | 3.74E-01 | -5.69E-02 | -1.52E-01 | 3.84E-02 |
| Black | mCAIDE (sensitivity) | Memory | 4.92E-01 | 6.29E-01 | -3.84E-02 | -1.48E-01 | 7.13E-02 |

| | | | | | | | |
|---|---|---|---|---|---|---|---|
| Black | mCAIDE (sensitivity) | NfL | 5.95E-01 | 7.15E-01 | -2.80E-02 | -1.32E-01 | 7.56E-02 |
| Black | mCAIDE (sensitivity) | pTau | 7.79E-01 | 8.47E-01 | -1.43E-02 | -1.15E-01 | 8.60E-02 |
| Black | mCAIDE (sensitivity) | Cortical Thickness | 9.20E-01 | 9.53E-01 | -6.53E-03 | -1.35E-01 | 1.22E-01 |
| Black | mCAIDE (sensitivity) | Aβ42/Aβ40 | 9.98E-01 | 9.98E-01 | 1.33E-04 | -1.09E-01 | 1.09E-01 |
| Black | mCAIDE (sensitivity) | Total Tau | 1.17E-01 | 2.17E-01 | 8.39E-02 | -2.10E-02 | 1.89E-01 |
| Black | mCAIDE (sensitivity) | CDR | 2.73E-01 | 4.02E-01 | 6.09E-02 | -4.82E-02 | 1.70E-01 |
| Black | mCAIDE (sensitivity) | MCI | 2.27E-01 | 3.61E-01 | 1.35E-01 | -8.36E-02 | 3.57E-01 |
| Black | mCAIDE (sensitivity) | Cog. Impair. | 9.76E-02 | 1.92E-01 | 1.76E-01 | -3.12E-02 | 3.87E-01 |
| Black | mCAIDE (sensitivity) | Dementia | 4.77E-02 | 1.15E-01 | 4.01E-01 | 1.03E-02 | 8.09E-01 |
| Black | WHICAP (sensitivity) | Aβ42/Aβ40 | 1.03E-01 | 1.98E-01 | -9.05E-02 | -1.99E-01 | 1.82E-02 |
| Black | WHICAP (sensitivity) | | 3.47E-01 | 4.75E-01 | -9.97E-02 | -3.10E-01 | 1.07E-01 |
| Black | WHICAP (sensitivity) | Hippocampal Volume | 5.37E-02 | 1.23E-01 | -9.36E-02 | -1.89E-01 | 1.49E-03 |
| Black | WHICAP (sensitivity) | pTau | 9.56E-02 | 1.92E-01 | -8.47E-02 | -1.84E-01 | 1.50E-02 |
| Black | WHICAP (sensitivity) | Verbal Ability | 6.94E-02 | 1.50E-01 | -1.33E-01 | -2.76E-01 | 1.06E-02 |
| Black | WHICAP (sensitivity) | NfL | 5.27E-02 | 1.22E-01 | -1.02E-01 | -2.04E-01 | 1.20E-03 |
| Black | WHICAP (sensitivity) | Executive Function | 2.90E-01 | 4.14E-01 | -7.76E-02 | -2.22E-01 | 6.64E-02 |
| Black | WHICAP (sensitivity) | Cortical Thickness | 1.87E-01 | 3.16E-01 | 8.54E-02 | -4.17E-02 | 2.12E-01 |
| Black | WHICAP (sensitivity) | CDR | 7.49E-01 | 8.45E-01 | 2.36E-02 | -1.21E-01 | 1.68E-01 |
| Black | WHICAP (sensitivity) | Memory | 4.12E-01 | 5.45E-01 | 6.07E-02 | -8.46E-02 | 2.06E-01 |
| Black | WHICAP (sensitivity) | Dementia | 7.69E-01 | 8.45E-01 | 5.97E-02 | -3.31E-01 | 4.69E-01 |
| Black | WHICAP (sensitivity) | Total Tau | 2.88E-01 | 4.14E-01 | 5.70E-02 | -4.83E-02 | 1.62E-01 |
| Black | WHICAP (sensitivity) | MMSE | 5.80E-01 | 7.05E-01 | 4.07E-02 | -1.04E-01 | 1.85E-01 |
| Black | WHICAP (sensitivity) | Cog. Impair. | 3.47E-01 | 4.75E-01 | 9.97E-02 | -1.07E-01 | 3.10E-01 |
| Black | WHICAP (sensitivity) | MCI | 3.21E-01 | 4.50E-01 | 1.11E-01 | -1.07E-01 | 3.35E-01 |
| Hispanic | CogDRisk (sensitivity) | MMSE | 5.16E-02 | 1.21E-01 | -6.28E-02 | -1.26E-01 | 4.25E-04 |
| Hispanic | CogDRisk (sensitivity) | | 6.17E-02 | 1.37E-01 | -1.47E-01 | -3.02E-01 | 7.10E-03 |
| Hispanic | CogDRisk (sensitivity) | Hippocampal Volume | 1.35E-03 | 6.48E-03 | -1.08E-01 | -1.74E-01 | -4.21E-02 |
| Hispanic | CogDRisk (sensitivity) | Aβ42/Aβ40 | 3.57E-02 | 9.14E-02 | -7.73E-02 | -1.49E-01 | -5.18E-03 |
| Hispanic | CogDRisk (sensitivity) | Executive Function | 1.32E-02 | 3.99E-02 | -8.73E-02 | -1.56E-01 | -1.83E-02 |
| Hispanic | CogDRisk (sensitivity) | Verbal Ability | 1.45E-01 | 2.60E-01 | -5.11E-02 | -1.20E-01 | 1.76E-02 |

| | | | | | | | |
|---|---|---|---|---|---|---|---|
| Hispanic | CogDRisk (sensitivity) | Cortical Thickness | 1.46E-01 | 2.60E-01 | -5.52E-02 | -1.30E-01 | 1.92E-02 |
| Hispanic | CogDRisk (sensitivity) | pTau | 7.69E-01 | 8.45E-01 | 9.88E-03 | -5.61E-02 | 7.58E-02 |
| Hispanic | CogDRisk (sensitivity) | Memory | 4.56E-01 | 5.93E-01 | 2.66E-02 | -4.35E-02 | 9.68E-02 |
| Hispanic | CogDRisk (sensitivity) | CDR | 1.21E-02 | 3.83E-02 | 8.66E-02 | 1.90E-02 | 1.54E-01 |
| Hispanic | CogDRisk (sensitivity) | Total Tau | 5.94E-05 | 4.14E-04 | 1.40E-01 | 7.19E-02 | 2.08E-01 |
| Hispanic | CogDRisk (sensitivity) | MCI | 1.20E-01 | 2.22E-01 | 1.38E-01 | -3.63E-02 | 3.13E-01 |
| Hispanic | CogDRisk (sensitivity) | Cog. Impair. | 6.17E-02 | 1.37E-01 | 1.47E-01 | -7.10E-03 | 3.02E-01 |
| Hispanic | CogDRisk (sensitivity) | NfL | 1.85E-06 | 2.00E-05 | 1.57E-01 | 9.29E-02 | 2.21E-01 |
| Hispanic | CogDRisk (sensitivity) | Dementia | 2.05E-01 | 3.36E-01 | 1.69E-01 | -9.34E-02 | 4.29E-01 |
| Hispanic | mCAIDE (sensitivity) | MMSE | 1.86E-01 | 3.16E-01 | -4.26E-02 | -1.06E-01 | 2.06E-02 |
| Hispanic | mCAIDE (sensitivity) | Hippocampal Volume | 6.04E-03 | 2.40E-02 | -9.22E-02 | -1.58E-01 | -2.65E-02 |
| Hispanic | mCAIDE (sensitivity) | Verbal Ability | 8.03E-03 | 2.77E-02 | -9.27E-02 | -1.61E-01 | -2.42E-02 |
| Hispanic | mCAIDE (sensitivity) | Aβ42/Aβ40 | 4.24E-02 | 1.05E-01 | -7.49E-02 | -1.47E-01 | -2.58E-03 |
| Hispanic | mCAIDE (sensitivity) | Executive Function | 2.47E-01 | 3.74E-01 | -4.09E-02 | -1.10E-01 | 2.83E-02 |
| Hispanic | mCAIDE (sensitivity) | MCI | 7.61E-01 | 8.45E-01 | -2.72E-02 | -2.03E-01 | 1.48E-01 |
| Hispanic | mCAIDE (sensitivity) | Cortical Thickness | 1.59E-02 | 4.65E-02 | -9.10E-02 | -1.65E-01 | -1.71E-02 |
| Hispanic | mCAIDE (sensitivity) | | 9.27E-01 | 9.53E-01 | -7.18E-03 | -1.62E-01 | 1.47E-01 |
| Hispanic | mCAIDE (sensitivity) | pTau | 8.62E-01 | 9.19E-01 | -5.85E-03 | -7.21E-02 | 6.04E-02 |
| Hispanic | mCAIDE (sensitivity) | Memory | 9.82E-01 | 9.87E-01 | 7.97E-04 | -6.93E-02 | 7.09E-02 |
| Hispanic | mCAIDE (sensitivity) | CDR | 8.91E-01 | 9.44E-01 | 4.73E-03 | -6.31E-02 | 7.25E-02 |
| Hispanic | mCAIDE (sensitivity) | Cog. Impair. | 9.27E-01 | 9.53E-01 | 7.18E-03 | -1.47E-01 | 1.62E-01 |
| Hispanic | mCAIDE (sensitivity) | NfL | 3.69E-01 | 5.02E-01 | 2.98E-02 | -3.53E-02 | 9.50E-02 |
| Hispanic | mCAIDE (sensitivity) | Dementia | 5.22E-01 | 6.51E-01 | 8.56E-02 | -1.77E-01 | 3.49E-01 |
| Hispanic | mCAIDE (sensitivity) | Total Tau | 5.02E-03 | 2.08E-02 | 9.83E-02 | 2.97E-02 | 1.67E-01 |
| Hispanic | WHICAP (sensitivity) | Verbal Ability | 1.26E-02 | 3.93E-02 | -8.72E-02 | -1.56E-01 | -1.87E-02 |
| Hispanic | WHICAP (sensitivity) | Hippocampal Volume | 5.68E-02 | 1.29E-01 | -6.43E-02 | -1.30E-01 | 1.87E-03 |
| Hispanic | WHICAP (sensitivity) | | 5.04E-01 | 6.35E-01 | -5.31E-02 | -2.10E-01 | 1.01E-01 |
| Hispanic | WHICAP (sensitivity) | MMSE | 7.01E-01 | 8.21E-01 | -1.24E-02 | -7.57E-02 | 5.09E-02 |
| Hispanic | WHICAP (sensitivity) | Executive Function | 2.86E-01 | 4.14E-01 | -3.76E-02 | -1.07E-01 | 3.15E-02 |
| Hispanic | WHICAP (sensitivity) | Aβ42/Aβ40 | 6.42E-01 | 7.62E-01 | -1.71E-02 | -8.90E-02 | 5.49E-02 |

| | | | | | | | |
|---|---|---|---|---|---|---|---|
| Hispanic | WHICAP (sensitivity) | Memory | 7.69E-01 | 8.45E-01 | -1.05E-02 | -8.06E-02 | 5.95E-02 |
| Hispanic | WHICAP (sensitivity) | Cortical Thickness | 6.89E-01 | 8.12E-01 | -1.52E-02 | -8.96E-02 | 5.92E-02 |
| Hispanic | WHICAP (sensitivity) | pTau | 9.58E-01 | 9.72E-01 | 1.77E-03 | -6.39E-02 | 6.75E-02 |
| Hispanic | WHICAP (sensitivity) | MCI | 7.40E-01 | 8.45E-01 | 2.99E-02 | -1.45E-01 | 2.08E-01 |
| Hispanic | WHICAP (sensitivity) | NfL | 1.48E-01 | 2.61E-01 | 4.77E-02 | -1.70E-02 | 1.12E-01 |
| Hispanic | WHICAP (sensitivity) | Cog. Impair. | 5.04E-01 | 6.35E-01 | 5.31E-02 | -1.01E-01 | 2.10E-01 |
| Hispanic | WHICAP (sensitivity) | CDR | 2.88E-01 | 4.14E-01 | 3.67E-02 | -3.11E-02 | 1.04E-01 |
| Hispanic | WHICAP (sensitivity) | Total Tau | 7.10E-03 | 2.66E-02 | 9.36E-02 | 2.55E-02 | 1.62E-01 |
| Hispanic | WHICAP (sensitivity) | Dementia | 3.38E-01 | 4.68E-01 | 1.32E-01 | -1.32E-01 | 4.09E-01 |
| NHW | CogDRisk (sensitivity) | | 1.61E-03 | 7.42E-03 | -2.65E-01 | -4.30E-01 | -1.00E-01 |
| NHW | CogDRisk (sensitivity) | MMSE | 2.19E-02 | 6.10E-02 | -7.50E-02 | -1.39E-01 | -1.09E-02 |
| NHW | CogDRisk (sensitivity) | Hippocampal Volume | 8.05E-02 | 1.72E-01 | -5.56E-02 | -1.18E-01 | 6.77E-03 |
| NHW | CogDRisk (sensitivity) | Verbal Ability | 1.23E-01 | 2.26E-01 | -5.05E-02 | -1.15E-01 | 1.38E-02 |
| NHW | CogDRisk (sensitivity) | Executive Function | 1.12E-01 | 2.12E-01 | -5.22E-02 | -1.16E-01 | 1.21E-02 |
| NHW | CogDRisk (sensitivity) | Memory | 5.93E-01 | 7.15E-01 | -1.76E-02 | -8.21E-02 | 4.70E-02 |
| NHW | CogDRisk (sensitivity) | pTau | 9.15E-01 | 9.53E-01 | -3.31E-03 | -6.44E-02 | 5.78E-02 |
| NHW | CogDRisk (sensitivity) | Cortical Thickness | 9.61E-01 | 9.72E-01 | 1.77E-03 | -6.98E-02 | 7.34E-02 |
| NHW | CogDRisk (sensitivity) | NfL | 9.24E-01 | 9.53E-01 | 2.79E-03 | -5.42E-02 | 5.98E-02 |
| NHW | CogDRisk (sensitivity) | Aβ42/Aβ40 | 1.50E-02 | 4.47E-02 | 8.04E-02 | 1.56E-02 | 1.45E-01 |
| NHW | CogDRisk (sensitivity) | CDR | 2.56E-03 | 1.11E-02 | 9.80E-02 | 3.44E-02 | 1.62E-01 |
| NHW | CogDRisk (sensitivity) | Total Tau | 9.58E-06 | 8.21E-05 | 1.44E-01 | 8.06E-02 | 2.08E-01 |
| NHW | CogDRisk (sensitivity) | MCI | 2.02E-02 | 5.84E-02 | 2.22E-01 | 3.42E-02 | 4.10E-01 |
| NHW | CogDRisk (sensitivity) | Cog. Impair. | 1.61E-03 | 7.42E-03 | 2.65E-01 | 1.00E-01 | 4.30E-01 |
| NHW | CogDRisk (sensitivity) | Dementia | 1.01E-02 | 3.30E-02 | 3.73E-01 | 8.73E-02 | 6.59E-01 |
| NHW | mCAIDE (sensitivity) | MMSE | 1.21E-02 | 3.83E-02 | -8.11E-02 | -1.44E-01 | -1.78E-02 |
| NHW | mCAIDE (sensitivity) | | 8.89E-02 | 1.82E-01 | -1.44E-01 | -3.10E-01 | 2.23E-02 |
| NHW | mCAIDE (sensitivity) | Verbal Ability | 3.49E-02 | 9.05E-02 | -6.83E-02 | -1.32E-01 | -4.87E-03 |
| NHW | mCAIDE (sensitivity) | NfL | 8.61E-02 | 1.79E-01 | -4.96E-02 | -1.06E-01 | 7.05E-03 |
| NHW | mCAIDE (sensitivity) | Memory | 1.09E-01 | 2.08E-01 | -5.21E-02 | -1.16E-01 | 1.16E-02 |
| NHW | mCAIDE (sensitivity) | Hippocampal Volume | 1.90E-01 | 3.19E-01 | -4.20E-02 | -1.05E-01 | 2.08E-02 |

| | | | | | | | |
|---|---|---|---|---|---|---|---|
| NHW | mCAIDE (sensitivity) | pTau | 1.92E-01 | 3.20E-01 | -4.05E-02 | -1.01E-01 | 2.04E-02 |
| NHW | mCAIDE (sensitivity) | Executive Function | 4.19E-01 | 5.49E-01 | -2.63E-02 | -9.02E-02 | 3.76E-02 |
| NHW | mCAIDE (sensitivity) | Cortical Thickness | 6.32E-01 | 7.55E-01 | -1.76E-02 | -8.96E-02 | 5.45E-02 |
| NHW | mCAIDE (sensitivity) | CDR | 1.60E-01 | 2.80E-01 | 4.53E-02 | -1.79E-02 | 1.08E-01 |
| NHW | mCAIDE (sensitivity) | Aβ42/Aβ40 | 9.52E-02 | 1.92E-01 | 5.50E-02 | -9.62E-03 | 1.20E-01 |
| NHW | mCAIDE (sensitivity) | Total Tau | 8.93E-03 | 2.95E-02 | 8.52E-02 | 2.14E-02 | 1.49E-01 |
| NHW | mCAIDE (sensitivity) | MCI | 1.66E-01 | 2.89E-01 | 1.33E-01 | -5.62E-02 | 3.21E-01 |
| NHW | mCAIDE (sensitivity) | Cog. Impair. | 8.89E-02 | 1.82E-01 | 1.44E-01 | -2.23E-02 | 3.10E-01 |
| NHW | mCAIDE (sensitivity) | Dementia | 2.61E-01 | 3.89E-01 | 1.73E-01 | -1.31E-01 | 4.73E-01 |
| NHW | WHICAP (sensitivity) | NfL | 4.87E-05 | 3.67E-04 | -1.17E-01 | -1.74E-01 | -6.09E-02 |
| NHW | WHICAP (sensitivity) | Dementia | 5.15E-01 | 6.45E-01 | -9.67E-02 | -3.87E-01 | 1.96E-01 |
| NHW | WHICAP (sensitivity) | pTau | 5.72E-03 | 2.32E-02 | -8.58E-02 | -1.47E-01 | -2.50E-02 |
| NHW | WHICAP (sensitivity) | CDR | 2.98E-01 | 4.23E-01 | -3.96E-02 | -1.14E-01 | 3.50E-02 |
| NHW | WHICAP (sensitivity) | Verbal Ability | 2.44E-01 | 3.74E-01 | -4.47E-02 | -1.20E-01 | 3.05E-02 |
| NHW | WHICAP (sensitivity) | MMSE | 7.13E-01 | 8.31E-01 | -1.41E-02 | -8.91E-02 | 6.10E-02 |
| NHW | WHICAP (sensitivity) | | 8.05E-01 | 8.68E-01 | -2.09E-02 | -1.88E-01 | 1.45E-01 |
| NHW | WHICAP (sensitivity) | Hippocampal Volume | 9.18E-01 | 9.53E-01 | -3.37E-03 | -6.76E-02 | 6.08E-02 |
| NHW | WHICAP (sensitivity) | Cortical Thickness | 2.46E-01 | 3.74E-01 | 4.38E-02 | -3.02E-02 | 1.18E-01 |
| NHW | WHICAP (sensitivity) | Memory | 7.52E-01 | 8.45E-01 | 1.21E-02 | -6.31E-02 | 8.73E-02 |
| NHW | WHICAP (sensitivity) | Executive Function | 4.80E-01 | 6.18E-01 | 2.71E-02 | -4.82E-02 | 1.03E-01 |
| NHW | WHICAP (sensitivity) | Cog. Impair. | 8.05E-01 | 8.68E-01 | 2.09E-02 | -1.45E-01 | 1.88E-01 |
| NHW | WHICAP (sensitivity) | Aβ42/Aβ40 | 6.49E-02 | 1.43E-01 | 6.10E-02 | -3.79E-03 | 1.26E-01 |
| NHW | WHICAP (sensitivity) | Total Tau | 4.94E-02 | 1.18E-01 | 6.43E-02 | 1.67E-04 | 1.28E-01 |
| NHW | WHICAP (sensitivity) | MCI | 4.59E-01 | 5.94E-01 | 7.25E-02 | -1.18E-01 | 2.66E-01 |
| all | CogDRisk (sensitivity) | MMSE | 7.61E-05 | 5.12E-04 | -7.60E-02 | -1.14E-01 | -3.84E-02 |
| all | CogDRisk (sensitivity) | | 9.36E-06 | 8.21E-05 | -2.17E-01 | -3.14E-01 | -1.21E-01 |
| all | CogDRisk (sensitivity) | Executive Function | 5.87E-05 | 4.14E-04 | -8.77E-02 | -1.30E-01 | -4.50E-02 |
| all | CogDRisk (sensitivity) | Verbal Ability | 1.32E-03 | 6.48E-03 | -6.99E-02 | -1.12E-01 | -2.73E-02 |
| all | CogDRisk (sensitivity) | Hippocampal Volume | 8.02E-03 | 2.77E-02 | -5.61E-02 | -9.76E-02 | -1.47E-02 |
| all | CogDRisk (sensitivity) | pTau | 2.73E-01 | 4.02E-01 | -2.27E-02 | -6.32E-02 | 1.79E-02 |

| | | | | | | | |
|---|---|---|---|---|---|---|---|
| all | CogDRisk (sensitivity) | Memory | 3.12E-01 | 4.40E-01 | -2.21E-02 | -6.51E-02 | 2.08E-02 |
| all | CogDRisk (sensitivity) | Aβ42/Aβ40 | 4.13E-01 | 5.45E-01 | -1.83E-02 | -6.22E-02 | 2.56E-02 |
| all | CogDRisk (sensitivity) | Cortical Thickness | 1.97E-01 | 3.26E-01 | -3.14E-02 | -7.92E-02 | 1.64E-02 |
| all | CogDRisk (sensitivity) | NfL | 8.61E-04 | 4.80E-03 | 6.77E-02 | 2.79E-02 | 1.07E-01 |
| all | CogDRisk (sensitivity) | CDR | 4.11E-07 | 6.25E-06 | 1.09E-01 | 6.69E-02 | 1.51E-01 |
| all | CogDRisk (sensitivity) | Total Tau | 7.82E-14 | 1.47E-11 | 1.62E-01 | 1.20E-01 | 2.04E-01 |
| all | CogDRisk (sensitivity) | MCI | 1.08E-03 | 5.58E-03 | 1.78E-01 | 7.12E-02 | 2.85E-01 |
| all | CogDRisk (sensitivity) | Cog. Impair. | 9.36E-06 | 8.21E-05 | 2.17E-01 | 1.21E-01 | 3.14E-01 |
| all | CogDRisk (sensitivity) | Dementia | 1.41E-04 | 9.25E-04 | 3.27E-01 | 1.59E-01 | 4.96E-01 |
| all | mCAIDE (sensitivity) | MMSE | 3.52E-04 | 2.09E-03 | -6.86E-02 | -1.06E-01 | -3.10E-02 |
| all | mCAIDE (sensitivity) | | 1.10E-03 | 5.58E-03 | -1.61E-01 | -2.57E-01 | -6.43E-02 |
| all | mCAIDE (sensitivity) | Verbal Ability | 8.27E-07 | 1.01E-05 | -1.07E-01 | -1.49E-01 | -6.45E-02 |
| all | mCAIDE (sensitivity) | Executive Function | 7.56E-04 | 4.34E-03 | -7.35E-02 | -1.16E-01 | -3.08E-02 |
| all | mCAIDE (sensitivity) | Memory | 8.13E-03 | 2.77E-02 | -5.79E-02 | -1.01E-01 | -1.50E-02 |
| all | mCAIDE (sensitivity) | Hippocampal Volume | 3.45E-02 | 9.04E-02 | -4.46E-02 | -8.59E-02 | -3.26E-03 |
| all | mCAIDE (sensitivity) | pTau | 3.72E-02 | 9.40E-02 | -4.31E-02 | -8.37E-02 | -2.56E-03 |
| all | mCAIDE (sensitivity) | NfL | 2.32E-01 | 3.63E-01 | -2.43E-02 | -6.42E-02 | 1.56E-02 |
| all | mCAIDE (sensitivity) | Cortical Thickness | 8.55E-02 | 1.79E-01 | -4.17E-02 | -8.92E-02 | 5.83E-03 |
| all | mCAIDE (sensitivity) | Aβ42/Aβ40 | 8.25E-01 | 8.84E-01 | -4.95E-03 | -4.89E-02 | 3.90E-02 |
| all | mCAIDE (sensitivity) | CDR | 2.48E-02 | 6.79E-02 | 4.83E-02 | 6.13E-03 | 9.05E-02 |
| all | mCAIDE (sensitivity) | Total Tau | 6.36E-07 | 8.94E-06 | 1.08E-01 | 6.58E-02 | 1.51E-01 |
| all | mCAIDE (sensitivity) | MCI | 1.28E-02 | 3.93E-02 | 1.36E-01 | 2.90E-02 | 2.44E-01 |
| all | mCAIDE (sensitivity) | Cog. Impair. | 1.10E-03 | 5.58E-03 | 1.61E-01 | 6.43E-02 | 2.57E-01 |
| all | mCAIDE (sensitivity) | Dementia | 8.78E-03 | 2.95E-02 | 2.30E-01 | 5.81E-02 | 4.02E-01 |
| all | WHICAP (sensitivity) | | 1.93E-10 | 1.01E-08 | -3.32E-01 | -4.35E-01 | -2.31E-01 |
| all | WHICAP (sensitivity) | MMSE | 1.95E-06 | 2.00E-05 | -9.53E-02 | -1.35E-01 | -5.62E-02 |
| all | WHICAP (sensitivity) | Verbal Ability | 4.69E-10 | 1.92E-08 | -1.41E-01 | -1.85E-01 | -9.68E-02 |
| all | WHICAP (sensitivity) | pTau | 4.58E-09 | 1.17E-07 | -1.21E-01 | -1.61E-01 | -8.07E-02 |
| all | WHICAP (sensitivity) | Executive Function | 2.37E-09 | 6.98E-08 | -1.36E-01 | -1.80E-01 | -9.13E-02 |
| all | WHICAP (sensitivity) | NfL | 5.71E-08 | 1.28E-06 | -1.10E-01 | -1.50E-01 | -7.07E-02 |

| | | | | | | | |
|---|---|---|---|---|---|---|---|
| all | WHICAP (sensitivity) | Memory | 1.06E-06 | 1.21E-05 | -1.11E-01 | -1.56E-01 | -6.66E-02 |
| all | WHICAP (sensitivity) | Cortical Thickness | 1.02E-01 | 1.98E-01 | 3.97E-02 | -7.90E-03 | 8.74E-02 |
| all | WHICAP (sensitivity) | Aβ42/Aβ40 | 7.72E-01 | 8.45E-01 | 6.50E-03 | -3.76E-02 | 5.06E-02 |
| all | WHICAP (sensitivity) | Hippocampal Volume | 3.91E-01 | 5.21E-01 | 1.81E-02 | -2.33E-02 | 5.96E-02 |
| all | WHICAP (sensitivity) | Total Tau | 3.84E-06 | 3.73E-05 | 1.01E-01 | 5.82E-02 | 1.44E-01 |
| all | WHICAP (sensitivity) | CDR | 3.14E-04 | 1.92E-03 | 8.09E-02 | 3.70E-02 | 1.25E-01 |
| all | WHICAP (sensitivity) | Dementia | 2.50E-03 | 1.11E-02 | 2.76E-01 | 9.94E-02 | 4.58E-01 |
| all | WHICAP (sensitivity) | Cog. Impair. | 1.93E-10 | 1.01E-08 | 3.32E-01 | 2.31E-01 | 4.35E-01 |
| all | WHICAP (sensitivity) | MCI | 9.97E-10 | 3.48E-08 | 3.60E-01 | 2.46E-01 | 4.77E-01 |

Table S3. Model performance metrics for logistic regression for diagnostic outcomes (AUC & Nagelkerke $R^2$) and linear regression for AD endophenotype outcomes ($R^2$) for demographics (age, sex, and years of education), demographics + *APOE* genotype, CRS, and CRS for sensitivity analysis.

| **Model** | **Race** | **Outcome** | **AUC** | **Nagelkerke $R^2$** | **$R^2$** |
|---|---|---|---|---|---|
| Demographics + APOE | Black | MCI | 0.66 | 5.32E-02 | |
| Demographics + APOE | Black | Dementia | 0.77 | 1.72E-01 | |
| Demographics + APOE | Black | Cog. Impair. | 0.68 | 6.97E-02 | |
| Demographics + APOE | Black | Executive Function | | | 6.77E-02 |
| Demographics + APOE | Black | Memory | | | 1.19E-01 |
| Demographics + APOE | Black | Verbal Ability | | | 1.04E-01 |
| Demographics + APOE | Black | MMSE | | | 2.10E-01 |
| Demographics + APOE | Black | CDR | | | 7.59E-02 |
| Demographics + APOE | Black | Aβ42/Aβ40 | | | 3.03E-02 |
| Demographics + APOE | Black | Total Tau | | | 1.17E-01 |
| Demographics + APOE | Black | pTau | | | 1.61E-01 |
| Demographics + APOE | Black | NfL | | | 1.40E-01 |
| Demographics + APOE | Black | Cortical Thickness | | | 1.37E-01 |
| Demographics + APOE | Black | Hippo. Volume | | | 3.27E-01 |
| CogDRisk | Black | MCI | 0.53 | 4.99E-03 | |
| CogDRisk | Black | Dementia | 0.70 | 9.63E-02 | |
| CogDRisk | Black | Cog. Impair. | 0.57 | 1.49E-02 | |
| CogDRisk | Black | Executive Function | | | 4.35E-02 |
| CogDRisk | Black | Memory | | | 2.27E-02 |
| CogDRisk | Black | Verbal Ability | | | 4.19E-02 |
| CogDRisk | Black | MMSE | | | 8.14E-02 |
| CogDRisk | Black | CDR | | | 5.52E-02 |
| CogDRisk | Black | Aβ42/Aβ40 | | | 1.80E-02 |
| CogDRisk | Black | Total Tau | | | 6.06E-02 |
| CogDRisk | Black | pTau | | | 1.36E-01 |
| CogDRisk | Black | NfL | | | 2.28E-01 |
| CogDRisk | Black | Cortical Thickness | | | 1.65E-01 |
| CogDRisk | Black | Hippo. Volume | | | 2.59E-01 |
| CogDRisk (sensitivity) | Black | MCI | 0.51 | 4.32E-04 | |
| CogDRisk (sensitivity) | Black | Dementia | 0.63 | 1.73E-02 | |
| CogDRisk (sensitivity) | Black | Cog. Impair. | 0.53 | 2.01E-03 | |

| | | | | | |
|---|---|---|---|---|---|
| CogDRisk (sensitivity) | Black | Executive Function | | | 1.80E-03 |
| CogDRisk (sensitivity) | Black | Memory | | | 1.28E-04 |
| CogDRisk (sensitivity) | Black | Verbal Ability | | | 3.02E-03 |
| CogDRisk (sensitivity) | Black | MMSE | | | 2.03E-03 |
| CogDRisk (sensitivity) | Black | CDR | | | 1.65E-02 |
| CogDRisk (sensitivity) | Black | Aβ42/Aβ40 | | | 1.39E-02 |
| CogDRisk (sensitivity) | Black | Total Tau | | | 5.39E-02 |
| CogDRisk (sensitivity) | Black | pTau | | | 8.36E-02 |
| CogDRisk (sensitivity) | Black | NfL | | | 1.20E-01 |
| CogDRisk (sensitivity) | Black | Cortical Thickness | | | 2.46E-02 |
| CogDRisk (sensitivity) | Black | Hippo. Volume | | | 1.49E-01 |
| LIBRA | Black | MCI | 0.56 | 1.02E-02 | |
| LIBRA | Black | Dementia | 0.64 | 4.50E-02 | |
| LIBRA | Black | Cog. Impair. | 0.57 | 1.58E-02 | |
| LIBRA | Black | Executive Function | | | 1.94E-02 |
| LIBRA | Black | Memory | | | 5.53E-03 |
| LIBRA | Black | Verbal Ability | | | 1.40E-02 |
| LIBRA | Black | MMSE | | | 1.12E-02 |
| LIBRA | Black | CDR | | | 3.02E-02 |
| LIBRA | Black | Aβ42/Aβ40 | | | 2.18E-02 |
| LIBRA | Black | Total Tau | | | 2.26E-02 |
| LIBRA | Black | pTau | | | 9.97E-03 |
| LIBRA | Black | NfL | | | 9.44E-03 |
| LIBRA | Black | Cortical Thickness | | | 2.37E-02 |
| LIBRA | Black | Hippo. Volume | | | 1.61E-01 |
| LIBRA (sensitivity) | Black | MCI | 0.59 | 1.97E-02 | |
| LIBRA (sensitivity) | Black | Dementia | 0.72 | 1.06E-01 | |
| LIBRA (sensitivity) | Black | Cog. Impair. | 0.61 | 3.11E-02 | |
| LIBRA (sensitivity) | Black | Executive Function | | | 5.34E-02 |
| LIBRA (sensitivity) | Black | Memory | | | 2.30E-02 |
| LIBRA (sensitivity) | Black | Verbal Ability | | | 4.01E-02 |
| LIBRA (sensitivity) | Black | MMSE | | | 5.33E-02 |
| LIBRA (sensitivity) | Black | CDR | | | 6.62E-02 |
| LIBRA (sensitivity) | Black | Aβ42/Aβ40 | | | 2.75E-02 |
| LIBRA (sensitivity) | Black | Total Tau | | | 2.89E-02 |
| LIBRA (sensitivity) | Black | pTau | | | 5.50E-02 |

| | | | | | |
|---|---|---|---|---|---|
| LIBRA (sensitivity) | Black | NfL | | | 6.36E-02 |
| LIBRA (sensitivity) | Black | Cortical Thickness | | | 8.65E-02 |
| LIBRA (sensitivity) | Black | Hippo. Volume | | | 2.19E-01 |
| mCAIDE | Black | MCI | 0.56 | 8.18E-03 | |
| mCAIDE | Black | Dementia | 0.63 | 2.70E-02 | |
| mCAIDE | Black | Cog. Impair. | 0.57 | 1.20E-02 | |
| mCAIDE | Black | Executive Function | | | 1.90E-02 |
| mCAIDE | Black | Memory | | | 1.88E-02 |
| mCAIDE | Black | Verbal Ability | | | 3.24E-02 |
| mCAIDE | Black | MMSE | | | 2.10E-02 |
| mCAIDE | Black | CDR | | | 1.21E-02 |
| mCAIDE | Black | Aβ42/Aβ40 | | | 7.22E-03 |
| mCAIDE | Black | Total Tau | | | 3.53E-02 |
| mCAIDE | Black | pTau | | | 9.99E-02 |
| mCAIDE | Black | NfL | | | 1.28E-01 |
| mCAIDE | Black | Cortical Thickness | | | 5.96E-02 |
| mCAIDE | Black | Hippo. Volume | | | 1.88E-01 |
| mCAIDE (sensitivity) | Black | MCI | 0.52 | 7.94E-04 | |
| mCAIDE (sensitivity) | Black | Dementia | 0.51 | 4.29E-04 | |
| mCAIDE (sensitivity) | Black | Cog. Impair. | 0.52 | 7.56E-04 | |
| mCAIDE (sensitivity) | Black | Executive Function | | | 2.19E-03 |
| mCAIDE (sensitivity) | Black | Memory | | | 9.95E-05 |
| mCAIDE (sensitivity) | Black | Verbal Ability | | | 7.65E-03 |
| mCAIDE (sensitivity) | Black | MMSE | | | 8.17E-05 |
| mCAIDE (sensitivity) | Black | CDR | | | 1.63E-04 |
| mCAIDE (sensitivity) | Black | Aβ42/Aβ40 | | | 6.57E-03 |
| mCAIDE (sensitivity) | Black | Total Tau | | | 4.14E-02 |
| mCAIDE (sensitivity) | Black | pTau | | | 8.35E-02 |
| mCAIDE (sensitivity) | Black | NfL | | | 1.18E-01 |
| mCAIDE (sensitivity) | Black | Cortical Thickness | | | 2.22E-02 |
| mCAIDE (sensitivity) | Black | Hippo. Volume | | | 1.50E-01 |
| WHICAP | Black | MCI | 0.54 | 7.67E-03 | |
| WHICAP | Black | Dementia | 0.64 | 6.04E-02 | |
| WHICAP | Black | Cog. Impair. | 0.56 | 1.47E-02 | |
| WHICAP | Black | Executive Function | | | 2.54E-02 |
| WHICAP | Black | Memory | | | 7.52E-03 |

| | | | | | |
|---|---|---|---|---|---|
| WHICAP | Black | Verbal Ability | | | 4.45E-02 |
| WHICAP | Black | MMSE | | | 4.36E-02 |
| WHICAP | Black | CDR | | | 3.20E-02 |
| WHICAP | Black | Aβ42/Aβ40 | | | 2.18E-02 |
| WHICAP | Black | Total Tau | | | 4.24E-02 |
| WHICAP | Black | pTau | | | 9.54E-02 |
| WHICAP | Black | NfL | | | 1.34E-01 |
| WHICAP | Black | Cortical Thickness | | | 5.53E-02 |
| WHICAP | Black | Hippo. Volume | | | 2.06E-01 |
| WHICAP (sensitivity) | Black | MCI | 0.51 | 2.14E-03 | |
| WHICAP (sensitivity) | Black | Dementia | 0.52 | 1.53E-05 | |
| WHICAP (sensitivity) | Black | Cog. Impair. | 0.52 | 1.25E-03 | |
| WHICAP (sensitivity) | Black | Executive Function | | | 1.51E-05 |
| WHICAP (sensitivity) | Black | Memory | | | 3.74E-03 |
| WHICAP (sensitivity) | Black | Verbal Ability | | | 1.07E-03 |
| WHICAP (sensitivity) | Black | MMSE | | | 1.67E-03 |
| WHICAP (sensitivity) | Black | CDR | | | 9.08E-05 |
| WHICAP (sensitivity) | Black | Aβ42/Aβ40 | | | 1.59E-02 |
| WHICAP (sensitivity) | Black | Total Tau | | | 3.51E-02 |
| WHICAP (sensitivity) | Black | pTau | | | 8.94E-02 |
| WHICAP (sensitivity) | Black | NfL | | | 1.26E-01 |
| WHICAP (sensitivity) | Black | Cortical Thickness | | | 2.63E-02 |
| WHICAP (sensitivity) | Black | Hippo. Volume | | | 1.47E-01 |
| Demographics + APOE | Hispanic | MCI | 0.56 | 6.40E-03 | |
| Demographics + APOE | Hispanic | Dementia | 0.73 | 1.12E-01 | |
| Demographics + APOE | Hispanic | Cog. Impair. | 0.60 | 2.17E-02 | |
| Demographics + APOE | Hispanic | Executive Function | | | 2.09E-02 |
| Demographics + APOE | Hispanic | Memory | | | 5.35E-02 |
| Demographics + APOE | Hispanic | Verbal Ability | | | 1.70E-02 |
| Demographics + APOE | Hispanic | MMSE | | | 3.43E-01 |
| Demographics + APOE | Hispanic | CDR | | | 6.29E-02 |
| Demographics + APOE | Hispanic | Aβ42/Aβ40 | | | 4.12E-02 |
| Demographics + APOE | Hispanic | Total Tau | | | 1.55E-01 |
| Demographics + APOE | Hispanic | pTau | | | 2.01E-01 |
| Demographics + APOE | Hispanic | NfL | | | 2.65E-01 |
| Demographics + APOE | Hispanic | Cortical Thickness | | | 1.31E-01 |

| | | | | | |
|---|---|---|---|---|---|
| Demographics + APOE | Hispanic | Hippo. Volume | | | 3.33E-01 |
| CogDRisk | Hispanic | MCI | 0.56 | 6.93E-03 | |
| CogDRisk | Hispanic | Dementia | 0.63 | 4.22E-02 | |
| CogDRisk | Hispanic | Cog. Impair. | 0.58 | 1.54E-02 | |
| CogDRisk | Hispanic | Executive Function | | | 1.50E-02 |
| CogDRisk | Hispanic | Memory | | | 3.25E-03 |
| CogDRisk | Hispanic | Verbal Ability | | | 1.01E-02 |
| CogDRisk | Hispanic | MMSE | | | 2.53E-01 |
| CogDRisk | Hispanic | CDR | | | 3.65E-02 |
| CogDRisk | Hispanic | Aβ42/Aβ40 | | | 2.65E-02 |
| CogDRisk | Hispanic | Total Tau | | | 1.16E-01 |
| CogDRisk | Hispanic | pTau | | | 1.84E-01 |
| CogDRisk | Hispanic | NfL | | | 3.01E-01 |
| CogDRisk | Hispanic | Cortical Thickness | | | 1.03E-01 |
| CogDRisk | Hispanic | Hippo. Volume | | | 2.66E-01 |
| CogDRisk (sensitivity) | Hispanic | MCI | 0.54 | 3.28E-03 | |
| CogDRisk (sensitivity) | Hispanic | Dementia | 0.57 | 6.53E-03 | |
| CogDRisk (sensitivity) | Hispanic | Cog. Impair. | 0.55 | 4.45E-03 | |
| CogDRisk (sensitivity) | Hispanic | Executive Function | | | 1.02E-02 |
| CogDRisk (sensitivity) | Hispanic | Memory | | | 2.77E-04 |
| CogDRisk (sensitivity) | Hispanic | Verbal Ability | | | 8.06E-03 |
| CogDRisk (sensitivity) | Hispanic | MMSE | | | 1.64E-01 |
| CogDRisk (sensitivity) | Hispanic | CDR | | | 1.68E-02 |
| CogDRisk (sensitivity) | Hispanic | Aβ42/Aβ40 | | | 8.54E-03 |
| CogDRisk (sensitivity) | Hispanic | Total Tau | | | 1.12E-01 |
| CogDRisk (sensitivity) | Hispanic | pTau | | | 1.69E-01 |
| CogDRisk (sensitivity) | Hispanic | NfL | | | 2.06E-01 |
| CogDRisk (sensitivity) | Hispanic | Cortical Thickness | | | 6.35E-03 |
| CogDRisk (sensitivity) | Hispanic | Hippo. Volume | | | 1.49E-01 |
| LIBRA | Hispanic | MCI | 0.54 | 4.33E-03 | |
| LIBRA | Hispanic | Dementia | 0.61 | 1.98E-02 | |
| LIBRA | Hispanic | Cog. Impair. | 0.56 | 8.22E-03 | |
| LIBRA | Hispanic | Executive Function | | | 1.65E-02 |
| LIBRA | Hispanic | Memory | | | 7.09E-03 |
| LIBRA | Hispanic | Verbal Ability | | | 2.01E-02 |
| LIBRA | Hispanic | MMSE | | | 1.70E-01 |

| | | | | | |
|---|---|---|---|---|---|
| LIBRA | Hispanic | CDR | | | 2.02E-02 |
| LIBRA | Hispanic | Aβ42/Aβ40 | | | 1.92E-03 |
| LIBRA | Hispanic | Total Tau | | | 2.70E-02 |
| LIBRA | Hispanic | pTau | | | 2.12E-02 |
| LIBRA | Hispanic | NfL | | | 7.12E-02 |
| LIBRA | Hispanic | Cortical Thickness | | | 8.12E-03 |
| LIBRA | Hispanic | Hippo. Volume | | | 1.57E-01 |
| LIBRA (sensitivity) | Hispanic | MCI | 0.55 | 4.89E-03 | |
| LIBRA (sensitivity) | Hispanic | Dementia | 0.66 | 4.80E-02 | |
| LIBRA (sensitivity) | Hispanic | Cog. Impair. | 0.58 | 1.40E-02 | |
| LIBRA (sensitivity) | Hispanic | Executive Function | | | 2.30E-02 |
| LIBRA (sensitivity) | Hispanic | Memory | | | 6.97E-03 |
| LIBRA (sensitivity) | Hispanic | Verbal Ability | | | 2.49E-02 |
| LIBRA (sensitivity) | Hispanic | MMSE | | | 2.75E-01 |
| LIBRA (sensitivity) | Hispanic | CDR | | | 4.23E-02 |
| LIBRA (sensitivity) | Hispanic | Aβ42/Aβ40 | | | 1.21E-02 |
| LIBRA (sensitivity) | Hispanic | Total Tau | | | 2.89E-02 |
| LIBRA (sensitivity) | Hispanic | pTau | | | 5.71E-02 |
| LIBRA (sensitivity) | Hispanic | NfL | | | 1.84E-01 |
| LIBRA (sensitivity) | Hispanic | Cortical Thickness | | | 5.47E-02 |
| LIBRA (sensitivity) | Hispanic | Hippo. Volume | | | 2.08E-01 |
| mCAIDE | Hispanic | MCI | 0.53 | 1.82E-03 | |
| mCAIDE | Hispanic | Dementia | 0.60 | 1.71E-02 | |
| mCAIDE | Hispanic | Cog. Impair. | 0.55 | 4.99E-03 | |
| mCAIDE | Hispanic | Executive Function | | | 7.65E-03 |
| mCAIDE | Hispanic | Memory | | | 3.66E-03 |
| mCAIDE | Hispanic | Verbal Ability | | | 1.79E-02 |
| mCAIDE | Hispanic | MMSE | | | 1.87E-01 |
| mCAIDE | Hispanic | CDR | | | 1.22E-02 |
| mCAIDE | Hispanic | Aβ42/Aβ40 | | | 1.26E-02 |
| mCAIDE | Hispanic | Total Tau | | | 9.40E-02 |
| mCAIDE | Hispanic | pTau | | | 1.70E-01 |
| mCAIDE | Hispanic | NfL | | | 1.89E-01 |
| mCAIDE | Hispanic | Cortical Thickness | | | 4.45E-02 |
| mCAIDE | Hispanic | Hippo. Volume | | | 1.87E-01 |
| mCAIDE (sensitivity) | Hispanic | MCI | 0.50 | 4.89E-05 | |

| | | | | | |
|---|---|---|---|---|---|
| mCAIDE (sensitivity) | Hispanic | Dementia | 0.53 | 2.29E-03 | |
| mCAIDE (sensitivity) | Hispanic | Cog. Impair. | 0.51 | 3.78E-04 | |
| mCAIDE (sensitivity) | Hispanic | Executive Function | | | 6.46E-03 |
| mCAIDE (sensitivity) | Hispanic | Memory | | | 2.71E-04 |
| mCAIDE (sensitivity) | Hispanic | Verbal Ability | | | 1.47E-02 |
| mCAIDE (sensitivity) | Hispanic | MMSE | | | 1.60E-01 |
| mCAIDE (sensitivity) | Hispanic | CDR | | | 7.45E-03 |
| mCAIDE (sensitivity) | Hispanic | Aβ42/Aβ40 | | | 7.58E-03 |
| mCAIDE (sensitivity) | Hispanic | Total Tau | | | 9.94E-02 |
| mCAIDE (sensitivity) | Hispanic | pTau | | | 1.69E-01 |
| mCAIDE (sensitivity) | Hispanic | NfL | | | 1.75E-01 |
| mCAIDE (sensitivity) | Hispanic | Cortical Thickness | | | 1.05E-02 |
| mCAIDE (sensitivity) | Hispanic | Hippo. Volume | | | 1.48E-01 |
| WHICAP | Hispanic | MCI | 0.52 | 1.16E-03 | |
| WHICAP | Hispanic | Dementia | 0.65 | 4.48E-02 | |
| WHICAP | Hispanic | Cog. Impair. | 0.55 | 7.78E-03 | |
| WHICAP | Hispanic | Executive Function | | | 1.05E-02 |
| WHICAP | Hispanic | Memory | | | 5.53E-03 |
| WHICAP | Hispanic | Verbal Ability | | | 1.43E-02 |
| WHICAP | Hispanic | MMSE | | | 2.63E-01 |
| WHICAP | Hispanic | CDR | | | 2.96E-02 |
| WHICAP | Hispanic | Aβ42/Aβ40 | | | 1.55E-02 |
| WHICAP | Hispanic | Total Tau | | | 9.50E-02 |
| WHICAP | Hispanic | pTau | | | 1.74E-01 |
| WHICAP | Hispanic | NfL | | | 2.33E-01 |
| WHICAP | Hispanic | Cortical Thickness | | | 5.56E-02 |
| WHICAP | Hispanic | Hippo. Volume | | | 1.89E-01 |
| WHICAP (sensitivity) | Hispanic | MCI | 0.51 | 7.74E-05 | |
| WHICAP (sensitivity) | Hispanic | Dementia | 0.53 | 2.49E-04 | |
| WHICAP (sensitivity) | Hispanic | Cog. Impair. | 0.51 | 1.25E-04 | |
| WHICAP (sensitivity) | Hispanic | Executive Function | | | 5.79E-03 |
| WHICAP (sensitivity) | Hispanic | Memory | | | 3.14E-04 |
| WHICAP (sensitivity) | Hispanic | Verbal Ability | | | 1.08E-02 |
| WHICAP (sensitivity) | Hispanic | MMSE | | | 1.58E-01 |
| WHICAP (sensitivity) | Hispanic | CDR | | | 7.44E-03 |
| WHICAP (sensitivity) | Hispanic | Aβ42/Aβ40 | | | 2.55E-03 |

| | | | | | |
|---|---|---|---|---|---|
| WHICAP (sensitivity) | Hispanic | Total Tau | | | 9.92E-02 |
| WHICAP (sensitivity) | Hispanic | pTau | | | 1.69E-01 |
| WHICAP (sensitivity) | Hispanic | NfL | | | 1.76E-01 |
| WHICAP (sensitivity) | Hispanic | Cortical Thickness | | | 2.82E-03 |
| WHICAP (sensitivity) | Hispanic | Hippo. Volume | | | 1.43E-01 |
| Demographics + APOE | NHW | MCI | 0.63 | 3.15E-02 | |
| Demographics + APOE | NHW | Dementia | 0.75 | 1.01E-01 | |
| Demographics + APOE | NHW | Cog. Impair. | 0.66 | 4.78E-02 | |
| Demographics + APOE | NHW | Executive Function | | | 4.96E-02 |
| Demographics + APOE | NHW | Memory | | | 1.41E-01 |
| Demographics + APOE | NHW | Verbal Ability | | | 3.44E-02 |
| Demographics + APOE | NHW | MMSE | | | 8.31E-02 |
| Demographics + APOE | NHW | CDR | | | 3.71E-02 |
| Demographics + APOE | NHW | Aβ42/Aβ40 | | | 8.56E-02 |
| Demographics + APOE | NHW | Total Tau | | | 1.14E-01 |
| Demographics + APOE | NHW | pTau | | | 2.23E-01 |
| Demographics + APOE | NHW | NfL | | | 3.64E-01 |
| Demographics + APOE | NHW | Cortical Thickness | | | 1.91E-01 |
| Demographics + APOE | NHW | Hippo. Volume | | | 3.56E-01 |
| CogDRisk | NHW | MCI | 0.57 | 1.41E-02 | |
| CogDRisk | NHW | Dementia | 0.66 | 5.27E-02 | |
| CogDRisk | NHW | Cog. Impair. | 0.60 | 2.56E-02 | |
| CogDRisk | NHW | Executive Function | | | 4.73E-02 |
| CogDRisk | NHW | Memory | | | 3.59E-02 |
| CogDRisk | NHW | Verbal Ability | | | 2.49E-02 |
| CogDRisk | NHW | MMSE | | | 5.01E-02 |
| CogDRisk | NHW | CDR | | | 2.50E-02 |
| CogDRisk | NHW | Aβ42/Aβ40 | | | 2.94E-02 |
| CogDRisk | NHW | Total Tau | | | 6.09E-02 |
| CogDRisk | NHW | pTau | | | 1.46E-01 |
| CogDRisk | NHW | NfL | | | 3.59E-01 |
| CogDRisk | NHW | Cortical Thickness | | | 1.36E-01 |
| CogDRisk | NHW | Hippo. Volume | | | 3.07E-01 |
| CogDRisk (sensitivity) | NHW | MCI | 0.58 | 1.04E-02 | |
| CogDRisk (sensitivity) | NHW | Dementia | 0.59 | 1.63E-02 | |
| CogDRisk (sensitivity) | NHW | Cog. Impair. | 0.58 | 1.35E-02 | |

| | | | | | |
|---|---|---|---|---|---|
| CogDRisk (sensitivity) | NHW | Executive Function | | | 3.09E-03 |
| CogDRisk (sensitivity) | NHW | Memory | | | 4.07E-04 |
| CogDRisk (sensitivity) | NHW | Verbal Ability | | | 4.57E-03 |
| CogDRisk (sensitivity) | NHW | MMSE | | | 9.39E-03 |
| CogDRisk (sensitivity) | NHW | CDR | | | 1.07E-02 |
| CogDRisk (sensitivity) | NHW | Aβ42/Aβ40 | | | 1.49E-02 |
| CogDRisk (sensitivity) | NHW | Total Tau | | | 6.16E-02 |
| CogDRisk (sensitivity) | NHW | pTau | | | 8.19E-02 |
| CogDRisk (sensitivity) | NHW | NfL | | | 2.20E-01 |
| CogDRisk (sensitivity) | NHW | Cortical Thickness | | | 6.88E-04 |
| CogDRisk (sensitivity) | NHW | Hippo. Volume | | | 1.02E-01 |
| LIBRA | NHW | MCI | 0.59 | 1.49E-02 | |
| LIBRA | NHW | Dementia | 0.57 | 1.42E-02 | |
| LIBRA | NHW | Cog. Impair. | 0.58 | 1.62E-02 | |
| LIBRA | NHW | Executive Function | | | 6.07E-03 |
| LIBRA | NHW | Memory | | | 5.31E-03 |
| LIBRA | NHW | Verbal Ability | | | 9.96E-03 |
| LIBRA | NHW | MMSE | | | 1.06E-02 |
| LIBRA | NHW | CDR | | | 8.91E-03 |
| LIBRA | NHW | Aβ42/Aβ40 | | | 7.50E-03 |
| LIBRA | NHW | Total Tau | | | 1.43E-02 |
| LIBRA | NHW | pTau | | | 4.65E-04 |
| LIBRA | NHW | NfL | | | 6.28E-03 |
| LIBRA | NHW | Cortical Thickness | | | 2.39E-03 |
| LIBRA | NHW | Hippo. Volume | | | 1.06E-01 |
| LIBRA (sensitivity) | NHW | MCI | 0.59 | 1.77E-02 | |
| LIBRA (sensitivity) | NHW | Dementia | 0.63 | 4.01E-02 | |
| LIBRA (sensitivity) | NHW | Cog. Impair. | 0.60 | 2.59E-02 | |
| LIBRA (sensitivity) | NHW | Executive Function | | | 3.17E-02 |
| LIBRA (sensitivity) | NHW | Memory | | | 2.42E-02 |
| LIBRA (sensitivity) | NHW | Verbal Ability | | | 2.45E-02 |
| LIBRA (sensitivity) | NHW | MMSE | | | 3.34E-02 |
| LIBRA (sensitivity) | NHW | CDR | | | 2.05E-02 |
| LIBRA (sensitivity) | NHW | Aβ42/Aβ40 | | | 4.46E-04 |
| LIBRA (sensitivity) | NHW | Total Tau | | | 1.45E-02 |
| LIBRA (sensitivity) | NHW | pTau | | | 3.98E-02 |

| | | | | | |
|---|---|---|---|---|---|
| LIBRA (sensitivity) | NHW | NfL | | | 1.10E-01 |
| LIBRA (sensitivity) | NHW | Cortical Thickness | | | 5.96E-02 |
| LIBRA (sensitivity) | NHW | Hippo. Volume | | | 1.96E-01 |
| mCAIDE | NHW | MCI | 0.59 | 1.20E-02 | |
| mCAIDE | NHW | Dementia | 0.59 | 1.43E-02 | |
| mCAIDE | NHW | Cog. Impair. | 0.59 | 1.44E-02 | |
| mCAIDE | NHW | Executive Function | | | 1.05E-02 |
| mCAIDE | NHW | Memory | | | 2.60E-02 |
| mCAIDE | NHW | Verbal Ability | | | 1.57E-02 |
| mCAIDE | NHW | MMSE | | | 3.02E-02 |
| mCAIDE | NHW | CDR | | | 7.48E-03 |
| mCAIDE | NHW | Aβ42/Aβ40 | | | 6.86E-03 |
| mCAIDE | NHW | Total Tau | | | 4.54E-02 |
| mCAIDE | NHW | pTau | | | 8.48E-02 |
| mCAIDE | NHW | NfL | | | 2.23E-01 |
| mCAIDE | NHW | Cortical Thickness | | | 2.90E-02 |
| mCAIDE | NHW | Hippo. Volume | | | 1.45E-01 |
| mCAIDE (sensitivity) | NHW | MCI | 0.55 | 3.96E-03 | |
| mCAIDE (sensitivity) | NHW | Dementia | 0.54 | 2.87E-03 | |
| mCAIDE (sensitivity) | NHW | Cog. Impair. | 0.55 | 4.05E-03 | |
| mCAIDE (sensitivity) | NHW | Executive Function | | | 1.11E-03 |
| mCAIDE (sensitivity) | NHW | Memory | | | 2.59E-03 |
| mCAIDE (sensitivity) | NHW | Verbal Ability | | | 5.64E-03 |
| mCAIDE (sensitivity) | NHW | MMSE | | | 1.03E-02 |
| mCAIDE (sensitivity) | NHW | CDR | | | 1.61E-03 |
| mCAIDE (sensitivity) | NHW | Aβ42/Aβ40 | | | 1.07E-02 |
| mCAIDE (sensitivity) | NHW | Total Tau | | | 5.20E-02 |
| mCAIDE (sensitivity) | NHW | pTau | | | 8.29E-02 |
| mCAIDE (sensitivity) | NHW | NfL | | | 2.22E-01 |
| mCAIDE (sensitivity) | NHW | Cortical Thickness | | | 8.96E-04 |
| mCAIDE (sensitivity) | NHW | Hippo. Volume | | | 1.02E-01 |
| WHICAP | NHW | MCI | 0.57 | 1.15E-02 | |
| WHICAP | NHW | Dementia | 0.59 | 1.56E-02 | |
| WHICAP | NHW | Cog. Impair. | 0.57 | 1.39E-02 | |
| WHICAP | NHW | Executive Function | | | 2.79E-02 |
| WHICAP | NHW | Memory | | | 2.23E-02 |

| | | | | | |
|---|---|---|---|---|---|
| WHICAP | NHW | Verbal Ability | | | 2.66E-02 |
| WHICAP | NHW | MMSE | | | 3.23E-02 |
| WHICAP | NHW | CDR | | | 8.24E-03 |
| WHICAP | NHW | Aβ42/Aβ40 | | | 8.41E-03 |
| WHICAP | NHW | Total Tau | | | 4.53E-02 |
| WHICAP | NHW | pTau | | | 9.47E-02 |
| WHICAP | NHW | NfL | | | 2.51E-01 |
| WHICAP | NHW | Cortical Thickness | | | 9.27E-02 |
| WHICAP | NHW | Hippo. Volume | | | 2.36E-01 |
| WHICAP (sensitivity) | NHW | MCI | 0.54 | 2.24E-03 | |
| WHICAP (sensitivity) | NHW | Dementia | 0.52 | 1.45E-03 | |
| WHICAP (sensitivity) | NHW | Cog. Impair. | 0.52 | 4.36E-04 | |
| WHICAP (sensitivity) | NHW | Executive Function | | | 5.73E-04 |
| WHICAP (sensitivity) | NHW | Memory | | | 2.83E-04 |
| WHICAP (sensitivity) | NHW | Verbal Ability | | | 2.16E-03 |
| WHICAP (sensitivity) | NHW | MMSE | | | 3.06E-03 |
| WHICAP (sensitivity) | NHW | CDR | | | 1.10E-03 |
| WHICAP (sensitivity) | NHW | Aβ42/Aβ40 | | | 1.51E-02 |
| WHICAP (sensitivity) | NHW | Total Tau | | | 4.75E-02 |
| WHICAP (sensitivity) | NHW | pTau | | | 9.44E-02 |
| WHICAP (sensitivity) | NHW | NfL | | | 2.38E-01 |
| WHICAP (sensitivity) | NHW | Cortical Thickness | | | 2.06E-03 |
| WHICAP (sensitivity) | NHW | Hippo. Volume | | | 1.00E-01 |
| Demographics + APOE | all | MCI | 0.58 | 1.15E-02 | |
| Demographics + APOE | all | Dementia | 0.73 | 9.36E-02 | |
| Demographics + APOE | all | Cog. Impair. | 0.61 | 2.61E-02 | |
| Demographics + APOE | all | Executive Function | | | 2.17E-02 |
| Demographics + APOE | all | Memory | | | 7.69E-02 |
| Demographics + APOE | all | Verbal Ability | | | 2.38E-02 |
| Demographics + APOE | all | MMSE | | | 3.46E-01 |
| Demographics + APOE | all | CDR | | | 4.63E-02 |
| Demographics + APOE | all | Aβ42/Aβ40 | | | 4.63E-02 |
| Demographics + APOE | all | Total Tau | | | 1.12E-01 |
| Demographics + APOE | all | pTau | | | 2.14E-01 |
| Demographics + APOE | all | NfL | | | 2.93E-01 |
| Demographics + APOE | all | Cortical Thickness | | | 1.57E-01 |

| | | | | | |
|---|---|---|---|---|---|
| Demographics + APOE | all | Hippo. Volume | | | 3.24E-01 |
| CogDRisk | all | MCI | 0.53 | 2.40E-03 | |
| CogDRisk | all | Dementia | 0.65 | 4.58E-02 | |
| CogDRisk | all | Cog. Impair. | 0.56 | 9.54E-03 | |
| CogDRisk | all | Executive Function | | | 1.69E-02 |
| CogDRisk | all | Memory | | | 8.43E-03 |
| CogDRisk | all | Verbal Ability | | | 1.20E-02 |
| CogDRisk | all | MMSE | | | 2.39E-01 |
| CogDRisk | all | CDR | | | 2.54E-02 |
| CogDRisk | all | Aβ42/Aβ40 | | | 2.02E-02 |
| CogDRisk | all | Total Tau | | | 6.26E-02 |
| CogDRisk | all | pTau | | | 1.63E-01 |
| CogDRisk | all | NfL | | | 3.11E-01 |
| CogDRisk | all | Cortical Thickness | | | 1.28E-01 |
| CogDRisk | all | Hippo. Volume | | | 2.56E-01 |
| CogDRisk (sensitivity) | all | MCI | 0.55 | 5.54E-03 | |
| CogDRisk (sensitivity) | all | Dementia | 0.60 | 1.45E-02 | |
| CogDRisk (sensitivity) | all | Cog. Impair. | 0.56 | 8.22E-03 | |
| CogDRisk (sensitivity) | all | Executive Function | | | 6.46E-03 |
| CogDRisk (sensitivity) | all | Memory | | | 8.75E-04 |
| CogDRisk (sensitivity) | all | Verbal Ability | | | 5.07E-03 |
| CogDRisk (sensitivity) | all | MMSE | | | 1.98E-01 |
| CogDRisk (sensitivity) | all | CDR | | | 1.42E-02 |
| CogDRisk (sensitivity) | all | Aβ42/Aβ40 | | | 2.57E-03 |
| CogDRisk (sensitivity) | all | Total Tau | | | 6.27E-02 |
| CogDRisk (sensitivity) | all | pTau | | | 1.14E-01 |
| CogDRisk (sensitivity) | all | NfL | | | 1.59E-01 |
| CogDRisk (sensitivity) | all | Cortical Thickness | | | 4.07E-03 |
| CogDRisk (sensitivity) | all | Hippo. Volume | | | 1.11E-01 |
| LIBRA | all | MCI | 0.58 | 1.34E-02 | |
| LIBRA | all | Dementia | 0.62 | 2.74E-02 | |
| LIBRA | all | Cog. Impair. | 0.59 | 1.80E-02 | |
| LIBRA | all | Executive Function | | | 1.83E-02 |
| LIBRA | all | Memory | | | 1.21E-02 |
| LIBRA | all | Verbal Ability | | | 1.65E-02 |
| LIBRA | all | MMSE | | | 2.05E-01 |

| | | | | | |
|---|---|---|---|---|---|
| LIBRA | all | CDR | | | 1.93E-02 |
| LIBRA | all | Aβ42/Aβ40 | | | 2.52E-04 |
| LIBRA | all | Total Tau | | | 2.13E-02 |
| LIBRA | all | pTau | | | 2.38E-03 |
| LIBRA | all | NfL | | | 1.37E-02 |
| LIBRA | all | Cortical Thickness | | | 5.69E-03 |
| LIBRA | all | Hippo. Volume | | | 1.16E-01 |
| LIBRA (sensitivity) | all | MCI | 0.57 | 1.05E-02 | |
| LIBRA (sensitivity) | all | Dementia | 0.67 | 5.47E-02 | |
| LIBRA (sensitivity) | all | Cog. Impair. | 0.59 | 2.03E-02 | |
| LIBRA (sensitivity) | all | Executive Function | | | 2.99E-02 |
| LIBRA (sensitivity) | all | Memory | | | 1.73E-02 |
| LIBRA (sensitivity) | all | Verbal Ability | | | 2.46E-02 |
| LIBRA (sensitivity) | all | MMSE | | | 2.55E-01 |
| LIBRA (sensitivity) | all | CDR | | | 3.39E-02 |
| LIBRA (sensitivity) | all | Aβ42/Aβ40 | | | 7.15E-03 |
| LIBRA (sensitivity) | all | Total Tau | | | 2.24E-02 |
| LIBRA (sensitivity) | all | pTau | | | 4.23E-02 |
| LIBRA (sensitivity) | all | NfL | | | 1.11E-01 |
| LIBRA (sensitivity) | all | Cortical Thickness | | | 5.99E-02 |
| LIBRA (sensitivity) | all | Hippo. Volume | | | 1.87E-01 |
| mCAIDE | all | MCI | 0.56 | 6.89E-03 | |
| mCAIDE | all | Dementia | 0.61 | 2.01E-02 | |
| mCAIDE | all | Cog. Impair. | 0.57 | 1.08E-02 | |
| mCAIDE | all | Executive Function | | | 1.17E-02 |
| mCAIDE | all | Memory | | | 1.63E-02 |
| mCAIDE | all | Verbal Ability | | | 1.87E-02 |
| mCAIDE | all | MMSE | | | 2.16E-01 |
| mCAIDE | all | CDR | | | 9.27E-03 |
| mCAIDE | all | Aβ42/Aβ40 | | | 4.54E-03 |
| mCAIDE | all | Total Tau | | | 4.23E-02 |
| mCAIDE | all | pTau | | | 1.17E-01 |
| mCAIDE | all | NfL | | | 1.65E-01 |
| mCAIDE | all | Cortical Thickness | | | 3.67E-02 |
| mCAIDE | all | Hippo. Volume | | | 1.40E-01 |
| mCAIDE (sensitivity) | all | MCI | 0.54 | 3.17E-03 | |

| | | | | | |
|---|---|---|---|---|---|
| mCAIDE (sensitivity) | all | Dementia | 0.55 | 3.65E-03 | |
| mCAIDE (sensitivity) | all | Cog. Impair. | 0.54 | 3.63E-03 | |
| mCAIDE (sensitivity) | all | Executive Function | | | 5.10E-03 |
| mCAIDE (sensitivity) | all | Memory | | | 2.51E-03 |
| mCAIDE (sensitivity) | all | Verbal Ability | | | 9.87E-03 |
| mCAIDE (sensitivity) | all | MMSE | | | 1.95E-01 |
| mCAIDE (sensitivity) | all | CDR | | | 2.87E-03 |
| mCAIDE (sensitivity) | all | Aβ42/Aβ40 | | | 2.24E-03 |
| mCAIDE (sensitivity) | all | Total Tau | | | 4.89E-02 |
| mCAIDE (sensitivity) | all | pTau | | | 1.15E-01 |
| mCAIDE (sensitivity) | all | NfL | | | 1.55E-01 |
| mCAIDE (sensitivity) | all | Cortical Thickness | | | 5.41E-03 |
| mCAIDE (sensitivity) | all | Hippo. Volume | | | 1.10E-01 |
| WHICAP | all | MCI | 0.57 | 1.09E-02 | |
| WHICAP | all | Dementia | 0.63 | 3.64E-02 | |
| WHICAP | all | Cog. Impair. | 0.59 | 1.77E-02 | |
| WHICAP | all | Executive Function | | | 3.51E-02 |
| WHICAP | all | Memory | | | 2.50E-02 |
| WHICAP | all | Verbal Ability | | | 3.19E-02 |
| WHICAP | all | MMSE | | | 2.63E-01 |
| WHICAP | all | CDR | | | 2.42E-02 |
| WHICAP | all | Aβ42/Aβ40 | | | 1.16E-02 |
| WHICAP | all | Total Tau | | | 4.73E-02 |
| WHICAP | all | pTau | | | 1.15E-01 |
| WHICAP | all | NfL | | | 1.81E-01 |
| WHICAP | all | Cortical Thickness | | | 4.51E-02 |
| WHICAP | all | Hippo. Volume | | | 1.47E-01 |
| WHICAP (sensitivity) | all | MCI | 0.60 | 1.86E-02 | |
| WHICAP (sensitivity) | all | Dementia | 0.55 | 3.19E-03 | |
| WHICAP (sensitivity) | all | Cog. Impair. | 0.59 | 1.41E-02 | |
| WHICAP (sensitivity) | all | Executive Function | | | 1.84E-02 |
| WHICAP (sensitivity) | all | Memory | | | 1.23E-02 |
| WHICAP (sensitivity) | all | Verbal Ability | | | 1.48E-02 |
| WHICAP (sensitivity) | all | MMSE | | | 2.00E-01 |
| WHICAP (sensitivity) | all | CDR | | | 4.89E-03 |
| WHICAP (sensitivity) | all | Aβ42/Aβ40 | | | 2.24E-03 |

| | | | | | |
|---|---|---|---|---|---|
| WHICAP (sensitivity) | all | Total Tau | | | 4.67E-02 |
| WHICAP (sensitivity) | all | pTau | | | 1.31E-01 |
| WHICAP (sensitivity) | all | NfL | | | 1.68E-01 |
| WHICAP (sensitivity) | all | Cortical Thickness | | | 4.30E-03 |
| WHICAP (sensitivity) | all | Hippo. Volume | | | 1.07E-01 |

Table S4. Z-test results comparing regression coefficients (β) between two racial groups (pop1 vs. pop2).

| Model | Outcome | Pop1 | Pop2 | Z-score | P-Value | P-Value FDR. adj. | Significance |
|---|---|---|---|---|---|---|---|
| CogDrisk | Aβ2/Aβ0 | AA | LA | 1.54 | 1.25E-01 | 4.58E-01 | |
| CogDrisk | Aβ2/Aβ0 | AA | NHW | 0.89 | 3.74E-01 | 7.37E-01 | |
| CogDrisk | Aβ2/Aβ0 | LA | NHW | -1.12 | 2.64E-01 | 6.59E-01 | |
| CogDrisk | Cortical Thickness | AA | LA | -4.00 | 6.37E-05 | 1.64E-03 | ** |
| CogDrisk | Cortical Thickness | AA | NHW | 1.00 | 3.15E-01 | 7.37E-01 | |
| CogDrisk | Cortical Thickness | LA | NHW | 7.44 | 9.73E-14 | 5.84E-12 | *** |
| CogDrisk | Healthy | AA | LA | 0.33 | 7.44E-01 | 9.36E-01 | |
| CogDrisk | Healthy | AA | NHW | 0.94 | 3.46E-01 | 7.37E-01 | |
| CogDrisk | Healthy | LA | NHW | 0.82 | 4.10E-01 | 7.70E-01 | |
| CogDrisk | MMSE | AA | LA | -0.13 | 8.93E-01 | 9.82E-01 | |
| CogDrisk | MMSE | AA | NHW | -0.72 | 4.71E-01 | 8.07E-01 | |
| CogDrisk | MMSE | LA | NHW | -0.68 | 4.94E-01 | 8.15E-01 | |
| CogDrisk | pTau | AA | LA | 0.21 | 8.35E-01 | 9.51E-01 | |
| CogDrisk | pTau | AA | NHW | -1.74 | 8.18E-02 | 3.78E-01 | |
| CogDrisk | pTau | LA | NHW | -2.91 | 3.60E-03 | 4.05E-02 | * |
| CogDrisk | Executive Function | AA | LA | -2.15 | 3.15E-02 | 1.58E-01 | |
| CogDrisk | Executive Function | AA | NHW | -0.79 | 4.28E-01 | 7.79E-01 | |
| CogDrisk | Executive Function | LA | NHW | 2.26 | 2.40E-02 | 1.31E-01 | |
| CogDrisk | Verbal Ability | AA | LA | -2.98 | 2.84E-03 | 3.41E-02 | * |
| CogDrisk | Verbal Ability | AA | NHW | -1.69 | 9.11E-02 | 3.90E-01 | |
| CogDrisk | Verbal Ability | LA | NHW | 2.00 | 4.55E-02 | 2.21E-01 | |
| CogDrisk | Hippo. Volume | AA | LA | 2.26 | 2.38E-02 | 1.31E-01 | |
| CogDrisk | Hippo. Volume | AA | NHW | 3.96 | 7.53E-05 | 1.69E-03 | ** |
| CogDrisk | Hippo. Volume | LA | NHW | 2.33 | 1.99E-02 | 1.19E-01 | |
| CogDrisk | NfL | AA | LA | -1.55 | 1.22E-01 | 4.56E-01 | |
| CogDrisk | NfL | AA | NHW | -1.56 | 1.20E-01 | 4.56E-01 | |
| CogDrisk | NfL | LA | NHW | 0.06 | 9.50E-01 | 9.84E-01 | |
| CogDrisk | Memory | AA | LA | -2.36 | 1.83E-02 | 1.19E-01 | |
| CogDrisk | Memory | AA | NHW | 0.24 | 8.12E-01 | 9.48E-01 | |
| CogDrisk | Memory | LA | NHW | 3.83 | 1.30E-04 | 2.61E-03 | ** |
| CogDrisk | Total Tau | AA | LA | 0.66 | 5.07E-01 | 8.15E-01 | |
| CogDrisk | Total Tau | AA | NHW | 1.69 | 9.16E-02 | 3.90E-01 | |

| | | | | | | | |
|---|---|---|---|---|---|---|---|
| CogDrisk | Total Tau | LA | NHW | 1.58 | 1.15E-01 | 4.50E-01 | |
| CogDrisk | CDR | AA | LA | 0.57 | 5.71E-01 | 8.94E-01 | |
| CogDrisk | CDR | AA | NHW | 0.78 | 4.35E-01 | 7.82E-01 | |
| CogDrisk | CDR | LA | NHW | 0.31 | 7.54E-01 | 9.36E-01 | |
| CogDrisk | MCI | AA | LA | -0.50 | 6.16E-01 | 9.34E-01 | |
| CogDrisk | MCI | AA | NHW | -1.02 | 3.06E-01 | 7.35E-01 | |
| CogDrisk | MCI | LA | NHW | -0.68 | 4.98E-01 | 8.15E-01 | |
| CogDrisk | Dementia | AA | LA | 0.81 | 4.20E-01 | 7.71E-01 | |
| CogDrisk | Dementia | AA | NHW | 0.26 | 7.92E-01 | 9.44E-01 | |
| CogDrisk | Dementia | LA | NHW | -0.70 | 4.84E-01 | 8.15E-01 | |
| CogDrisk | Cog. Impair. | AA | LA | -0.33 | 7.44E-01 | 9.36E-01 | |
| CogDrisk | Cog. Impair. | AA | NHW | -0.94 | 3.46E-01 | 7.37E-01 | |
| CogDrisk | Cog. Impair. | LA | NHW | -0.82 | 4.10E-01 | 7.70E-01 | |
| LIBRA | Aβ2/Aβ0 | AA | LA | -1.43 | 1.52E-01 | 5.02E-01 | |
| LIBRA | Aβ2/Aβ0 | AA | NHW | -3.01 | 2.65E-03 | 3.41E-02 | * |
| LIBRA | Aβ2/Aβ0 | LA | NHW | -2.41 | 1.57E-02 | 1.19E-01 | |
| LIBRA | Cortical Thickness | AA | LA | 1.59 | 1.12E-01 | 4.47E-01 | |
| LIBRA | Cortical Thickness | AA | NHW | -0.24 | 8.07E-01 | 9.48E-01 | |
| LIBRA | Cortical Thickness | LA | NHW | -2.54 | 1.12E-02 | 9.61E-02 | |
| LIBRA | Healthy | AA | LA | -0.39 | 7.00E-01 | 9.36E-01 | |
| LIBRA | Healthy | AA | NHW | 0.34 | 7.34E-01 | 9.36E-01 | |
| LIBRA | Healthy | LA | NHW | 0.89 | 3.73E-01 | 7.37E-01 | |
| LIBRA | MMSE | AA | LA | -0.11 | 9.13E-01 | 9.82E-01 | |
| LIBRA | MMSE | AA | NHW | -0.21 | 8.35E-01 | 9.51E-01 | |
| LIBRA | MMSE | LA | NHW | -0.08 | 9.33E-01 | 9.82E-01 | |
| LIBRA | pTau | AA | LA | -0.37 | 7.13E-01 | 9.36E-01 | |
| LIBRA | pTau | AA | NHW | 1.21 | 2.25E-01 | 6.23E-01 | |
| LIBRA | pTau | LA | NHW | 2.21 | 2.69E-02 | 1.38E-01 | |
| LIBRA | Executive Function | AA | LA | -0.90 | 3.69E-01 | 7.37E-01 | |
| LIBRA | Executive Function | AA | NHW | -1.61 | 1.08E-01 | 4.41E-01 | |
| LIBRA | Executive Function | LA | NHW | -1.01 | 3.13E-01 | 7.37E-01 | |
| LIBRA | Verbal Ability | AA | LA | -0.88 | 3.76E-01 | 7.37E-01 | |
| LIBRA | Verbal Ability | AA | NHW | -1.40 | 1.63E-01 | 5.14E-01 | |
| LIBRA | Verbal Ability | LA | NHW | -0.68 | 4.94E-01 | 8.15E-01 | |
| LIBRA | Hippo. Volume | AA | LA | 0.95 | 3.43E-01 | 7.37E-01 | |

| | | | | | | | |
|---|---|---|---|---|---|---|---|
| LIBRA | Hippo. Volume | AA | NHW | -0.09 | 9.29E-01 | 9.82E-01 | |
| LIBRA | Hippo. Volume | LA | NHW | -1.28 | 2.00E-01 | 6.11E-01 | |
| LIBRA | NfL | AA | LA | -2.54 | 1.12E-02 | 9.61E-02 | |
| LIBRA | NfL | AA | NHW | 0.34 | 7.37E-01 | 9.36E-01 | |
| LIBRA | NfL | LA | NHW | 3.50 | 4.69E-04 | 7.24E-03 | ** |
| LIBRA | Memory | AA | LA | -0.26 | 7.97E-01 | 9.44E-01 | |
| LIBRA | Memory | AA | NHW | 0.08 | 9.33E-01 | 9.82E-01 | |
| LIBRA | Memory | LA | NHW | 0.47 | 6.37E-01 | 9.35E-01 | |
| LIBRA | Total Tau | AA | LA | 0.11 | 9.09E-01 | 9.82E-01 | |
| LIBRA | Total Tau | AA | NHW | 0.75 | 4.54E-01 | 7.99E-01 | |
| LIBRA | Total Tau | LA | NHW | 0.92 | 3.60E-01 | 7.37E-01 | |
| LIBRA | CDR | AA | LA | 0.41 | 6.78E-01 | 9.36E-01 | |
| LIBRA | CDR | AA | NHW | 0.62 | 5.34E-01 | 8.51E-01 | |
| LIBRA | CDR | LA | NHW | 0.29 | 7.75E-01 | 9.36E-01 | |
| LIBRA | MCI | AA | LA | 0.51 | 6.13E-01 | 9.34E-01 | |
| LIBRA | MCI | AA | NHW | -0.44 | 6.57E-01 | 9.35E-01 | |
| LIBRA | MCI | LA | NHW | -1.12 | 2.65E-01 | 6.59E-01 | |
| LIBRA | Dementia | AA | LA | 0.21 | 8.33E-01 | 9.51E-01 | |
| LIBRA | Dementia | AA | NHW | 0.28 | 7.80E-01 | 9.36E-01 | |
| LIBRA | Dementia | LA | NHW | 0.09 | 9.26E-01 | 9.82E-01 | |
| LIBRA | Cog. Impair. | AA | LA | 0.39 | 7.00E-01 | 9.36E-01 | |
| LIBRA | Cog. Impair. | AA | NHW | -0.34 | 7.34E-01 | 9.36E-01 | |
| LIBRA | Cog. Impair. | LA | NHW | -0.89 | 3.73E-01 | 7.37E-01 | |
| mCAIDE | Aβ2/Aβ0 | AA | LA | 1.46 | 1.45E-01 | 5.02E-01 | |
| mCAIDE | Aβ2/Aβ0 | AA | NHW | -0.04 | 9.64E-01 | 9.92E-01 | |
| mCAIDE | Aβ2/Aβ0 | LA | NHW | -2.37 | 1.76E-02 | 1.19E-01 | |
| mCAIDE | Cortical Thickness | AA | LA | 6.12 | 9.23E-10 | 4.15E-08 | *** |
| mCAIDE | Cortical Thickness | AA | NHW | 3.10 | 1.95E-03 | 2.70E-02 | * |
| mCAIDE | Cortical Thickness | LA | NHW | -4.02 | 5.79E-05 | 1.64E-03 | ** |
| mCAIDE | Healthy | AA | LA | -1.43 | 1.54E-01 | 5.02E-01 | |
| mCAIDE | Healthy | AA | NHW | -0.41 | 6.83E-01 | 9.36E-01 | |
| mCAIDE | Healthy | LA | NHW | 1.23 | 2.19E-01 | 6.23E-01 | |
| mCAIDE | MMSE | AA | LA | -0.08 | 9.40E-01 | 9.83E-01 | |
| mCAIDE | MMSE | AA | NHW | -0.11 | 9.13E-01 | 9.82E-01 | |
| mCAIDE | MMSE | LA | NHW | -0.02 | 9.84E-01 | 9.92E-01 | |
| mCAIDE | pTau | AA | LA | 0.72 | 4.71E-01 | 8.07E-01 | |

| | | | | | | | |
|---|---|---|---|---|---|---|---|
| mCAIDE | pTau | AA | NHW | 0.29 | 7.70E-01 | 9.36E-01 | |
| mCAIDE | pTau | LA | NHW | -0.60 | 5.48E-01 | 8.65E-01 | |
| mCAIDE | Executive Function | AA | LA | -2.28 | 2.25E-02 | 1.30E-01 | |
| mCAIDE | Executive Function | AA | NHW | -1.68 | 9.31E-02 | 3.90E-01 | |
| mCAIDE | Executive Function | LA | NHW | 0.97 | 3.30E-01 | 7.37E-01 | |
| mCAIDE | Verbal Ability | AA | LA | -2.35 | 1.86E-02 | 1.19E-01 | |
| mCAIDE | Verbal Ability | AA | NHW | -2.21 | 2.69E-02 | 1.38E-01 | |
| mCAIDE | Verbal Ability | LA | NHW | 0.23 | 8.16E-01 | 9.48E-01 | |
| mCAIDE | Hippo. Volume | AA | LA | 0.86 | 3.87E-01 | 7.50E-01 | |
| mCAIDE | Hippo. Volume | AA | NHW | 0.84 | 4.00E-01 | 7.66E-01 | |
| mCAIDE | Hippo. Volume | LA | NHW | 0.00 | 9.97E-01 | 9.97E-01 | |
| mCAIDE | NfL | AA | LA | -0.74 | 4.57E-01 | 7.99E-01 | |
| mCAIDE | NfL | AA | NHW | 0.49 | 6.28E-01 | 9.35E-01 | |
| mCAIDE | NfL | LA | NHW | 1.52 | 1.27E-01 | 4.59E-01 | |
| mCAIDE | Memory | AA | LA | -2.36 | 1.84E-02 | 1.19E-01 | |
| mCAIDE | Memory | AA | NHW | -0.33 | 7.45E-01 | 9.36E-01 | |
| mCAIDE | Memory | LA | NHW | 2.83 | 4.67E-03 | 4.95E-02 | * |
| mCAIDE | Total Tau | AA | LA | 0.02 | 9.86E-01 | 9.92E-01 | |
| mCAIDE | Total Tau | AA | NHW | 0.33 | 7.43E-01 | 9.36E-01 | |
| mCAIDE | Total Tau | LA | NHW | 0.46 | 6.45E-01 | 9.35E-01 | |
| mCAIDE | CDR | AA | LA | 0.67 | 5.00E-01 | 8.15E-01 | |
| mCAIDE | CDR | AA | NHW | 0.40 | 6.87E-01 | 9.36E-01 | |
| mCAIDE | CDR | LA | NHW | -0.52 | 6.00E-01 | 9.31E-01 | |
| mCAIDE | MCI | AA | LA | 1.40 | 1.60E-01 | 5.14E-01 | |
| mCAIDE | MCI | AA | NHW | 0.13 | 8.97E-01 | 9.82E-01 | |
| mCAIDE | MCI | LA | NHW | -1.46 | 1.45E-01 | 5.02E-01 | |
| mCAIDE | Dementia | AA | LA | 1.25 | 2.11E-01 | 6.23E-01 | |
| mCAIDE | Dementia | AA | NHW | 1.13 | 2.57E-01 | 6.59E-01 | |
| mCAIDE | Dementia | LA | NHW | -0.10 | 9.20E-01 | 9.82E-01 | |
| mCAIDE | Cog. Impair. | AA | LA | 1.43 | 1.54E-01 | 5.02E-01 | |
| mCAIDE | Cog. Impair. | AA | NHW | 0.41 | 6.83E-01 | 9.36E-01 | |
| mCAIDE | Cog. Impair. | LA | NHW | -1.23 | 2.19E-01 | 6.23E-01 | |
| WHICAP | Aβ2/Aβ0 | AA | LA | 0.20 | 8.41E-01 | 9.52E-01 | |
| WHICAP | Aβ2/Aβ0 | AA | NHW | -0.29 | 7.74E-01 | 9.36E-01 | |
| WHICAP | Aβ2/Aβ0 | LA | NHW | -0.96 | 3.35E-01 | 7.37E-01 | |

| | | | | | | | |
|---|---|---|---|---|---|---|---|
| WHICAP | Cortical Thickness | AA | LA | 4.66 | 3.16E-06 | 1.14E-04 | *** |
| WHICAP | Cortical Thickness | AA | NHW | 11.61 | 0.00E+00 | 0.00E+00 | *** |
| WHICAP | Cortical Thickness | LA | NHW | 11.93 | 0.00E+00 | 0.00E+00 | *** |
| WHICAP | Healthy | AA | LA | -0.29 | 7.74E-01 | 9.36E-01 | |
| WHICAP | Healthy | AA | NHW | 0.019 | 9.85E-01 | 9.92E-01 | |
| WHICAP | Healthy | LA | NHW | 0.44 | 6.58E-01 | 9.35E-01 | |
| WHICAP | MMSE | AA | LA | 0.14 | 8.91E-01 | 9.82E-01 | |
| WHICAP | MMSE | AA | NHW | -0.50 | 6.17E-01 | 9.34E-01 | |
| WHICAP | MMSE | LA | NHW | -0.92 | 3.56E-01 | 7.37E-01 | |
| WHICAP | pTau | AA | LA | -0.09 | 9.30E-01 | 9.82E-01 | |
| WHICAP | pTau | AA | NHW | -0.89 | 3.72E-01 | 7.37E-01 | |
| WHICAP | pTau | LA | NHW | -1.38 | 1.67E-01 | 5.19E-01 | |
| WHICAP | Executive Function | AA | LA | -2.56 | 1.05E-02 | 9.61E-02 | |
| WHICAP | Executive Function | AA | NHW | -1.22 | 2.23E-01 | 6.23E-01 | |
| WHICAP | Executive Function | LA | NHW | 2.35 | 1.86E-02 | 1.19E-01 | |
| WHICAP | Verbal Ability | AA | LA | -3.57 | 3.55E-04 | 6.39E-03 | ** |
| WHICAP | Verbal Ability | AA | NHW | -2.33 | 1.99E-02 | 1.19E-01 | |
| WHICAP | Verbal Ability | LA | NHW | 1.95 | 5.06E-02 | 2.40E-01 | |
| WHICAP | Hippo. Volume | AA | LA | 0.06 | 9.51E-01 | 9.84E-01 | |
| WHICAP | Hippo. Volume | AA | NHW | 2.39 | 1.67E-02 | 1.19E-01 | |
| WHICAP | Hippo. Volume | LA | NHW | 3.49 | 4.82E-04 | 7.24E-03 | ** |
| WHICAP | NfL | AA | LA | -1.69 | 9.19E-02 | 3.90E-01 | |
| WHICAP | NfL | AA | NHW | -0.99 | 3.22E-01 | 7.37E-01 | |
| WHICAP | NfL | LA | NHW | 0.98 | 3.28E-01 | 7.37E-01 | |
| WHICAP | Memory | AA | LA | -1.19 | 2.34E-01 | 6.38E-01 | |
| WHICAP | Memory | AA | NHW | 0.44 | 6.59E-01 | 9.35E-01 | |
| WHICAP | Memory | LA | NHW | 2.59 | 9.67E-03 | 9.61E-02 | |
| WHICAP | Total Tau | AA | LA | 0.76 | 4.49E-01 | 7.99E-01 | |
| WHICAP | Total Tau | AA | NHW | 1.17 | 2.40E-01 | 6.45E-01 | |
| WHICAP | Total Tau | LA | NHW | 0.81 | 4.17E-01 | 7.71E-01 | |
| WHICAP | CDR | AA | LA | 0.66 | 5.07E-01 | 8.15E-01 | |
| WHICAP | CDR | AA | NHW | 1.26 | 2.08E-01 | 6.23E-01 | |
| WHICAP | CDR | LA | NHW | 1.14 | 2.55E-01 | 6.59E-01 | |
| WHICAP | MCI | AA | LA | 0.28 | 7.77E-01 | 9.36E-01 | |
| WHICAP | MCI | AA | NHW | -0.48 | 6.34E-01 | 9.35E-01 | |

| | | | | | | |
|---|---|---|---|---|---|---|
| WHICAP | MCI | LA | NHW | -1.13 | 2.59E-01 | 6.59E-01 |
| WHICAP | Dementia | AA | LA | 0.41 | 6.80E-01 | 9.36E-01 |
| WHICAP | Dementia | AA | NHW | 1.11 | 2.67E-01 | 6.59E-01 |
| WHICAP | Dementia | LA | NHW | 1.06 | 2.88E-01 | 7.00E-01 |
| WHICAP | Cog. Impair. | AA | LA | 0.29 | 7.74E-01 | 9.36E-01 |
| WHICAP | Cog. Impair. | AA | NHW | -0.019 | 9.85E-01 | 9.92E-01 |
| WHICAP | Cog. Impair. | LA | NHW | -0.44 | 6.58E-01 | 9.35E-01 |

Table S5. Regression model estimates for original CRS and CRS sensitivity analysis for mCAIDE, WHICAP, and CogDRisk for complete cases. For the sensitivity analysis, age, sex, and years of education covariates were excluded from the scoring.

| Race | CRS | Outcome | P-value | P-value, FDR adj. | β | Lower Conf. Interval | Upper Conf. Interval |
|---|---|---|---|---|---|---|---|
| Black | CogDRisk | Cortical Thickness | 1.28E-08 | 5.32E-08 | -3.69E-01 | -4.92E-01 | -2.46E-01 |
| Black | CogDRisk | MMSE | 1.29E-07 | 4.91E-07 | -3.27E-01 | -4.47E-01 | -2.08E-01 |
| Black | CogDRisk | Executive Function | 1.77E-05 | 4.49E-05 | -2.67E-01 | -3.88E-01 | -1.46E-01 |
| Black | CogDRisk | Verbal Ability | 8.80E-05 | 2.01E-04 | -2.44E-01 | -3.65E-01 | -1.23E-01 |
| Black | CogDRisk | Hippocampal Volume | 2.55E-06 | 7.86E-06 | -2.35E-01 | -3.31E-01 | -1.39E-01 |
| Black | CogDRisk | Memory | 2.60E-03 | 4.30E-03 | -1.90E-01 | -3.14E-01 | -6.69E-02 |
| Black | CogDRisk | Aβ42/Aβ40 | 6.57E-01 | 6.71E-01 | -2.65E-02 | -1.44E-01 | 9.06E-02 |
| Black | CogDRisk | Total Tau | 8.74E-03 | 1.28E-02 | 1.48E-01 | 3.75E-02 | 2.58E-01 |
| Black | CogDRisk | MCI | 1.77E-01 | 1.93E-01 | 1.55E-01 | -6.96E-02 | 3.82E-01 |
| Black | CogDRisk | pTau | 2.57E-03 | 4.28E-03 | 1.62E-01 | 5.71E-02 | 2.67E-01 |
| Black | CogDRisk | CDR | 6.29E-04 | 1.21E-03 | 2.15E-01 | 9.24E-02 | 3.37E-01 |
| Black | CogDRisk | NfL | 3.31E-06 | 9.77E-06 | 2.54E-01 | 1.48E-01 | 3.59E-01 |
| Black | CogDRisk | Cog. Impair. | 1.24E-02 | 1.70E-02 | 2.81E-01 | 6.37E-02 | 5.05E-01 |
| Black | CogDRisk | Dementia | 1.08E-04 | 2.39E-04 | 7.97E-01 | 4.08E-01 | 1.22E+00 |
| Black | LIBRA | Executive Function | 3.26E-03 | 5.31E-03 | -1.70E-01 | -2.83E-01 | -5.73E-02 |
| Black | LIBRA | Aβ42/Aβ40 | 2.90E-03 | 4.78E-03 | -1.69E-01 | -2.80E-01 | -5.84E-02 |
| Black | LIBRA | Verbal Ability | 3.65E-03 | 5.91E-03 | -1.68E-01 | -2.81E-01 | -5.52E-02 |
| Black | LIBRA | MMSE | 7.12E-02 | 8.53E-02 | -1.05E-01 | -2.20E-01 | 9.15E-03 |
| Black | LIBRA | Hippocampal Volume | 5.77E-02 | 7.01E-02 | -9.50E-02 | -1.93E-01 | 3.14E-03 |
| Black | LIBRA | Memory | 1.65E-01 | 1.83E-01 | -8.15E-02 | -1.97E-01 | 3.36E-02 |
| Black | LIBRA | Cortical Thickness | 2.40E-01 | 2.54E-01 | -7.87E-02 | -2.11E-01 | 5.30E-02 |
| Black | LIBRA | NfL | 4.51E-02 | 5.66E-02 | 1.12E-01 | 2.44E-03 | 2.21E-01 |
| Black | LIBRA | pTau | 1.17E-02 | 1.64E-02 | 1.38E-01 | 3.08E-02 | 2.44E-01 |
| Black | LIBRA | CDR | 5.07E-03 | 7.69E-03 | 1.63E-01 | 4.94E-02 | 2.77E-01 |
| Black | LIBRA | Total Tau | 5.85E-04 | 1.13E-03 | 1.90E-01 | 8.22E-02 | 2.97E-01 |
| Black | LIBRA | MCI | 2.76E-02 | 3.54E-02 | 2.57E-01 | 3.04E-02 | 4.88E-01 |
| Black | LIBRA | Cog. Impair. | 9.46E-03 | 1.36E-02 | 2.89E-01 | 7.29E-02 | 5.10E-01 |
| Black | LIBRA | Dementia | 4.35E-02 | 5.47E-02 | 4.20E-01 | 1.94E-02 | 8.41E-01 |
| Black | mCAIDE | Verbal Ability | 4.15E-05 | 1.02E-04 | -2.32E-01 | -3.43E-01 | -1.22E-01 |
| Black | mCAIDE | Executive Function | 1.18E-03 | 2.15E-03 | -1.85E-01 | -2.96E-01 | -7.36E-02 |
| Black | mCAIDE | Memory | 2.36E-03 | 3.97E-03 | -1.75E-01 | -2.87E-01 | -6.26E-02 |
| Black | mCAIDE | Hippocampal Volume | 4.93E-04 | 9.65E-04 | -1.71E-01 | -2.66E-01 | -7.53E-02 |

| | | | | | | | |
|---|---|---|---|---|---|---|---|
| Black | mCAIDE | MMSE | 4.02E-03 | 6.40E-03 | -1.65E-01 | -2.77E-01 | -5.28E-02 |
| Black | mCAIDE | Cortical Thickness | 3.49E-02 | 4.42E-02 | -1.42E-01 | -2.73E-01 | -1.01E-02 |
| Black | mCAIDE | Aβ42/Aβ40 | 9.21E-01 | 9.25E-01 | -5.65E-03 | -1.18E-01 | 1.07E-01 |
| Black | mCAIDE | Total Tau | 3.63E-01 | 3.75E-01 | 4.99E-02 | -5.78E-02 | 1.58E-01 |
| Black | mCAIDE | NfL | 1.74E-01 | 1.91E-01 | 7.33E-02 | -3.25E-02 | 1.79E-01 |
| Black | mCAIDE | pTau | 1.34E-01 | 1.52E-01 | 7.79E-02 | -2.41E-02 | 1.80E-01 |
| Black | mCAIDE | CDR | 3.49E-02 | 4.42E-02 | 1.21E-01 | 8.69E-03 | 2.34E-01 |
| Black | mCAIDE | MCI | 1.20E-02 | 1.67E-02 | 2.88E-01 | 6.54E-02 | 5.15E-01 |
| Black | mCAIDE | Cog. Impair. | 1.74E-03 | 3.04E-03 | 3.44E-01 | 1.31E-01 | 5.63E-01 |
| Black | mCAIDE | Dementia | 3.67E-03 | 5.91E-03 | 6.57E-01 | 2.29E-01 | 1.12E+00 |
| Black | WHICAP | Verbal Ability | 1.20E-05 | 3.10E-05 | -3.49E-01 | -5.03E-01 | -1.94E-01 |
| Black | WHICAP | MMSE | 1.16E-04 | 2.54E-04 | -3.10E-01 | -4.66E-01 | -1.54E-01 |
| Black | WHICAP | Executive Function | 5.09E-04 | 9.89E-04 | -2.79E-01 | -4.35E-01 | -1.23E-01 |
| Black | WHICAP | Hippocampal Volume | 2.94E-05 | 7.32E-05 | -2.07E-01 | -3.03E-01 | -1.11E-01 |
| Black | WHICAP | Cortical Thickness | 1.04E-02 | 1.48E-02 | -1.71E-01 | -3.01E-01 | -4.05E-02 |
| Black | WHICAP | Memory | 4.59E-02 | 5.71E-02 | -1.62E-01 | -3.22E-01 | -2.99E-03 |
| Black | WHICAP | Aβ42/Aβ40 | 1.10E-01 | 1.28E-01 | -9.35E-02 | -2.09E-01 | 2.14E-02 |
| Black | WHICAP | pTau | 1.48E-01 | 1.67E-01 | 7.72E-02 | -2.75E-02 | 1.82E-01 |
| Black | WHICAP | Total Tau | 3.04E-02 | 3.88E-02 | 1.22E-01 | 1.16E-02 | 2.32E-01 |
| Black | WHICAP | NfL | 2.17E-02 | 2.84E-02 | 1.26E-01 | 1.85E-02 | 2.33E-01 |
| Black | WHICAP | MCI | 1.14E-01 | 1.31E-01 | 1.83E-01 | -4.26E-02 | 4.14E-01 |
| Black | WHICAP | CDR | 1.20E-03 | 2.17E-03 | 2.62E-01 | 1.04E-01 | 4.19E-01 |
| Black | WHICAP | Cog. Impair. | 1.08E-02 | 1.53E-02 | 2.93E-01 | 7.24E-02 | 5.25E-01 |
| Black | WHICAP | Dementia | 6.67E-04 | 1.27E-03 | 6.96E-01 | 3.13E-01 | 1.12E+00 |
| Hispanic | CogDRisk | Hippocampal Volume | 1.48E-29 | 4.44E-28 | -3.67E-01 | -4.28E-01 | -3.06E-01 |
| Hispanic | CogDRisk | Cortical Thickness | 9.81E-17 | 9.62E-16 | -3.11E-01 | -3.83E-01 | -2.39E-01 |
| Hispanic | CogDRisk | MMSE | 1.83E-19 | 2.47E-18 | -2.89E-01 | -3.50E-01 | -2.27E-01 |
| Hispanic | CogDRisk | Aβ42/Aβ40 | 7.81E-06 | 2.14E-05 | -1.77E-01 | -2.54E-01 | -9.97E-02 |
| Hispanic | CogDRisk | Executive Function | 1.00E-03 | 1.88E-03 | -1.17E-01 | -1.87E-01 | -4.76E-02 |
| Hispanic | CogDRisk | Verbal Ability | 1.70E-01 | 1.88E-01 | -4.87E-02 | -1.18E-01 | 2.09E-02 |
| Hispanic | CogDRisk | Memory | 2.60E-01 | 2.73E-01 | -4.08E-02 | -1.12E-01 | 3.02E-02 |
| Hispanic | CogDRisk | Total Tau | 5.76E-02 | 7.01E-02 | 7.03E-02 | -2.29E-03 | 1.43E-01 |
| Hispanic | CogDRisk | pTau | 1.67E-04 | 3.59E-04 | 1.36E-01 | 6.54E-02 | 2.06E-01 |
| Hispanic | CogDRisk | CDR | 5.89E-06 | 1.64E-05 | 1.58E-01 | 9.02E-02 | 2.26E-01 |
| Hispanic | CogDRisk | MCI | 1.17E-02 | 1.64E-02 | 2.25E-01 | 4.97E-02 | 4.00E-01 |
| Hispanic | CogDRisk | Cog. Impair. | 1.79E-04 | 3.81E-04 | 2.99E-01 | 1.43E-01 | 4.57E-01 |
| Hispanic | CogDRisk | NfL | 7.33E-25 | 1.44E-23 | 3.52E-01 | 2.87E-01 | 4.16E-01 |

| | | | | | | | |
|---|---|---|---|---|---|---|---|
| Hispanic | CogDRisk | Dementia | 2.02E-04 | 4.21E-04 | 4.88E-01 | 2.30E-01 | 7.46E-01 |
| Hispanic | LIBRA | Hippocampal Volume | 3.29E-05 | 8.12E-05 | -1.39E-01 | -2.04E-01 | -7.37E-02 |
| Hispanic | LIBRA | Verbal Ability | 4.73E-04 | 9.33E-04 | -1.23E-01 | -1.92E-01 | -5.43E-02 |
| Hispanic | LIBRA | Executive Function | 1.15E-03 | 2.11E-03 | -1.15E-01 | -1.85E-01 | -4.60E-02 |
| Hispanic | LIBRA | MMSE | 2.13E-03 | 3.62E-03 | -1.00E-01 | -1.64E-01 | -3.64E-02 |
| Hispanic | LIBRA | Memory | 5.82E-02 | 7.03E-02 | -6.82E-02 | -1.39E-01 | 2.39E-03 |
| Hispanic | LIBRA | Cortical Thickness | 1.11E-01 | 1.28E-01 | -6.08E-02 | -1.36E-01 | 1.40E-02 |
| Hispanic | LIBRA | Aβ42/Aβ40 | 1.71E-01 | 1.89E-01 | -5.04E-02 | -1.23E-01 | 2.18E-02 |
| Hispanic | LIBRA | CDR | 1.92E-04 | 4.03E-04 | 1.30E-01 | 6.19E-02 | 1.98E-01 |
| Hispanic | LIBRA | pTau | 4.47E-05 | 1.09E-04 | 1.49E-01 | 7.79E-02 | 2.21E-01 |
| Hispanic | LIBRA | MCI | 8.16E-02 | 9.63E-02 | 1.58E-01 | -1.96E-02 | 3.37E-01 |
| Hispanic | LIBRA | Total Tau | 1.38E-06 | 4.48E-06 | 1.75E-01 | 1.05E-01 | 2.46E-01 |
| Hispanic | LIBRA | Cog. Impair. | 4.48E-03 | 7.02E-03 | 2.31E-01 | 7.23E-02 | 3.91E-01 |
| Hispanic | LIBRA | NfL | 1.07E-13 | 8.57E-13 | 2.67E-01 | 1.97E-01 | 3.36E-01 |
| Hispanic | LIBRA | Dementia | 1.81E-03 | 3.13E-03 | 4.46E-01 | 1.68E-01 | 7.30E-01 |
| Hispanic | mCAIDE | Hippocampal Volume | 3.42E-10 | 1.86E-09 | -2.11E-01 | -2.76E-01 | -1.46E-01 |
| Hispanic | mCAIDE | Cortical Thickness | 3.59E-07 | 1.29E-06 | -1.93E-01 | -2.67E-01 | -1.19E-01 |
| Hispanic | mCAIDE | MMSE | 7.15E-07 | 2.43E-06 | -1.62E-01 | -2.25E-01 | -9.81E-02 |
| Hispanic | mCAIDE | Aβ42/Aβ40 | 1.99E-03 | 3.39E-03 | -1.19E-01 | -1.95E-01 | -4.39E-02 |
| Hispanic | mCAIDE | Verbal Ability | 4.28E-03 | 6.78E-03 | -1.01E-01 | -1.71E-01 | -3.19E-02 |
| Hispanic | mCAIDE | Executive Function | 1.23E-01 | 1.40E-01 | -5.52E-02 | -1.25E-01 | 1.50E-02 |
| Hispanic | mCAIDE | Memory | 1.88E-01 | 2.03E-01 | -4.77E-02 | -1.19E-01 | 2.33E-02 |
| Hispanic | mCAIDE | pTau | 3.54E-01 | 3.67E-01 | 3.28E-02 | -3.66E-02 | 1.02E-01 |
| Hispanic | mCAIDE | Total Tau | 1.49E-01 | 1.67E-01 | 5.30E-02 | -1.90E-02 | 1.25E-01 |
| Hispanic | mCAIDE | CDR | 8.17E-02 | 9.63E-02 | 6.12E-02 | -7.73E-03 | 1.30E-01 |
| Hispanic | mCAIDE | MCI | 3.89E-01 | 4.00E-01 | 7.78E-02 | -9.87E-02 | 2.55E-01 |
| Hispanic | mCAIDE | NfL | 7.73E-04 | 1.46E-03 | 1.16E-01 | 4.85E-02 | 1.83E-01 |
| Hispanic | mCAIDE | Cog. Impair. | 7.60E-02 | 9.03E-02 | 1.43E-01 | -1.43E-02 | 3.01E-01 |
| Hispanic | mCAIDE | Dementia | 2.51E-02 | 3.25E-02 | 3.17E-01 | 4.24E-02 | 5.98E-01 |
| Hispanic | WHICAP | MMSE | 2.58E-22 | 4.33E-21 | -3.32E-01 | -3.97E-01 | -2.67E-01 |
| Hispanic | WHICAP | Hippocampal Volume | 1.69E-09 | 7.78E-09 | -2.01E-01 | -2.66E-01 | -1.36E-01 |
| Hispanic | WHICAP | Cortical Thickness | 1.71E-07 | 6.41E-07 | -1.99E-01 | -2.72E-01 | -1.25E-01 |
| Hispanic | WHICAP | Aβ42/Aβ40 | 1.42E-03 | 2.51E-03 | -1.19E-01 | -1.91E-01 | -4.60E-02 |
| Hispanic | WHICAP | Executive Function | 1.72E-02 | 2.34E-02 | -9.13E-02 | -1.66E-01 | -1.62E-02 |
| Hispanic | WHICAP | Verbal Ability | 2.16E-02 | 2.84E-02 | -8.74E-02 | -1.62E-01 | -1.29E-02 |
| Hispanic | WHICAP | Memory | 8.34E-02 | 9.78E-02 | -6.70E-02 | -1.43E-01 | 8.87E-03 |

| | | | | | | | |
|---|---|---|---|---|---|---|---|
| Hispanic | WHICAP | Total Tau | 1.75E-01 | 1.92E-01 | 4.81E-02 | -2.15E-02 | 1.18E-01 |
| Hispanic | WHICAP | pTau | 2.18E-02 | 2.84E-02 | 7.81E-02 | 1.14E-02 | 1.45E-01 |
| Hispanic | WHICAP | MCI | 1.94E-01 | 2.08E-01 | 1.17E-01 | -6.00E-02 | 2.95E-01 |
| Hispanic | WHICAP | CDR | 7.96E-05 | 1.85E-04 | 1.47E-01 | 7.45E-02 | 2.20E-01 |
| Hispanic | WHICAP | Cog. Impair. | 4.80E-03 | 7.41E-03 | 2.27E-01 | 6.96E-02 | 3.85E-01 |
| Hispanic | WHICAP | NfL | 4.39E-13 | 3.30E-12 | 2.39E-01 | 1.76E-01 | 3.03E-01 |
| Hispanic | WHICAP | Dementia | 1.40E-04 | 3.06E-04 | 5.31E-01 | 2.60E-01 | 8.08E-01 |
| NHW | CogDRisk | Hippocampal Volume | 3.55E-53 | 2.79E-51 | -4.62E-01 | -5.17E-01 | -4.07E-01 |
| NHW | CogDRisk | Cortical Thickness | 8.59E-26 | 1.86E-24 | -3.72E-01 | -4.39E-01 | -3.05E-01 |
| NHW | CogDRisk | Executive Function | 3.92E-10 | 2.05E-09 | -1.96E-01 | -2.56E-01 | -1.35E-01 |
| NHW | CogDRisk | MMSE | 4.02E-10 | 2.07E-09 | -1.95E-01 | -2.56E-01 | -1.35E-01 |
| NHW | CogDRisk | Memory | 5.74E-09 | 2.44E-08 | -1.85E-01 | -2.47E-01 | -1.23E-01 |
| NHW | CogDRisk | Verbal Ability | 8.06E-05 | 1.85E-04 | -1.24E-01 | -1.86E-01 | -6.26E-02 |
| NHW | CogDRisk | Aβ42/Aβ40 | 1.95E-03 | 3.35E-03 | -1.10E-01 | -1.80E-01 | -4.06E-02 |
| NHW | CogDRisk | Total Tau | 7.41E-01 | 7.50E-01 | -1.16E-02 | -8.04E-02 | 5.72E-02 |
| NHW | CogDRisk | CDR | 1.86E-04 | 3.93E-04 | 1.17E-01 | 5.57E-02 | 1.78E-01 |
| NHW | CogDRisk | pTau | 7.17E-17 | 7.33E-16 | 2.74E-01 | 2.11E-01 | 3.37E-01 |
| NHW | CogDRisk | MCI | 1.63E-03 | 2.87E-03 | 3.02E-01 | 1.14E-01 | 4.90E-01 |
| NHW | CogDRisk | NfL | 1.85E-30 | 6.29E-29 | 3.43E-01 | 2.86E-01 | 3.99E-01 |
| NHW | CogDRisk | Cog. Impair. | 8.75E-06 | 2.33E-05 | 3.78E-01 | 2.12E-01 | 5.46E-01 |
| NHW | CogDRisk | Dementia | 8.87E-05 | 2.01E-04 | 5.95E-01 | 2.99E-01 | 8.96E-01 |
| NHW | LIBRA | Verbal Ability | 6.05E-03 | 9.06E-03 | -8.93E-02 | -1.53E-01 | -2.56E-02 |
| NHW | LIBRA | Memory | 8.52E-03 | 1.26E-02 | -8.56E-02 | -1.49E-01 | -2.19E-02 |
| NHW | LIBRA | MMSE | 1.31E-02 | 1.79E-02 | -8.05E-02 | -1.44E-01 | -1.70E-02 |
| NHW | LIBRA | Hippocampal Volume | 1.59E-02 | 2.17E-02 | -7.79E-02 | -1.41E-01 | -1.46E-02 |
| NHW | LIBRA | Executive Function | 2.11E-02 | 2.80E-02 | -7.51E-02 | -1.39E-01 | -1.13E-02 |
| NHW | LIBRA | Cortical Thickness | 2.72E-01 | 2.84E-01 | -4.08E-02 | -1.14E-01 | 3.20E-02 |
| NHW | LIBRA | pTau | 2.14E-01 | 2.28E-01 | 4.09E-02 | -2.36E-02 | 1.05E-01 |
| NHW | LIBRA | Aβ42/Aβ40 | 2.20E-02 | 2.86E-02 | 7.65E-02 | 1.11E-02 | 1.42E-01 |
| NHW | LIBRA | CDR | 7.49E-03 | 1.12E-02 | 8.61E-02 | 2.30E-02 | 1.49E-01 |
| NHW | LIBRA | NfL | 5.09E-03 | 7.69E-03 | 9.33E-02 | 2.81E-02 | 1.58E-01 |
| NHW | LIBRA | Total Tau | 3.24E-04 | 6.54E-04 | 1.21E-01 | 5.50E-02 | 1.86E-01 |
| NHW | LIBRA | MCI | 2.29E-03 | 3.86E-03 | 2.96E-01 | 1.06E-01 | 4.87E-01 |
| NHW | LIBRA | Cog. Impair. | 2.85E-04 | 5.82E-04 | 3.10E-01 | 1.43E-01 | 4.79E-01 |
| NHW | LIBRA | Dementia | 1.82E-02 | 2.46E-02 | 3.58E-01 | 6.11E-02 | 6.57E-01 |
| NHW | mCAIDE | Hippocampal Volume | 7.01E-12 | 4.63E-11 | -2.24E-01 | -2.88E-01 | -1.61E-01 |

| | | | | | | | |
|---|---|---|---|---|---|---|---|
| NHW | mCAIDE | Cortical Thickness | 5.57E-06 | 1.58E-05 | -1.71E-01 | -2.44E-01 | -9.75E-02 |
| NHW | mCAIDE | Memory | 2.65E-07 | 9.69E-07 | -1.68E-01 | -2.32E-01 | -1.05E-01 |
| NHW | mCAIDE | MMSE | 1.33E-06 | 4.35E-06 | -1.58E-01 | -2.21E-01 | -9.42E-02 |
| NHW | mCAIDE | Verbal Ability | 3.73E-04 | 7.41E-04 | -1.17E-01 | -1.81E-01 | -5.26E-02 |
| NHW | mCAIDE | Executive Function | 4.62E-03 | 7.20E-03 | -9.34E-02 | -1.58E-01 | -2.88E-02 |
| NHW | mCAIDE | Total Tau | 7.98E-01 | 8.04E-01 | 8.65E-03 | -5.75E-02 | 7.48E-02 |
| NHW | mCAIDE | Aβ42/Aβ40 | 7.40E-01 | 7.50E-01 | 1.13E-02 | -5.53E-02 | 7.79E-02 |
| NHW | mCAIDE | NfL | 1.02E-01 | 1.19E-01 | 4.86E-02 | -9.72E-03 | 1.07E-01 |
| NHW | mCAIDE | pTau | 4.55E-02 | 5.68E-02 | 6.38E-02 | 1.29E-03 | 1.26E-01 |
| NHW | mCAIDE | CDR | 5.21E-03 | 7.84E-03 | 9.11E-02 | 2.72E-02 | 1.55E-01 |
| NHW | mCAIDE | MCI | 1.04E-02 | 1.48E-02 | 2.54E-01 | 6.05E-02 | 4.50E-01 |
| NHW | mCAIDE | Cog. Impair. | 1.07E-03 | 1.99E-03 | 2.85E-01 | 1.15E-01 | 4.58E-01 |
| NHW | mCAIDE | Dementia | 2.09E-02 | 2.78E-02 | 3.63E-01 | 5.68E-02 | 6.74E-01 |
| NHW | WHICAP | Hippocampal Volume | 5.11E-35 | 2.07E-33 | -3.82E-01 | -4.41E-01 | -3.24E-01 |
| NHW | WHICAP | Cortical Thickness | 1.32E-17 | 1.69E-16 | -3.09E-01 | -3.78E-01 | -2.40E-01 |
| NHW | WHICAP | MMSE | 2.58E-07 | 9.53E-07 | -2.02E-01 | -2.78E-01 | -1.26E-01 |
| NHW | WHICAP | Executive Function | 1.00E-06 | 3.32E-06 | -1.92E-01 | -2.69E-01 | -1.16E-01 |
| NHW | WHICAP | Memory | 3.83E-06 | 1.12E-05 | -1.84E-01 | -2.61E-01 | -1.06E-01 |
| NHW | WHICAP | Verbal Ability | 3.28E-06 | 9.77E-06 | -1.83E-01 | -2.60E-01 | -1.06E-01 |
| NHW | WHICAP | Aβ42/Aβ40 | 9.63E-02 | 1.12E-01 | -5.80E-02 | -1.26E-01 | 1.04E-02 |
| NHW | WHICAP | Total Tau | 9.63E-01 | 9.63E-01 | -1.59E-03 | -6.93E-02 | 6.62E-02 |
| NHW | WHICAP | CDR | 1.16E-01 | 1.33E-01 | 6.16E-02 | -1.52E-02 | 1.38E-01 |
| NHW | WHICAP | pTau | 1.86E-06 | 5.88E-06 | 1.55E-01 | 9.14E-02 | 2.18E-01 |
| NHW | WHICAP | NfL | 4.55E-10 | 2.30E-09 | 1.88E-01 | 1.30E-01 | 2.47E-01 |
| NHW | WHICAP | MCI | 4.98E-03 | 7.64E-03 | 2.64E-01 | 7.89E-02 | 4.49E-01 |
| NHW | WHICAP | Cog. Impair. | 1.30E-03 | 2.32E-03 | 2.70E-01 | 1.05E-01 | 4.35E-01 |
| NHW | WHICAP | Dementia | 5.49E-02 | 6.73E-02 | 2.86E-01 | -9.88E-03 | 5.77E-01 |
| all | CogDRisk | Hippocampal Volume | 2.36E-82 | 5.92E-80 | -3.91E-01 | -4.29E-01 | -3.53E-01 |
| all | CogDRisk | Cortical Thickness | 7.94E-50 | 4.75E-48 | -3.53E-01 | -3.98E-01 | -3.08E-01 |
| all | CogDRisk | MMSE | 1.47E-26 | 3.48E-25 | -2.04E-01 | -2.41E-01 | -1.67E-01 |
| all | CogDRisk | Aβ42/Aβ40 | 1.81E-10 | 1.01E-09 | -1.51E-01 | -1.97E-01 | -1.05E-01 |
| all | CogDRisk | Executive Function | 3.88E-10 | 2.05E-09 | -1.38E-01 | -1.80E-01 | -9.47E-02 |
| all | CogDRisk | Memory | 1.12E-05 | 2.93E-05 | -9.70E-02 | -1.40E-01 | -5.38E-02 |
| all | CogDRisk | Verbal Ability | 5.21E-05 | 1.25E-04 | -8.89E-02 | -1.32E-01 | -4.59E-02 |
| all | CogDRisk | Total Tau | 3.91E-03 | 6.26E-03 | 6.59E-02 | 2.11E-02 | 1.11E-01 |
| all | CogDRisk | CDR | 3.12E-08 | 1.24E-07 | 1.20E-01 | 7.78E-02 | 1.63E-01 |

| | | | | | | | |
|---|---|---|---|---|---|---|---|
| all | CogDRisk | MCI | 2.72E-02 | 3.50E-02 | 1.22E-01 | 1.34E-02 | 2.29E-01 |
| all | CogDRisk | pTau | 3.11E-23 | 5.59E-22 | 2.14E-01 | 1.72E-01 | 2.55E-01 |
| all | CogDRisk | Cog. Impair. | 5.61E-06 | 1.58E-05 | 2.25E-01 | 1.28E-01 | 3.23E-01 |
| all | CogDRisk | NfL | 3.22E-74 | 3.68E-72 | 3.70E-01 | 3.32E-01 | 4.08E-01 |
| all | CogDRisk | Dementia | 8.61E-10 | 4.11E-09 | 5.34E-01 | 3.64E-01 | 7.05E-01 |
| all | LIBRA | Verbal Ability | 1.44E-09 | 6.74E-09 | -1.33E-01 | -1.76E-01 | -9.01E-02 |
| all | LIBRA | Executive Function | 1.78E-09 | 8.04E-09 | -1.33E-01 | -1.76E-01 | -8.96E-02 |
| all | LIBRA | MMSE | 3.98E-08 | 1.56E-07 | -1.07E-01 | -1.44E-01 | -6.87E-02 |
| all | LIBRA | Memory | 2.61E-06 | 7.97E-06 | -1.04E-01 | -1.47E-01 | -6.07E-02 |
| all | LIBRA | Hippocampal Volume | 5.61E-05 | 1.32E-04 | -8.59E-02 | -1.28E-01 | -4.42E-02 |
| all | LIBRA | Cortical Thickness | 5.18E-02 | 6.38E-02 | -4.78E-02 | -9.60E-02 | 3.82E-04 |
| all | LIBRA | Aβ42/Aβ40 | 5.30E-01 | 5.44E-01 | -1.43E-02 | -5.89E-02 | 3.03E-02 |
| all | LIBRA | pTau | 1.19E-02 | 1.66E-02 | 5.61E-02 | 1.24E-02 | 9.99E-02 |
| all | LIBRA | NfL | 1.43E-08 | 5.79E-08 | 1.27E-01 | 8.32E-02 | 1.71E-01 |
| all | LIBRA | CDR | 7.81E-10 | 3.79E-09 | 1.34E-01 | 9.14E-02 | 1.76E-01 |
| all | LIBRA | Total Tau | 3.30E-12 | 2.24E-11 | 1.56E-01 | 1.13E-01 | 2.00E-01 |
| all | LIBRA | MCI | 4.18E-07 | 1.49E-06 | 2.84E-01 | 1.74E-01 | 3.94E-01 |
| all | LIBRA | Cog. Impair. | 1.58E-10 | 9.02E-10 | 3.26E-01 | 2.27E-01 | 4.27E-01 |
| all | LIBRA | Dementia | 6.41E-07 | 2.21E-06 | 4.57E-01 | 2.78E-01 | 6.39E-01 |
| all | mCAIDE | Hippocampal Volume | 6.89E-17 | 7.33E-16 | -1.77E-01 | -2.19E-01 | -1.36E-01 |
| all | mCAIDE | Cortical Thickness | 1.84E-12 | 1.27E-11 | -1.73E-01 | -2.20E-01 | -1.25E-01 |
| all | mCAIDE | MMSE | 2.42E-15 | 2.21E-14 | -1.54E-01 | -1.92E-01 | -1.16E-01 |
| all | mCAIDE | Verbal Ability | 6.59E-10 | 3.26E-09 | -1.37E-01 | -1.80E-01 | -9.36E-02 |
| all | mCAIDE | Memory | 2.21E-09 | 9.82E-09 | -1.33E-01 | -1.77E-01 | -8.98E-02 |
| all | mCAIDE | Executive Function | 2.26E-06 | 7.07E-06 | -1.05E-01 | -1.49E-01 | -6.18E-02 |
| all | mCAIDE | Aβ42/Aβ40 | 9.38E-03 | 1.36E-02 | -5.98E-02 | -1.05E-01 | -1.47E-02 |
| all | mCAIDE | pTau | 5.11E-02 | 6.33E-02 | 4.13E-02 | -2.01E-04 | 8.29E-02 |
| all | mCAIDE | Total Tau | 5.04E-03 | 7.69E-03 | 6.28E-02 | 1.89E-02 | 1.07E-01 |
| all | mCAIDE | CDR | 5.52E-05 | 1.31E-04 | 8.89E-02 | 4.57E-02 | 1.32E-01 |
| all | mCAIDE | NfL | 1.03E-05 | 2.73E-05 | 9.16E-02 | 5.09E-02 | 1.32E-01 |
| all | mCAIDE | MCI | 3.13E-04 | 6.35E-04 | 2.01E-01 | 9.22E-02 | 3.11E-01 |
| all | mCAIDE | Cog. Impair. | 4.36E-07 | 1.52E-06 | 2.56E-01 | 1.57E-01 | 3.56E-01 |
| all | mCAIDE | Dementia | 5.29E-06 | 1.53E-05 | 4.23E-01 | 2.42E-01 | 6.07E-01 |
| all | WHICAP | MMSE | 4.75E-43 | 2.29E-41 | -3.01E-01 | -3.43E-01 | -2.59E-01 |
| all | WHICAP | Executive Function | 4.24E-17 | 4.98E-16 | -2.13E-01 | -2.62E-01 | -1.64E-01 |
| all | WHICAP | Cortical Thickness | 4.79E-17 | 5.37E-16 | -2.07E-01 | -2.55E-01 | -1.59E-01 |

| | | | | | | | |
|---|---|---|---|---|---|---|---|
| all | WHICAP | Hippocampal Volume | 5.01E-20 | 7.15E-19 | -1.96E-01 | -2.38E-01 | -1.55E-01 |
| all | WHICAP | Verbal Ability | 9.87E-15 | 8.43E-14 | -1.95E-01 | -2.45E-01 | -1.46E-01 |
| all | WHICAP | Memory | 1.46E-12 | 1.04E-11 | -1.79E-01 | -2.29E-01 | -1.30E-01 |
| all | WHICAP | Aβ42/Aβ40 | 1.45E-06 | 4.65E-06 | -1.10E-01 | -1.54E-01 | -6.52E-02 |
| all | WHICAP | pTau | 1.94E-02 | 2.61E-02 | 4.91E-02 | 7.94E-03 | 9.03E-02 |
| all | WHICAP | Total Tau | 9.57E-05 | 2.13E-04 | 8.66E-02 | 4.32E-02 | 1.30E-01 |
| all | WHICAP | CDR | 8.52E-11 | 5.15E-10 | 1.62E-01 | 1.14E-01 | 2.11E-01 |
| all | WHICAP | NfL | 4.43E-16 | 4.18E-15 | 1.67E-01 | 1.27E-01 | 2.06E-01 |
| all | WHICAP | MCI | 2.76E-06 | 8.34E-06 | 2.60E-01 | 1.51E-01 | 3.69E-01 |
| all | WHICAP | Cog. Impair. | 2.17E-11 | 1.34E-10 | 3.38E-01 | 2.39E-01 | 4.37E-01 |
| all | WHICAP | Dementia | 1.27E-10 | 7.53E-10 | 5.66E-01 | 3.94E-01 | 7.39E-01 |
| Black | CogDRisk (sensitivity) | | 2.82E-01 | 4.14E-01 | -1.17E-01 | -3.32E-01 | 9.56E-02 |
| Black | CogDRisk (sensitivity) | Aβ42/Aβ40 | 7.92E-02 | 1.61E-01 | -1.00E-01 | -2.12E-01 | 1.18E-02 |
| Black | CogDRisk (sensitivity) | MMSE | 5.08E-01 | 6.39E-01 | -3.69E-02 | -1.47E-01 | 7.27E-02 |
| Black | CogDRisk (sensitivity) | Executive Function | 1.89E-01 | 3.12E-01 | -7.30E-02 | -1.82E-01 | 3.61E-02 |
| Black | CogDRisk (sensitivity) | Verbal Ability | 2.46E-01 | 3.73E-01 | -6.44E-02 | -1.73E-01 | 4.45E-02 |
| Black | CogDRisk (sensitivity) | pTau | 3.96E-01 | 5.32E-01 | -4.43E-02 | -1.47E-01 | 5.83E-02 |
| Black | CogDRisk (sensitivity) | Cortical Thickness | 1.29E-01 | 2.28E-01 | -1.02E-01 | -2.34E-01 | 3.00E-02 |
| Black | CogDRisk (sensitivity) | Memory | 8.54E-01 | 8.99E-01 | -1.04E-02 | -1.21E-01 | 9.99E-02 |
| Black | CogDRisk (sensitivity) | Hippocampal Volume | 9.37E-01 | 9.47E-01 | -3.98E-03 | -1.04E-01 | 9.59E-02 |
| Black | CogDRisk (sensitivity) | NfL | 6.57E-01 | 7.67E-01 | 2.40E-02 | -8.23E-02 | 1.30E-01 |
| Black | CogDRisk (sensitivity) | MCI | 5.21E-01 | 6.45E-01 | 7.33E-02 | -1.51E-01 | 2.98E-01 |
| Black | CogDRisk (sensitivity) | Cog. Impair. | 2.82E-01 | 4.14E-01 | 1.17E-01 | -9.56E-02 | 3.32E-01 |
| Black | CogDRisk (sensitivity) | Total Tau | 1.44E-03 | 6.57E-03 | 1.74E-01 | 6.75E-02 | 2.81E-01 |
| Black | CogDRisk (sensitivity) | CDR | 3.24E-02 | 8.16E-02 | 1.19E-01 | 1.01E-02 | 2.29E-01 |
| Black | CogDRisk (sensitivity) | Dementia | 1.07E-01 | 2.02E-01 | 3.23E-01 | -7.07E-02 | 7.20E-01 |
| Black | mCAIDE (sensitivity) | | 1.03E-01 | 1.98E-01 | -1.75E-01 | -3.88E-01 | 3.42E-02 |
| Black | mCAIDE (sensitivity) | Verbal Ability | 1.06E-02 | 3.44E-02 | -1.41E-01 | -2.49E-01 | -3.31E-02 |
| Black | mCAIDE (sensitivity) | Executive Function | 4.81E-02 | 1.13E-01 | -1.09E-01 | -2.18E-01 | -9.14E-04 |
| Black | mCAIDE (sensitivity) | MMSE | 6.56E-01 | 7.67E-01 | -2.48E-02 | -1.34E-01 | 8.46E-02 |
| Black | mCAIDE (sensitivity) | Hippocampal Volume | 2.07E-01 | 3.33E-01 | -6.16E-02 | -1.57E-01 | 3.42E-02 |
| Black | mCAIDE (sensitivity) | Memory | 6.04E-01 | 7.21E-01 | -2.90E-02 | -1.39E-01 | 8.10E-02 |

| | | | | | | | |
|---|---|---|---|---|---|---|---|
| Black | mCAIDE (sensitivity) | NfL | 6.63E-01 | 7.68E-01 | -2.35E-02 | -1.30E-01 | 8.27E-02 |
| Black | mCAIDE (sensitivity) | pTau | 7.65E-01 | 8.68E-01 | -1.56E-02 | -1.18E-01 | 8.69E-02 |
| Black | mCAIDE (sensitivity) | Aβ42/Aβ40 | 9.38E-01 | 9.47E-01 | -4.43E-03 | -1.16E-01 | 1.08E-01 |
| Black | mCAIDE (sensitivity) | Cortical Thickness | 8.07E-01 | 8.86E-01 | -1.62E-02 | -1.46E-01 | 1.14E-01 |
| Black | mCAIDE (sensitivity) | CDR | 3.28E-01 | 4.62E-01 | 5.46E-02 | -5.50E-02 | 1.64E-01 |
| Black | mCAIDE (sensitivity) | Total Tau | 7.70E-02 | 1.61E-01 | 9.68E-02 | -1.06E-02 | 2.04E-01 |
| Black | mCAIDE (sensitivity) | MCI | 2.05E-01 | 3.33E-01 | 1.43E-01 | -7.72E-02 | 3.65E-01 |
| Black | mCAIDE (sensitivity) | Cog. Impair. | 1.03E-01 | 1.98E-01 | 1.75E-01 | -3.42E-02 | 3.88E-01 |
| Black | mCAIDE (sensitivity) | Dementia | 7.18E-02 | 1.53E-01 | 3.80E-01 | -2.77E-02 | 8.06E-01 |
| Black | WHICAP (sensitivity) | Aβ42/Aβ40 | 7.75E-02 | 1.61E-01 | -1.01E-01 | -2.14E-01 | 1.12E-02 |
| Black | WHICAP (sensitivity) | | 2.98E-01 | 4.33E-01 | -1.14E-01 | -3.31E-01 | 9.96E-02 |
| Black | WHICAP (sensitivity) | Hippocampal Volume | 4.90E-02 | 1.14E-01 | -9.87E-02 | -1.97E-01 | -4.43E-04 |
| Black | WHICAP (sensitivity) | pTau | 1.04E-01 | 1.98E-01 | -8.51E-02 | -1.88E-01 | 1.75E-02 |
| Black | WHICAP (sensitivity) | NfL | 7.03E-02 | 1.51E-01 | -9.79E-02 | -2.04E-01 | 8.17E-03 |
| Black | WHICAP (sensitivity) | Verbal Ability | 1.09E-01 | 2.04E-01 | -1.20E-01 | -2.67E-01 | 2.70E-02 |
| Black | WHICAP (sensitivity) | Executive Function | 4.12E-01 | 5.46E-01 | -6.17E-02 | -2.09E-01 | 8.59E-02 |
| Black | WHICAP (sensitivity) | Dementia | 9.56E-01 | 9.61E-01 | -1.15E-02 | -4.17E-01 | 4.10E-01 |
| Black | WHICAP (sensitivity) | Cortical Thickness | 3.67E-01 | 5.00E-01 | 6.04E-02 | -7.14E-02 | 1.92E-01 |
| Black | WHICAP (sensitivity) | Memory | 3.54E-01 | 4.92E-01 | 7.03E-02 | -7.86E-02 | 2.19E-01 |
| Black | WHICAP (sensitivity) | CDR | 6.42E-01 | 7.62E-01 | 3.51E-02 | -1.13E-01 | 1.84E-01 |
| Black | WHICAP (sensitivity) | Total Tau | 3.63E-01 | 4.98E-01 | 5.03E-02 | -5.83E-02 | 1.59E-01 |
| Black | WHICAP (sensitivity) | MMSE | 5.05E-01 | 6.39E-01 | 5.03E-02 | -9.79E-02 | 1.98E-01 |
| Black | WHICAP (sensitivity) | Cog. Impair. | 2.98E-01 | 4.33E-01 | 1.14E-01 | -9.96E-02 | 3.31E-01 |
| Black | WHICAP (sensitivity) | MCI | 2.12E-01 | 3.36E-01 | 1.45E-01 | -8.06E-02 | 3.76E-01 |
| Hispanic | CogDRisk (sensitivity) | MMSE | 4.74E-02 | 1.13E-01 | -6.46E-02 | -1.29E-01 | -7.30E-04 |
| Hispanic | CogDRisk (sensitivity) | | 4.58E-02 | 1.10E-01 | -1.60E-01 | -3.18E-01 | -3.15E-03 |
| Hispanic | CogDRisk (sensitivity) | Hippocampal Volume | 7.95E-04 | 4.14E-03 | -1.13E-01 | -1.79E-01 | -4.71E-02 |
| Hispanic | CogDRisk (sensitivity) | Aβ42/Aβ40 | 2.84E-02 | 7.32E-02 | -8.16E-02 | -1.55E-01 | -8.67E-03 |
| Hispanic | CogDRisk (sensitivity) | Executive Function | 1.17E-02 | 3.67E-02 | -8.96E-02 | -1.59E-01 | -2.00E-02 |
| Hispanic | CogDRisk (sensitivity) | Verbal Ability | 8.53E-02 | 1.72E-01 | -6.08E-02 | -1.30E-01 | 8.47E-03 |

| | | | | | | | |
|---|---|---|---|---|---|---|---|
| Hispanic | CogDRisk (sensitivity) | Cortical Thickness | 1.35E-01 | 2.32E-01 | -5.71E-02 | -1.32E-01 | 1.79E-02 |
| Hispanic | CogDRisk (sensitivity) | pTau | 7.84E-01 | 8.70E-01 | 9.35E-03 | -5.74E-02 | 7.61E-02 |
| Hispanic | CogDRisk (sensitivity) | Memory | 5.52E-01 | 6.74E-01 | 2.15E-02 | -4.93E-02 | 9.22E-02 |
| Hispanic | CogDRisk (sensitivity) | CDR | 6.98E-03 | 2.51E-02 | 9.42E-02 | 2.59E-02 | 1.63E-01 |
| Hispanic | CogDRisk (sensitivity) | Total Tau | 5.29E-05 | 3.83E-04 | 1.42E-01 | 7.37E-02 | 2.11E-01 |
| Hispanic | CogDRisk (sensitivity) | MCI | 1.07E-01 | 2.02E-01 | 1.45E-01 | -3.14E-02 | 3.22E-01 |
| Hispanic | CogDRisk (sensitivity) | NfL | 1.19E-06 | 1.45E-05 | 1.62E-01 | 9.69E-02 | 2.27E-01 |
| Hispanic | CogDRisk (sensitivity) | Cog. Impair. | 4.58E-02 | 1.10E-01 | 1.60E-01 | 3.15E-03 | 3.18E-01 |
| Hispanic | CogDRisk (sensitivity) | Dementia | 1.50E-01 | 2.55E-01 | 1.97E-01 | -7.21E-02 | 4.65E-01 |
| Hispanic | mCAIDE (sensitivity) | MMSE | 1.88E-01 | 3.12E-01 | -4.30E-02 | -1.07E-01 | 2.11E-02 |
| Hispanic | mCAIDE (sensitivity) | Hippocampal Volume | 6.72E-03 | 2.46E-02 | -9.16E-02 | -1.58E-01 | -2.54E-02 |
| Hispanic | mCAIDE (sensitivity) | Verbal Ability | 9.97E-03 | 3.33E-02 | -9.11E-02 | -1.60E-01 | -2.19E-02 |
| Hispanic | mCAIDE (sensitivity) | Aβ42/Aβ40 | 4.53E-02 | 1.10E-01 | -7.54E-02 | -1.49E-01 | -1.59E-03 |
| Hispanic | mCAIDE (sensitivity) | Executive Function | 2.73E-01 | 4.06E-01 | -3.90E-02 | -1.09E-01 | 3.08E-02 |
| Hispanic | mCAIDE (sensitivity) | | 7.84E-01 | 8.70E-01 | -2.19E-02 | -1.78E-01 | 1.35E-01 |
| Hispanic | mCAIDE (sensitivity) | MCI | 8.77E-01 | 9.15E-01 | -1.39E-02 | -1.91E-01 | 1.62E-01 |
| Hispanic | mCAIDE (sensitivity) | Cortical Thickness | 1.96E-02 | 5.55E-02 | -8.90E-02 | -1.64E-01 | -1.43E-02 |
| Hispanic | mCAIDE (sensitivity) | pTau | 9.20E-01 | 9.39E-01 | -3.45E-03 | -7.11E-02 | 6.42E-02 |
| Hispanic | mCAIDE (sensitivity) | Memory | 8.53E-01 | 8.99E-01 | 6.69E-03 | -6.41E-02 | 7.75E-02 |
| Hispanic | mCAIDE (sensitivity) | Cog. Impair. | 7.84E-01 | 8.70E-01 | 2.19E-02 | -1.35E-01 | 1.78E-01 |
| Hispanic | mCAIDE (sensitivity) | CDR | 6.58E-01 | 7.67E-01 | 1.55E-02 | -5.32E-02 | 8.43E-02 |
| Hispanic | mCAIDE (sensitivity) | NfL | 3.52E-01 | 4.92E-01 | 3.15E-02 | -3.49E-02 | 9.78E-02 |
| Hispanic | mCAIDE (sensitivity) | Total Tau | 1.83E-03 | 8.16E-03 | 1.11E-01 | 4.14E-02 | 1.81E-01 |
| Hispanic | mCAIDE (sensitivity) | Dementia | 4.34E-01 | 5.66E-01 | 1.08E-01 | -1.63E-01 | 3.80E-01 |
| Hispanic | WHICAP (sensitivity) | Verbal Ability | 1.28E-02 | 3.83E-02 | -8.79E-02 | -1.57E-01 | -1.88E-02 |
| Hispanic | WHICAP (sensitivity) | Hippocampal Volume | 3.14E-02 | 7.98E-02 | -7.26E-02 | -1.39E-01 | -6.51E-03 |
| Hispanic | WHICAP (sensitivity) | | 5.15E-01 | 6.40E-01 | -5.25E-02 | -2.12E-01 | 1.04E-01 |
| Hispanic | WHICAP (sensitivity) | MMSE | 6.98E-01 | 8.03E-01 | -1.27E-02 | -7.67E-02 | 5.13E-02 |
| Hispanic | WHICAP (sensitivity) | Executive Function | 3.14E-01 | 4.45E-01 | -3.58E-02 | -1.06E-01 | 3.39E-02 |
| Hispanic | WHICAP (sensitivity) | Aβ42/Aβ40 | 5.63E-01 | 6.83E-01 | -2.15E-02 | -9.43E-02 | 5.13E-02 |

| | | | | | | | |
|---|---|---|---|---|---|---|---|
| Hispanic | WHICAP (sensitivity) | Memory | 8.38E-01 | 8.99E-01 | -7.38E-03 | -7.80E-02 | 6.32E-02 |
| Hispanic | WHICAP (sensitivity) | Cortical Thickness | 5.69E-01 | 6.87E-01 | -2.18E-02 | -9.69E-02 | 5.33E-02 |
| Hispanic | WHICAP (sensitivity) | pTau | 9.04E-01 | 9.34E-01 | 4.07E-03 | -6.23E-02 | 7.05E-02 |
| Hispanic | WHICAP (sensitivity) | MCI | 8.35E-01 | 8.99E-01 | 1.89E-02 | -1.58E-01 | 2.00E-01 |
| Hispanic | WHICAP (sensitivity) | NfL | 1.18E-01 | 2.15E-01 | 5.20E-02 | -1.32E-02 | 1.17E-01 |
| Hispanic | WHICAP (sensitivity) | Cog. Impair. | 5.15E-01 | 6.40E-01 | 5.25E-02 | -1.04E-01 | 2.12E-01 |
| Hispanic | WHICAP (sensitivity) | CDR | 2.11E-01 | 3.36E-01 | 4.36E-02 | -2.48E-02 | 1.12E-01 |
| Hispanic | WHICAP (sensitivity) | Total Tau | 3.71E-03 | 1.46E-02 | 1.02E-01 | 3.32E-02 | 1.71E-01 |
| Hispanic | WHICAP (sensitivity) | Dementia | 2.57E-01 | 3.88E-01 | 1.61E-01 | -1.11E-01 | 4.48E-01 |
| NHW | CogDRisk (sensitivity) | | 2.68E-03 | 1.13E-02 | -2.53E-01 | -4.19E-01 | -8.79E-02 |
| NHW | CogDRisk (sensitivity) | MMSE | 6.44E-02 | 1.40E-01 | -6.11E-02 | -1.26E-01 | 3.65E-03 |
| NHW | CogDRisk (sensitivity) | Hippocampal Volume | 9.57E-02 | 1.87E-01 | -5.34E-02 | -1.16E-01 | 9.45E-03 |
| NHW | CogDRisk (sensitivity) | Executive Function | 1.79E-01 | 2.99E-01 | -4.46E-02 | -1.10E-01 | 2.04E-02 |
| NHW | CogDRisk (sensitivity) | Verbal Ability | 2.28E-01 | 3.54E-01 | -3.99E-02 | -1.05E-01 | 2.51E-02 |
| NHW | CogDRisk (sensitivity) | Memory | 7.34E-01 | 8.37E-01 | -1.13E-02 | -7.65E-02 | 5.39E-02 |
| NHW | CogDRisk (sensitivity) | pTau | 9.73E-01 | 9.73E-01 | -1.04E-03 | -6.23E-02 | 6.02E-02 |
| NHW | CogDRisk (sensitivity) | Cortical Thickness | 8.51E-01 | 8.99E-01 | 6.89E-03 | -6.53E-02 | 7.91E-02 |
| NHW | CogDRisk (sensitivity) | NfL | 8.48E-01 | 8.99E-01 | 5.59E-03 | -5.17E-02 | 6.29E-02 |
| NHW | CogDRisk (sensitivity) | CDR | 1.20E-02 | 3.70E-02 | 8.24E-02 | 1.82E-02 | 1.47E-01 |
| NHW | CogDRisk (sensitivity) | Aβ42/Aβ40 | 9.44E-03 | 3.21E-02 | 8.63E-02 | 2.12E-02 | 1.51E-01 |
| NHW | CogDRisk (sensitivity) | Total Tau | 2.74E-05 | 2.15E-04 | 1.38E-01 | 7.35E-02 | 2.02E-01 |
| NHW | CogDRisk (sensitivity) | MCI | 2.15E-02 | 5.94E-02 | 2.20E-01 | 3.21E-02 | 4.08E-01 |
| NHW | CogDRisk (sensitivity) | Cog. Impair. | 2.68E-03 | 1.13E-02 | 2.53E-01 | 8.79E-02 | 4.19E-01 |
| NHW | CogDRisk (sensitivity) | Dementia | 2.12E-02 | 5.93E-02 | 3.38E-01 | 4.84E-02 | 6.26E-01 |
| NHW | mCAIDE (sensitivity) | | 6.12E-02 | 1.35E-01 | -1.61E-01 | -3.29E-01 | 7.85E-03 |
| NHW | mCAIDE (sensitivity) | MMSE | 1.53E-02 | 4.46E-02 | -7.96E-02 | -1.44E-01 | -1.53E-02 |
| NHW | mCAIDE (sensitivity) | Verbal Ability | 2.64E-02 | 6.91E-02 | -7.30E-02 | -1.37E-01 | -8.57E-03 |
| NHW | mCAIDE (sensitivity) | Hippocampal Volume | 1.26E-01 | 2.24E-01 | -4.97E-02 | -1.13E-01 | 1.40E-02 |
| NHW | mCAIDE (sensitivity) | Memory | 8.97E-02 | 1.79E-01 | -5.59E-02 | -1.21E-01 | 8.69E-03 |
| NHW | mCAIDE (sensitivity) | NfL | 1.25E-01 | 2.24E-01 | -4.50E-02 | -1.03E-01 | 1.25E-02 |

| | | | | | | | |
|---|---|---|---|---|---|---|---|
| NHW | mCAIDE (sensitivity) | pTau | 2.27E-01 | 3.54E-01 | -3.79E-02 | -9.95E-02 | 2.37E-02 |
| NHW | mCAIDE (sensitivity) | Executive Function | 4.15E-01 | 5.47E-01 | -2.70E-02 | -9.18E-02 | 3.79E-02 |
| NHW | mCAIDE (sensitivity) | Cortical Thickness | 5.77E-01 | 6.92E-01 | -2.07E-02 | -9.38E-02 | 5.23E-02 |
| NHW | mCAIDE (sensitivity) | CDR | 1.33E-01 | 2.32E-01 | 4.91E-02 | -1.50E-02 | 1.13E-01 |
| NHW | mCAIDE (sensitivity) | Aβ42/Aβ40 | 6.17E-02 | 1.35E-01 | 6.24E-02 | -3.06E-03 | 1.28E-01 |
| NHW | mCAIDE (sensitivity) | Total Tau | 1.87E-02 | 5.38E-02 | 7.78E-02 | 1.30E-02 | 1.43E-01 |
| NHW | mCAIDE (sensitivity) | MCI | 1.21E-01 | 2.19E-01 | 1.51E-01 | -4.07E-02 | 3.43E-01 |
| NHW | mCAIDE (sensitivity) | Cog. Impair. | 6.12E-02 | 1.35E-01 | 1.61E-01 | -7.85E-03 | 3.29E-01 |
| NHW | mCAIDE (sensitivity) | Dementia | 2.37E-01 | 3.62E-01 | 1.82E-01 | -1.22E-01 | 4.82E-01 |
| NHW | WHICAP (sensitivity) | NfL | 5.07E-05 | 3.82E-04 | -1.18E-01 | -1.74E-01 | -6.09E-02 |
| NHW | WHICAP (sensitivity) | Dementia | 4.77E-01 | 6.10E-01 | -1.07E-01 | -4.01E-01 | 1.89E-01 |
| NHW | WHICAP (sensitivity) | pTau | 4.23E-03 | 1.63E-02 | -8.90E-02 | -1.50E-01 | -2.81E-02 |
| NHW | WHICAP (sensitivity) | CDR | 2.32E-01 | 3.57E-01 | -4.59E-02 | -1.21E-01 | 2.94E-02 |
| NHW | WHICAP (sensitivity) | Verbal Ability | 2.61E-01 | 3.91E-01 | -4.36E-02 | -1.20E-01 | 3.24E-02 |
| NHW | WHICAP (sensitivity) | | 8.15E-01 | 8.87E-01 | -1.99E-02 | -1.88E-01 | 1.46E-01 |
| NHW | WHICAP (sensitivity) | MMSE | 7.92E-01 | 8.75E-01 | -1.02E-02 | -8.60E-02 | 6.57E-02 |
| NHW | WHICAP (sensitivity) | Hippocampal Volume | 9.05E-01 | 9.34E-01 | -3.93E-03 | -6.86E-02 | 6.08E-02 |
| NHW | WHICAP (sensitivity) | Cortical Thickness | 2.15E-01 | 3.38E-01 | 4.71E-02 | -2.74E-02 | 1.22E-01 |
| NHW | WHICAP (sensitivity) | Memory | 7.09E-01 | 8.13E-01 | 1.45E-02 | -6.16E-02 | 9.05E-02 |
| NHW | WHICAP (sensitivity) | Cog. Impair. | 8.15E-01 | 8.87E-01 | 1.99E-02 | -1.46E-01 | 1.88E-01 |
| NHW | WHICAP (sensitivity) | Executive Function | 4.22E-01 | 5.52E-01 | 3.12E-02 | -4.50E-02 | 1.07E-01 |
| NHW | WHICAP (sensitivity) | Total Tau | 5.76E-02 | 1.30E-01 | 6.26E-02 | -2.02E-03 | 1.27E-01 |
| NHW | WHICAP (sensitivity) | Aβ42/Aβ40 | 5.15E-02 | 1.18E-01 | 6.48E-02 | -4.18E-04 | 1.30E-01 |
| NHW | WHICAP (sensitivity) | MCI | 4.50E-01 | 5.79E-01 | 7.40E-02 | -1.16E-01 | 2.68E-01 |
| all | CogDRisk (sensitivity) | MMSE | 2.33E-04 | 1.46E-03 | -7.14E-02 | -1.09E-01 | -3.34E-02 |
| all | CogDRisk (sensitivity) | | 8.53E-06 | 6.98E-05 | -2.22E-01 | -3.20E-01 | -1.24E-01 |
| all | CogDRisk (sensitivity) | Executive Function | 8.45E-05 | 5.90E-04 | -8.68E-02 | -1.30E-01 | -4.36E-02 |
| all | CogDRisk (sensitivity) | Verbal Ability | 2.24E-03 | 9.79E-03 | -6.73E-02 | -1.10E-01 | -2.42E-02 |
| all | CogDRisk (sensitivity) | Hippocampal Volume | 9.32E-03 | 3.21E-02 | -5.56E-02 | -9.75E-02 | -1.37E-02 |
| all | CogDRisk (sensitivity) | Aβ42/Aβ40 | 4.04E-01 | 5.39E-01 | -1.89E-02 | -6.34E-02 | 2.56E-02 |

| | | | | | | | |
|---|---|---|---|---|---|---|---|
| all | CogDRisk (sensitivity) | pTau | 3.62E-01 | 4.98E-01 | -1.91E-02 | -6.00E-02 | 2.19E-02 |
| all | CogDRisk (sensitivity) | Memory | 3.86E-01 | 5.22E-01 | -1.92E-02 | -6.27E-02 | 2.42E-02 |
| all | CogDRisk (sensitivity) | Cortical Thickness | 1.95E-01 | 3.19E-01 | -3.20E-02 | -8.03E-02 | 1.64E-02 |
| all | CogDRisk (sensitivity) | NfL | 8.59E-04 | 4.35E-03 | 6.84E-02 | 2.82E-02 | 1.09E-01 |
| all | CogDRisk (sensitivity) | CDR | 1.39E-06 | 1.60E-05 | 1.05E-01 | 6.25E-02 | 1.48E-01 |
| all | CogDRisk (sensitivity) | Total Tau | 1.21E-13 | 2.28E-11 | 1.63E-01 | 1.20E-01 | 2.05E-01 |
| all | CogDRisk (sensitivity) | MCI | 6.35E-04 | 3.40E-03 | 1.88E-01 | 8.03E-02 | 2.97E-01 |
| all | CogDRisk (sensitivity) | Cog. Impair. | 8.53E-06 | 6.98E-05 | 2.22E-01 | 1.24E-01 | 3.20E-01 |
| all | CogDRisk (sensitivity) | Dementia | 3.31E-04 | 1.96E-03 | 3.17E-01 | 1.44E-01 | 4.90E-01 |
| all | mCAIDE (sensitivity) | MMSE | 5.71E-04 | 3.25E-03 | -6.67E-02 | -1.05E-01 | -2.88E-02 |
| all | mCAIDE (sensitivity) | | 5.94E-04 | 3.25E-03 | -1.71E-01 | -2.69E-01 | -7.37E-02 |
| all | mCAIDE (sensitivity) | Verbal Ability | 1.00E-06 | 1.31E-05 | -1.07E-01 | -1.50E-01 | -6.43E-02 |
| all | mCAIDE (sensitivity) | Executive Function | 1.07E-03 | 5.30E-03 | -7.20E-02 | -1.15E-01 | -2.89E-02 |
| all | mCAIDE (sensitivity) | Hippocampal Volume | 2.30E-02 | 6.25E-02 | -4.83E-02 | -9.00E-02 | -6.68E-03 |
| all | mCAIDE (sensitivity) | Memory | 1.21E-02 | 3.70E-02 | -5.54E-02 | -9.87E-02 | -1.21E-02 |
| all | mCAIDE (sensitivity) | pTau | 4.26E-02 | 1.06E-01 | -4.26E-02 | -8.38E-02 | -1.42E-03 |
| all | mCAIDE (sensitivity) | NfL | 3.03E-01 | 4.36E-01 | -2.13E-02 | -6.18E-02 | 1.92E-02 |
| all | mCAIDE (sensitivity) | Cortical Thickness | 7.91E-02 | 1.61E-01 | -4.30E-02 | -9.11E-02 | 5.00E-03 |
| all | mCAIDE (sensitivity) | Aβ42/Aβ40 | 9.18E-01 | 9.39E-01 | -2.35E-03 | -4.71E-02 | 4.24E-02 |
| all | mCAIDE (sensitivity) | CDR | 1.47E-02 | 4.35E-02 | 5.31E-02 | 1.04E-02 | 9.58E-02 |
| all | mCAIDE (sensitivity) | Total Tau | 4.56E-07 | 6.93E-06 | 1.12E-01 | 6.84E-02 | 1.55E-01 |
| all | mCAIDE (sensitivity) | MCI | 6.31E-03 | 2.36E-02 | 1.51E-01 | 4.27E-02 | 2.59E-01 |
| all | mCAIDE (sensitivity) | Cog. Impair. | 5.94E-04 | 3.25E-03 | 1.71E-01 | 7.37E-02 | 2.69E-01 |
| all | mCAIDE (sensitivity) | Dementia | 1.03E-02 | 3.41E-02 | 2.31E-01 | 5.45E-02 | 4.08E-01 |
| all | WHICAP (sensitivity) | | 1.11E-09 | 5.79E-08 | -3.22E-01 | -4.26E-01 | -2.19E-01 |
| all | WHICAP (sensitivity) | MMSE | 2.76E-06 | 2.88E-05 | -9.50E-02 | -1.35E-01 | -5.54E-02 |
| all | WHICAP (sensitivity) | Verbal Ability | 1.75E-09 | 7.17E-08 | -1.38E-01 | -1.83E-01 | -9.34E-02 |
| all | WHICAP (sensitivity) | pTau | 1.71E-08 | 4.36E-07 | -1.18E-01 | -1.58E-01 | -7.69E-02 |
| all | WHICAP (sensitivity) | Executive Function | 1.33E-08 | 3.93E-07 | -1.31E-01 | -1.76E-01 | -8.60E-02 |
| all | WHICAP (sensitivity) | NfL | 1.45E-07 | 2.63E-06 | -1.08E-01 | -1.48E-01 | -6.79E-02 |

| | | | | | | | |
|---|---|---|---|---|---|---|---|
| all | WHICAP (sensitivity) | Memory | 7.24E-06 | 6.37E-05 | -1.04E-01 | -1.49E-01 | -5.85E-02 |
| all | WHICAP (sensitivity) | Cortical Thickness | 1.34E-01 | 2.32E-01 | 3.68E-02 | -1.14E-02 | 8.51E-02 |
| all | WHICAP (sensitivity) | Aβ42/Aβ40 | 7.75E-01 | 8.70E-01 | 6.52E-03 | -3.81E-02 | 5.12E-02 |
| all | WHICAP (sensitivity) | Hippocampal Volume | 4.45E-01 | 5.76E-01 | 1.63E-02 | -2.56E-02 | 5.82E-02 |
| all | WHICAP (sensitivity) | Total Tau | 3.21E-06 | 3.12E-05 | 1.03E-01 | 5.97E-02 | 1.46E-01 |
| all | WHICAP (sensitivity) | CDR | 1.77E-04 | 1.15E-03 | 8.54E-02 | 4.08E-02 | 1.30E-01 |
| all | WHICAP (sensitivity) | Dementia | 4.93E-03 | 1.87E-02 | 2.62E-01 | 8.17E-02 | 4.48E-01 |
| all | WHICAP (sensitivity) | Cog. Impair. | 1.11E-09 | 5.79E-08 | 3.22E-01 | 2.19E-01 | 4.26E-01 |
| all | WHICAP (sensitivity) | MCI | 3.68E-09 | 1.28E-07 | 3.52E-01 | 2.36E-01 | 4.70E-01 |

Table S6. Model performance metrics for logistic regression for diagnostic outcomes (AUC & Nagelkerke $R^2$) and linear regression for AD endophenotype outcomes ($R^2$) for demographics (age, sex, and years of education), demographics + *APOE* genotype, CRS, and CRS for sensitivity analysis using complete cases.

| **Model** | **Race** | **Outcome** | **AUC** | **Nagelkerke $R^2$** | **$R^2$** |
|---|---|---|---|---|---|
| Demographics + APOE | Black | MCI | 0.66 | 5.32E-02 | |
| Demographics + APOE | Black | Dementia | 0.77 | 1.72E-01 | |
| Demographics + APOE | Black | Cog. Impair. | 0.68 | 6.97E-02 | |
| Demographics + APOE | Black | Executive Function | | | 6.77E-02 |
| Demographics + APOE | Black | Memory | | | 1.19E-01 |
| Demographics + APOE | Black | Verbal Ability | | | 1.04E-01 |
| Demographics + APOE | Black | MMSE | | | 2.10E-01 |
| Demographics + APOE | Black | CDR | | | 7.59E-02 |
| Demographics + APOE | Black | Aβ42/Aβ40 | | | 2.64E-02 |
| Demographics + APOE | Black | Total Tau | | | 1.22E-01 |
| Demographics + APOE | Black | pTau | | | 1.62E-01 |
| Demographics + APOE | Black | NfL | | | 1.44E-01 |
| Demographics + APOE | Black | Cortical Thickness | | | 1.37E-01 |
| Demographics + APOE | Black | Hippo. Volume | | | 3.27E-01 |
| CogDRisk | Black | MCI | 0.54 | 6.13E-03 | |
| CogDRisk | Black | Dementia | 0.71 | 1.04E-01 | |
| CogDRisk | Black | Cog. Impair. | 0.57 | 1.67E-02 | |
| CogDRisk | Black | Executive Function | | | 5.00E-02 |
| CogDRisk | Black | Memory | | | 2.43E-02 |
| CogDRisk | Black | Verbal Ability | | | 4.57E-02 |
| CogDRisk | Black | MMSE | | | 8.56E-02 |
| CogDRisk | Black | CDR | | | 5.28E-02 |
| CogDRisk | Black | Aβ42/Aβ40 | | | 1.51E-02 |
| CogDRisk | Black | Total Tau | | | 7.06E-02 |
| CogDRisk | Black | pTau | | | 1.40E-01 |
| CogDRisk | Black | NfL | | | 2.36E-01 |
| CogDRisk | Black | Cortical Thickness | | | 1.64E-01 |
| CogDRisk | Black | Hippo. Volume | | | 2.54E-01 |
| CogDRisk (sensitivity) | Black | MCI | 0.52 | 1.19E-03 | |
| CogDRisk (sensitivity) | Black | Dementia | 0.62 | 1.41E-02 | |
| CogDRisk (sensitivity) | Black | Cog. Impair. | 0.53 | 2.80E-03 | |

| | | | | | |
|---|---|---|---|---|---|
| CogDRisk (sensitivity) | Black | Executive Function | | | 2.04E-03 |
| CogDRisk (sensitivity) | Black | Memory | | | 2.87E-04 |
| CogDRisk (sensitivity) | Black | Verbal Ability | | | 2.71E-03 |
| CogDRisk (sensitivity) | Black | MMSE | | | 1.27E-03 |
| CogDRisk (sensitivity) | Black | CDR | | | 1.57E-02 |
| CogDRisk (sensitivity) | Black | Aβ42/Aβ40 | | | 1.47E-02 |
| CogDRisk (sensitivity) | Black | Total Tau | | | 6.05E-02 |
| CogDRisk (sensitivity) | Black | pTau | | | 8.17E-02 |
| CogDRisk (sensitivity) | Black | NfL | | | 1.18E-01 |
| CogDRisk (sensitivity) | Black | Cortical Thickness | | | 2.40E-02 |
| CogDRisk (sensitivity) | Black | Hippo. Volume | | | 1.45E-01 |
| LIBRA | Black | MCI | 0.56 | 1.15E-02 | |
| LIBRA | Black | Dementia | 0.64 | 3.96E-02 | |
| LIBRA | Black | Cog. Impair. | 0.58 | 1.64E-02 | |
| LIBRA | Black | Executive Function | | | 1.84E-02 |
| LIBRA | Black | Memory | | | 4.78E-03 |
| LIBRA | Black | Verbal Ability | | | 1.31E-02 |
| LIBRA | Black | MMSE | | | 9.15E-03 |
| LIBRA | Black | CDR | | | 3.12E-02 |
| LIBRA | Black | Aβ42/Aβ40 | | | 2.49E-02 |
| LIBRA | Black | Total Tau | | | 2.64E-02 |
| LIBRA | Black | pTau | | | 1.22E-02 |
| LIBRA | Black | NfL | | | 1.00E-02 |
| LIBRA | Black | Cortical Thickness | | | 2.23E-02 |
| LIBRA | Black | Hippo. Volume | | | 1.56E-01 |
| LIBRA (sensitivity) | Black | MCI | 0.59 | 2.17E-02 | |
| LIBRA (sensitivity) | Black | Dementia | 0.72 | 1.09E-01 | |
| LIBRA (sensitivity) | Black | Cog. Impair. | 0.61 | 3.35E-02 | |
| LIBRA (sensitivity) | Black | Executive Function | | | 5.64E-02 |
| LIBRA (sensitivity) | Black | Memory | | | 2.46E-02 |
| LIBRA (sensitivity) | Black | Verbal Ability | | | 4.18E-02 |
| LIBRA (sensitivity) | Black | MMSE | | | 5.35E-02 |
| LIBRA (sensitivity) | Black | CDR | | | 6.96E-02 |
| LIBRA (sensitivity) | Black | Aβ42/Aβ40 | | | 2.77E-02 |
| LIBRA (sensitivity) | Black | Total Tau | | | 3.41E-02 |
| LIBRA (sensitivity) | Black | pTau | | | 6.09E-02 |

| | | | | | |
|---|---|---|---|---|---|
| LIBRA (sensitivity) | Black | NfL | | | 6.70E-02 |
| LIBRA (sensitivity) | Black | Cortical Thickness | | | 8.60E-02 |
| LIBRA (sensitivity) | Black | Hippo. Volume | | | 2.11E-01 |
| mCAIDE | Black | MCI | 0.56 | 8.75E-03 | |
| mCAIDE | Black | Dementia | 0.62 | 2.39E-02 | |
| mCAIDE | Black | Cog. Impair. | 0.57 | 1.20E-02 | |
| mCAIDE | Black | Executive Function | | | 1.79E-02 |
| mCAIDE | Black | Memory | | | 1.76E-02 |
| mCAIDE | Black | Verbal Ability | | | 3.11E-02 |
| mCAIDE | Black | MMSE | | | 1.87E-02 |
| mCAIDE | Black | CDR | | | 1.13E-02 |
| mCAIDE | Black | Aβ42/Aβ40 | | | 7.04E-03 |
| mCAIDE | Black | Total Tau | | | 4.01E-02 |
| mCAIDE | Black | pTau | | | 1.02E-01 |
| mCAIDE | Black | NfL | | | 1.30E-01 |
| mCAIDE | Black | Cortical Thickness | | | 6.05E-02 |
| mCAIDE | Black | Hippo. Volume | | | 1.88E-01 |
| mCAIDE (sensitivity) | Black | MCI | 0.52 | 8.93E-04 | |
| mCAIDE (sensitivity) | Black | Dementia | 0.50 | 5.99E-05 | |
| mCAIDE (sensitivity) | Black | Cog. Impair. | 0.52 | 6.90E-04 | |
| mCAIDE (sensitivity) | Black | Executive Function | | | 1.77E-03 |
| mCAIDE (sensitivity) | Black | Memory | | | 2.51E-04 |
| mCAIDE (sensitivity) | Black | Verbal Ability | | | 6.97E-03 |
| mCAIDE (sensitivity) | Black | MMSE | | | 3.02E-04 |
| mCAIDE (sensitivity) | Black | CDR | | | 7.78E-05 |
| mCAIDE (sensitivity) | Black | Aβ42/Aβ40 | | | 6.28E-03 |
| mCAIDE (sensitivity) | Black | Total Tau | | | 4.71E-02 |
| mCAIDE (sensitivity) | Black | pTau | | | 8.55E-02 |
| mCAIDE (sensitivity) | Black | NfL | | | 1.20E-01 |
| mCAIDE (sensitivity) | Black | Cortical Thickness | | | 2.30E-02 |
| mCAIDE (sensitivity) | Black | Hippo. Volume | | | 1.49E-01 |
| WHICAP | Black | MCI | 0.54 | 8.48E-03 | |
| WHICAP | Black | Dementia | 0.63 | 6.33E-02 | |
| WHICAP | Black | Cog. Impair. | 0.56 | 1.57E-02 | |
| WHICAP | Black | Executive Function | | | 2.48E-02 |
| WHICAP | Black | Memory | | | 8.31E-03 |

| | | | | | |
|---|---|---|---|---|---|
| WHICAP | Black | Verbal Ability | | | 4.48E-02 |
| WHICAP | Black | MMSE | | | 4.54E-02 |
| WHICAP | Black | CDR | | | 3.54E-02 |
| WHICAP | Black | Aβ42/Aβ40 | | | 2.10E-02 |
| WHICAP | Black | Total Tau | | | 4.64E-02 |
| WHICAP | Black | pTau | | | 9.84E-02 |
| WHICAP | Black | NfL | | | 1.37E-01 |
| WHICAP | Black | Cortical Thickness | | | 5.47E-02 |
| WHICAP | Black | Hippo. Volume | | | 2.01E-01 |
| WHICAP (sensitivity) | Black | MCI | 0.51 | 2.59E-03 | |
| WHICAP (sensitivity) | Black | Dementia | 0.50 | 6.01E-04 | |
| WHICAP (sensitivity) | Black | Cog. Impair. | 0.51 | 1.18E-03 | |
| WHICAP (sensitivity) | Black | Executive Function | | | 1.59E-05 |
| WHICAP (sensitivity) | Black | Memory | | | 4.69E-03 |
| WHICAP (sensitivity) | Black | Verbal Ability | | | 7.83E-04 |
| WHICAP (sensitivity) | Black | MMSE | | | 2.07E-03 |
| WHICAP (sensitivity) | Black | CDR | | | 1.81E-04 |
| WHICAP (sensitivity) | Black | Aβ42/Aβ40 | | | 1.65E-02 |
| WHICAP (sensitivity) | Black | Total Tau | | | 3.79E-02 |
| WHICAP (sensitivity) | Black | pTau | | | 9.04E-02 |
| WHICAP (sensitivity) | Black | NfL | | | 1.27E-01 |
| WHICAP (sensitivity) | Black | Cortical Thickness | | | 2.20E-02 |
| WHICAP (sensitivity) | Black | Hippo. Volume | | | 1.43E-01 |
| Demographics + APOE | Hispanic | MCI | 0.56 | 6.40E-03 | |
| Demographics + APOE | Hispanic | Dementia | 0.73 | 1.12E-01 | |
| Demographics + APOE | Hispanic | Cog. Impair. | 0.60 | 2.17E-02 | |
| Demographics + APOE | Hispanic | Executive Function | | | 2.09E-02 |
| Demographics + APOE | Hispanic | Memory | | | 5.35E-02 |
| Demographics + APOE | Hispanic | Verbal Ability | | | 1.70E-02 |
| Demographics + APOE | Hispanic | MMSE | | | 3.43E-01 |
| Demographics + APOE | Hispanic | CDR | | | 6.29E-02 |
| Demographics + APOE | Hispanic | Aβ42/Aβ40 | | | 4.01E-02 |
| Demographics + APOE | Hispanic | Total Tau | | | 1.58E-01 |
| Demographics + APOE | Hispanic | pTau | | | 2.00E-01 |
| Demographics + APOE | Hispanic | NfL | | | 2.67E-01 |
| Demographics + APOE | Hispanic | Cortical Thickness | | | 1.31E-01 |

| | | | | | |
|---|---|---|---|---|---|
| Demographics + APOE | Hispanic | Hippo. Volume | | | 3.33E-01 |
| CogDRisk | Hispanic | MCI | 0.56 | 7.48E-03 | |
| CogDRisk | Hispanic | Dementia | 0.63 | 4.18E-02 | |
| CogDRisk | Hispanic | Cog. Impair. | 0.58 | 1.57E-02 | |
| CogDRisk | Hispanic | Executive Function | | | 1.59E-02 |
| CogDRisk | Hispanic | Memory | | | 3.61E-03 |
| CogDRisk | Hispanic | Verbal Ability | | | 1.04E-02 |
| CogDRisk | Hispanic | MMSE | | | 2.49E-01 |
| CogDRisk | Hispanic | CDR | | | 3.88E-02 |
| CogDRisk | Hispanic | Aβ42/Aβ40 | | | 2.59E-02 |
| CogDRisk | Hispanic | Total Tau | | | 1.18E-01 |
| CogDRisk | Hispanic | pTau | | | 1.83E-01 |
| CogDRisk | Hispanic | NfL | | | 3.03E-01 |
| CogDRisk | Hispanic | Cortical Thickness | | | 9.80E-02 |
| CogDRisk | Hispanic | Hippo. Volume | | | 2.81E-01 |
| CogDRisk (sensitivity) | Hispanic | MCI | 0.54 | 3.56E-03 | |
| CogDRisk (sensitivity) | Hispanic | Dementia | 0.58 | 8.17E-03 | |
| CogDRisk (sensitivity) | Hispanic | Cog. Impair. | 0.55 | 5.05E-03 | |
| CogDRisk (sensitivity) | Hispanic | Executive Function | | | 1.12E-02 |
| CogDRisk (sensitivity) | Hispanic | Memory | | | 2.54E-04 |
| CogDRisk (sensitivity) | Hispanic | Verbal Ability | | | 1.02E-02 |
| CogDRisk (sensitivity) | Hispanic | MMSE | | | 1.62E-01 |
| CogDRisk (sensitivity) | Hispanic | CDR | | | 1.64E-02 |
| CogDRisk (sensitivity) | Hispanic | Aβ42/Aβ40 | | | 9.05E-03 |
| CogDRisk (sensitivity) | Hispanic | Total Tau | | | 1.12E-01 |
| CogDRisk (sensitivity) | Hispanic | pTau | | | 1.69E-01 |
| CogDRisk (sensitivity) | Hispanic | NfL | | | 2.06E-01 |
| CogDRisk (sensitivity) | Hispanic | Cortical Thickness | | | 6.03E-03 |
| CogDRisk (sensitivity) | Hispanic | Hippo. Volume | | | 1.64E-01 |
| LIBRA | Hispanic | MCI | 0.54 | 4.52E-03 | |
| LIBRA | Hispanic | Dementia | 0.62 | 2.26E-02 | |
| LIBRA | Hispanic | Cog. Impair. | 0.56 | 8.76E-03 | |
| LIBRA | Hispanic | Executive Function | | | 1.63E-02 |
| LIBRA | Hispanic | Memory | | | 6.72E-03 |
| LIBRA | Hispanic | Verbal Ability | | | 2.16E-02 |
| LIBRA | Hispanic | MMSE | | | 1.66E-01 |

| | | | | | |
|---|---|---|---|---|---|
| LIBRA | Hispanic | CDR | | | 2.06E-02 |
| LIBRA | Hispanic | Aβ42/Aβ40 | | | 2.30E-03 |
| LIBRA | Hispanic | Total Tau | | | 2.81E-02 |
| LIBRA | Hispanic | pTau | | | 2.26E-02 |
| LIBRA | Hispanic | NfL | | | 7.50E-02 |
| LIBRA | Hispanic | Cortical Thickness | | | 7.16E-03 |
| LIBRA | Hispanic | Hippo. Volume | | | 1.71E-01 |
| LIBRA (sensitivity) | Hispanic | MCI | 0.55 | 5.42E-03 | |
| LIBRA (sensitivity) | Hispanic | Dementia | 0.67 | 5.21E-02 | |
| LIBRA (sensitivity) | Hispanic | Cog. Impair. | 0.58 | 1.48E-02 | |
| LIBRA (sensitivity) | Hispanic | Executive Function | | | 2.33E-02 |
| LIBRA (sensitivity) | Hispanic | Memory | | | 6.70E-03 |
| LIBRA (sensitivity) | Hispanic | Verbal Ability | | | 2.57E-02 |
| LIBRA (sensitivity) | Hispanic | MMSE | | | 2.70E-01 |
| LIBRA (sensitivity) | Hispanic | CDR | | | 4.48E-02 |
| LIBRA (sensitivity) | Hispanic | Aβ42/Aβ40 | | | 1.29E-02 |
| LIBRA (sensitivity) | Hispanic | Total Tau | | | 2.92E-02 |
| LIBRA (sensitivity) | Hispanic | pTau | | | 5.90E-02 |
| LIBRA (sensitivity) | Hispanic | NfL | | | 1.92E-01 |
| LIBRA (sensitivity) | Hispanic | Cortical Thickness | | | 5.11E-02 |
| LIBRA (sensitivity) | Hispanic | Hippo. Volume | | | 2.21E-01 |
| mCAIDE | Hispanic | MCI | 0.53 | 2.19E-03 | |
| mCAIDE | Hispanic | Dementia | 0.59 | 1.58E-02 | |
| mCAIDE | Hispanic | Cog. Impair. | 0.55 | 5.11E-03 | |
| mCAIDE | Hispanic | Executive Function | | | 7.79E-03 |
| mCAIDE | Hispanic | Memory | | | 3.24E-03 |
| mCAIDE | Hispanic | Verbal Ability | | | 1.74E-02 |
| mCAIDE | Hispanic | MMSE | | | 1.83E-01 |
| mCAIDE | Hispanic | CDR | | | 1.16E-02 |
| mCAIDE | Hispanic | Aβ42/Aβ40 | | | 1.27E-02 |
| mCAIDE | Hispanic | Total Tau | | | 9.23E-02 |
| mCAIDE | Hispanic | pTau | | | 1.70E-01 |
| mCAIDE | Hispanic | NfL | | | 1.87E-01 |
| mCAIDE | Hispanic | Cortical Thickness | | | 4.17E-02 |
| mCAIDE | Hispanic | Hippo. Volume | | | 1.92E-01 |
| mCAIDE (sensitivity) | Hispanic | MCI | 0.50 | 1.49E-04 | |

| | | | | | |
|---|---|---|---|---|---|
| mCAIDE (sensitivity) | Hispanic | Dementia | 0.54 | 3.14E-03 | |
| mCAIDE (sensitivity) | Hispanic | Cog. Impair. | 0.51 | 6.41E-04 | |
| mCAIDE (sensitivity) | Hispanic | Executive Function | | | 6.89E-03 |
| mCAIDE (sensitivity) | Hispanic | Memory | | | 3.98E-04 |
| mCAIDE (sensitivity) | Hispanic | Verbal Ability | | | 1.52E-02 |
| mCAIDE (sensitivity) | Hispanic | MMSE | | | 1.57E-01 |
| mCAIDE (sensitivity) | Hispanic | CDR | | | 6.49E-03 |
| mCAIDE (sensitivity) | Hispanic | Aβ42/Aβ40 | | | 7.50E-03 |
| mCAIDE (sensitivity) | Hispanic | Total Tau | | | 9.89E-02 |
| mCAIDE (sensitivity) | Hispanic | pTau | | | 1.69E-01 |
| mCAIDE (sensitivity) | Hispanic | NfL | | | 1.73E-01 |
| mCAIDE (sensitivity) | Hispanic | Cortical Thickness | | | 9.81E-03 |
| mCAIDE (sensitivity) | Hispanic | Hippo. Volume | | | 1.56E-01 |
| WHICAP | Hispanic | MCI | 0.52 | 1.43E-03 | |
| WHICAP | Hispanic | Dementia | 0.65 | 4.26E-02 | |
| WHICAP | Hispanic | Cog. Impair. | 0.55 | 7.87E-03 | |
| WHICAP | Hispanic | Executive Function | | | 1.09E-02 |
| WHICAP | Hispanic | Memory | | | 5.37E-03 |
| WHICAP | Hispanic | Verbal Ability | | | 1.37E-02 |
| WHICAP | Hispanic | MMSE | | | 2.58E-01 |
| WHICAP | Hispanic | CDR | | | 2.81E-02 |
| WHICAP | Hispanic | Aβ42/Aβ40 | | | 1.58E-02 |
| WHICAP | Hispanic | Total Tau | | | 9.52E-02 |
| WHICAP | Hispanic | pTau | | | 1.73E-01 |
| WHICAP | Hispanic | NfL | | | 2.33E-01 |
| WHICAP | Hispanic | Cortical Thickness | | | 5.02E-02 |
| WHICAP | Hispanic | Hippo. Volume | | | 1.99E-01 |
| WHICAP (sensitivity) | Hispanic | MCI | 0.51 | 1.22E-05 | |
| WHICAP (sensitivity) | Hispanic | Dementia | 0.54 | 5.33E-04 | |
| WHICAP (sensitivity) | Hispanic | Cog. Impair. | 0.51 | 8.74E-05 | |
| WHICAP (sensitivity) | Hispanic | Executive Function | | | 6.30E-03 |
| WHICAP (sensitivity) | Hispanic | Memory | | | 2.75E-04 |
| WHICAP (sensitivity) | Hispanic | Verbal Ability | | | 1.17E-02 |
| WHICAP (sensitivity) | Hispanic | MMSE | | | 1.55E-01 |
| WHICAP (sensitivity) | Hispanic | CDR | | | 6.45E-03 |
| WHICAP (sensitivity) | Hispanic | Aβ42/Aβ40 | | | 2.65E-03 |

| | | | | | |
|---|---|---|---|---|---|
| WHICAP (sensitivity) | Hispanic | Total Tau | | | 1.00E-01 |
| WHICAP (sensitivity) | Hispanic | pTau | | | 1.68E-01 |
| WHICAP (sensitivity) | Hispanic | NfL | | | 1.76E-01 |
| WHICAP (sensitivity) | Hispanic | Cortical Thickness | | | 2.75E-03 |
| WHICAP (sensitivity) | Hispanic | Hippo. Volume | | | 1.57E-01 |
| Demographics + APOE | NHW | MCI | 0.63 | 3.15E-02 | |
| Demographics + APOE | NHW | Dementia | 0.75 | 1.01E-01 | |
| Demographics + APOE | NHW | Cog. Impair. | 0.66 | 4.78E-02 | |
| Demographics + APOE | NHW | Executive Function | | | 4.96E-02 |
| Demographics + APOE | NHW | Memory | | | 1.41E-01 |
| Demographics + APOE | NHW | Verbal Ability | | | 3.44E-02 |
| Demographics + APOE | NHW | MMSE | | | 8.31E-02 |
| Demographics + APOE | NHW | CDR | | | 3.71E-02 |
| Demographics + APOE | NHW | Aβ42/Aβ40 | | | 8.68E-02 |
| Demographics + APOE | NHW | Total Tau | | | 1.19E-01 |
| Demographics + APOE | NHW | pTau | | | 2.30E-01 |
| Demographics + APOE | NHW | NfL | | | 3.65E-01 |
| Demographics + APOE | NHW | Cortical Thickness | | | 1.91E-01 |
| Demographics + APOE | NHW | Hippo. Volume | | | 3.56E-01 |
| CogDRisk | NHW | MCI | 0.57 | 1.37E-02 | |
| CogDRisk | NHW | Dementia | 0.65 | 5.00E-02 | |
| CogDRisk | NHW | Cog. Impair. | 0.60 | 2.45E-02 | |
| CogDRisk | NHW | Executive Function | | | 4.51E-02 |
| CogDRisk | NHW | Memory | | | 3.40E-02 |
| CogDRisk | NHW | Verbal Ability | | | 2.17E-02 |
| CogDRisk | NHW | MMSE | | | 4.79E-02 |
| CogDRisk | NHW | CDR | | | 2.28E-02 |
| CogDRisk | NHW | Aβ42/Aβ40 | | | 2.81E-02 |
| CogDRisk | NHW | Total Tau | | | 6.12E-02 |
| CogDRisk | NHW | pTau | | | 1.52E-01 |
| CogDRisk | NHW | NfL | | | 3.62E-01 |
| CogDRisk | NHW | Cortical Thickness | | | 1.38E-01 |
| CogDRisk | NHW | Hippo. Volume | | | 3.08E-01 |
| CogDRisk (sensitivity) | NHW | MCI | 0.57 | 1.02E-02 | |
| CogDRisk (sensitivity) | NHW | Dementia | 0.58 | 1.40E-02 | |
| CogDRisk (sensitivity) | NHW | Cog. Impair. | 0.58 | 1.25E-02 | |

| | | | | | |
|---|---|---|---|---|---|
| CogDRisk (sensitivity) | NHW | Executive Function | | | 2.33E-03 |
| CogDRisk (sensitivity) | NHW | Memory | | | 1.93E-04 |
| CogDRisk (sensitivity) | NHW | Verbal Ability | | | 3.58E-03 |
| CogDRisk (sensitivity) | NHW | MMSE | | | 7.82E-03 |
| CogDRisk (sensitivity) | NHW | CDR | | | 8.31E-03 |
| CogDRisk (sensitivity) | NHW | Aβ42/Aβ40 | | | 1.55E-02 |
| CogDRisk (sensitivity) | NHW | Total Tau | | | 5.97E-02 |
| CogDRisk (sensitivity) | NHW | pTau | | | 8.60E-02 |
| CogDRisk (sensitivity) | NHW | NfL | | | 2.22E-01 |
| CogDRisk (sensitivity) | NHW | Cortical Thickness | | | 1.00E-03 |
| CogDRisk (sensitivity) | NHW | Hippo. Volume | | | 1.00E-01 |
| LIBRA | NHW | MCI | 0.58 | 1.48E-02 | |
| LIBRA | NHW | Dementia | 0.57 | 1.18E-02 | |
| LIBRA | NHW | Cog. Impair. | 0.58 | 1.52E-02 | |
| LIBRA | NHW | Executive Function | | | 5.01E-03 |
| LIBRA | NHW | Memory | | | 4.65E-03 |
| LIBRA | NHW | Verbal Ability | | | 9.47E-03 |
| LIBRA | NHW | MMSE | | | 9.30E-03 |
| LIBRA | NHW | CDR | | | 6.58E-03 |
| LIBRA | NHW | Aβ42/Aβ40 | | | 8.42E-03 |
| LIBRA | NHW | Total Tau | | | 1.30E-02 |
| LIBRA | NHW | pTau | | | 5.09E-04 |
| LIBRA | NHW | NfL | | | 6.55E-03 |
| LIBRA | NHW | Cortical Thickness | | | 2.36E-03 |
| LIBRA | NHW | Hippo. Volume | | | 1.05E-01 |
| LIBRA (sensitivity) | NHW | MCI | 0.59 | 1.75E-02 | |
| LIBRA (sensitivity) | NHW | Dementia | 0.63 | 3.64E-02 | |
| LIBRA (sensitivity) | NHW | Cog. Impair. | 0.60 | 2.47E-02 | |
| LIBRA (sensitivity) | NHW | Executive Function | | | 2.96E-02 |
| LIBRA (sensitivity) | NHW | Memory | | | 2.29E-02 |
| LIBRA (sensitivity) | NHW | Verbal Ability | | | 2.35E-02 |
| LIBRA (sensitivity) | NHW | MMSE | | | 3.10E-02 |
| LIBRA (sensitivity) | NHW | CDR | | | 1.75E-02 |
| LIBRA (sensitivity) | NHW | Aβ42/Aβ40 | | | 2.20E-04 |
| LIBRA (sensitivity) | NHW | Total Tau | | | 1.31E-02 |
| LIBRA (sensitivity) | NHW | pTau | | | 4.08E-02 |

| | | | | | |
|---|---|---|---|---|---|
| LIBRA (sensitivity) | NHW | NfL | | | 1.11E-01 |
| LIBRA (sensitivity) | NHW | Cortical Thickness | | | 5.99E-02 |
| LIBRA (sensitivity) | NHW | Hippo. Volume | | | 1.95E-01 |
| mCAIDE | NHW | MCI | 0.59 | 1.18E-02 | |
| mCAIDE | NHW | Dementia | 0.59 | 1.51E-02 | |
| mCAIDE | NHW | Cog. Impair. | 0.59 | 1.44E-02 | |
| mCAIDE | NHW | Executive Function | | | 9.87E-03 |
| mCAIDE | NHW | Memory | | | 2.67E-02 |
| mCAIDE | NHW | Verbal Ability | | | 1.57E-02 |
| mCAIDE | NHW | MMSE | | | 2.88E-02 |
| mCAIDE | NHW | CDR | | | 7.85E-03 |
| mCAIDE | NHW | Aβ42/Aβ40 | | | 7.45E-03 |
| mCAIDE | NHW | Total Tau | | | 4.32E-02 |
| mCAIDE | NHW | pTau | | | 8.78E-02 |
| mCAIDE | NHW | NfL | | | 2.24E-01 |
| mCAIDE | NHW | Cortical Thickness | | | 2.89E-02 |
| mCAIDE | NHW | Hippo. Volume | | | 1.48E-01 |
| mCAIDE (sensitivity) | NHW | MCI | 0.56 | 4.85E-03 | |
| mCAIDE (sensitivity) | NHW | Dementia | 0.55 | 3.17E-03 | |
| mCAIDE (sensitivity) | NHW | Cog. Impair. | 0.55 | 4.81E-03 | |
| mCAIDE (sensitivity) | NHW | Executive Function | | | 1.15E-03 |
| mCAIDE (sensitivity) | NHW | Memory | | | 2.92E-03 |
| mCAIDE (sensitivity) | NHW | Verbal Ability | | | 6.25E-03 |
| mCAIDE (sensitivity) | NHW | MMSE | | | 1.01E-02 |
| mCAIDE (sensitivity) | NHW | CDR | | | 1.83E-03 |
| mCAIDE (sensitivity) | NHW | Aβ42/Aβ40 | | | 1.20E-02 |
| mCAIDE (sensitivity) | NHW | Total Tau | | | 4.88E-02 |
| mCAIDE (sensitivity) | NHW | pTau | | | 8.54E-02 |
| mCAIDE (sensitivity) | NHW | NfL | | | 2.22E-01 |
| mCAIDE (sensitivity) | NHW | Cortical Thickness | | | 7.49E-04 |
| mCAIDE (sensitivity) | NHW | Hippo. Volume | | | 1.03E-01 |
| WHICAP | NHW | MCI | 0.57 | 1.15E-02 | |
| WHICAP | NHW | Dementia | 0.59 | 1.50E-02 | |
| WHICAP | NHW | Cog. Impair. | 0.57 | 1.37E-02 | |
| WHICAP | NHW | Executive Function | | | 2.74E-02 |
| WHICAP | NHW | Memory | | | 2.16E-02 |

| | | | | | |
|---|---|---|---|---|---|
| WHICAP | NHW | Verbal Ability | | | 2.62E-02 |
| WHICAP | NHW | MMSE | | | 3.29E-02 |
| WHICAP | NHW | CDR | | | 7.85E-03 |
| WHICAP | NHW | Aβ42/Aβ40 | | | 7.57E-03 |
| WHICAP | NHW | Total Tau | | | 4.49E-02 |
| WHICAP | NHW | pTau | | | 9.98E-02 |
| WHICAP | NHW | NfL | | | 2.54E-01 |
| WHICAP | NHW | Cortical Thickness | | | 9.44E-02 |
| WHICAP | NHW | Hippo. Volume | | | 2.39E-01 |
| WHICAP (sensitivity) | NHW | MCI | 0.54 | 2.42E-03 | |
| WHICAP (sensitivity) | NHW | Dementia | 0.52 | 1.51E-03 | |
| WHICAP (sensitivity) | NHW | Cog. Impair. | 0.52 | 4.93E-04 | |
| WHICAP (sensitivity) | NHW | Executive Function | | | 7.41E-04 |
| WHICAP (sensitivity) | NHW | Memory | | | 3.39E-04 |
| WHICAP (sensitivity) | NHW | Verbal Ability | | | 2.37E-03 |
| WHICAP (sensitivity) | NHW | MMSE | | | 3.22E-03 |
| WHICAP (sensitivity) | NHW | CDR | | | 1.29E-03 |
| WHICAP (sensitivity) | NHW | Aβ42/Aβ40 | | | 1.47E-02 |
| WHICAP (sensitivity) | NHW | Total Tau | | | 4.69E-02 |
| WHICAP (sensitivity) | NHW | pTau | | | 9.91E-02 |
| WHICAP (sensitivity) | NHW | NfL | | | 2.41E-01 |
| WHICAP (sensitivity) | NHW | Cortical Thickness | | | 2.58E-03 |
| WHICAP (sensitivity) | NHW | Hippo. Volume | | | 9.85E-02 |
| Demographics + APOE | all | MCI | 0.58 | 1.15E-02 | |
| Demographics + APOE | all | Dementia | 0.73 | 9.36E-02 | |
| Demographics + APOE | all | Cog. Impair. | 0.61 | 2.61E-02 | |
| Demographics + APOE | all | Executive Function | | | 2.17E-02 |
| Demographics + APOE | all | Memory | | | 7.69E-02 |
| Demographics + APOE | all | Verbal Ability | | | 2.38E-02 |
| Demographics + APOE | all | MMSE | | | 3.46E-01 |
| Demographics + APOE | all | CDR | | | 4.63E-02 |
| Demographics + APOE | all | Aβ42/Aβ40 | | | 4.52E-02 |
| Demographics + APOE | all | Total Tau | | | 1.16E-01 |
| Demographics + APOE | all | pTau | | | 2.17E-01 |
| Demographics + APOE | all | NfL | | | 2.96E-01 |
| Demographics + APOE | all | Cortical Thickness | | | 1.57E-01 |

| | | | | | |
|---|---|---|---|---|---|
| Demographics + APOE | all | Hippo. Volume | | | 3.24E-01 |
| CogDRisk | all | MCI | 0.53 | 2.76E-03 | |
| CogDRisk | all | Dementia | 0.65 | 4.57E-02 | |
| CogDRisk | all | Cog. Impair. | 0.56 | 9.92E-03 | |
| CogDRisk | all | Executive Function | | | 1.73E-02 |
| CogDRisk | all | Memory | | | 8.47E-03 |
| CogDRisk | all | Verbal Ability | | | 1.11E-02 |
| CogDRisk | all | MMSE | | | 2.40E-01 |
| CogDRisk | all | CDR | | | 2.46E-02 |
| CogDRisk | all | Aβ42/Aβ40 | | | 1.92E-02 |
| CogDRisk | all | Total Tau | | | 6.56E-02 |
| CogDRisk | all | pTau | | | 1.67E-01 |
| CogDRisk | all | NfL | | | 3.16E-01 |
| CogDRisk | all | Cortical Thickness | | | 1.27E-01 |
| CogDRisk | all | Hippo. Volume | | | 2.58E-01 |
| CogDRisk (sensitivity) | all | MCI | 0.55 | 6.20E-03 | |
| CogDRisk (sensitivity) | all | Dementia | 0.59 | 1.35E-02 | |
| CogDRisk (sensitivity) | all | Cog. Impair. | 0.56 | 8.58E-03 | |
| CogDRisk (sensitivity) | all | Executive Function | | | 6.24E-03 |
| CogDRisk (sensitivity) | all | Memory | | | 7.10E-04 |
| CogDRisk (sensitivity) | all | Verbal Ability | | | 4.90E-03 |
| CogDRisk (sensitivity) | all | MMSE | | | 2.00E-01 |
| CogDRisk (sensitivity) | all | CDR | | | 1.30E-02 |
| CogDRisk (sensitivity) | all | Aβ42/Aβ40 | | | 2.13E-03 |
| CogDRisk (sensitivity) | all | Total Tau | | | 6.41E-02 |
| CogDRisk (sensitivity) | all | pTau | | | 1.16E-01 |
| CogDRisk (sensitivity) | all | NfL | | | 1.61E-01 |
| CogDRisk (sensitivity) | all | Cortical Thickness | | | 3.80E-03 |
| CogDRisk (sensitivity) | all | Hippo. Volume | | | 1.12E-01 |
| LIBRA | all | MCI | 0.58 | 1.38E-02 | |
| LIBRA | all | Dementia | 0.62 | 2.56E-02 | |
| LIBRA | all | Cog. Impair. | 0.59 | 1.79E-02 | |
| LIBRA | all | Executive Function | | | 1.71E-02 |
| LIBRA | all | Memory | | | 1.12E-02 |
| LIBRA | all | Verbal Ability | | | 1.63E-02 |
| LIBRA | all | MMSE | | | 2.05E-01 |

| | | | | | |
|---|---|---|---|---|---|
| LIBRA | all | CDR | | | 1.86E-02 |
| LIBRA | all | Aβ42/Aβ40 | | | 2.56E-04 |
| LIBRA | all | Total Tau | | | 2.16E-02 |
| LIBRA | all | pTau | | | 2.75E-03 |
| LIBRA | all | NfL | | | 1.42E-02 |
| LIBRA | all | Cortical Thickness | | | 5.24E-03 |
| LIBRA | all | Hippo. Volume | | | 1.18E-01 |
| LIBRA (sensitivity) | all | MCI | 0.57 | 1.12E-02 | |
| LIBRA (sensitivity) | all | Dementia | 0.66 | 5.44E-02 | |
| LIBRA (sensitivity) | all | Cog. Impair. | 0.59 | 2.08E-02 | |
| LIBRA (sensitivity) | all | Executive Function | | | 2.91E-02 |
| LIBRA (sensitivity) | all | Memory | | | 1.69E-02 |
| LIBRA (sensitivity) | all | Verbal Ability | | | 2.43E-02 |
| LIBRA (sensitivity) | all | MMSE | | | 2.53E-01 |
| LIBRA (sensitivity) | all | CDR | | | 3.37E-02 |
| LIBRA (sensitivity) | all | Aβ42/Aβ40 | | | 6.95E-03 |
| LIBRA (sensitivity) | all | Total Tau | | | 2.26E-02 |
| LIBRA (sensitivity) | all | pTau | | | 4.42E-02 |
| LIBRA (sensitivity) | all | NfL | | | 1.14E-01 |
| LIBRA (sensitivity) | all | Cortical Thickness | | | 5.90E-02 |
| LIBRA (sensitivity) | all | Hippo. Volume | | | 1.87E-01 |
| mCAIDE | all | MCI | 0.56 | 7.35E-03 | |
| mCAIDE | all | Dementia | 0.60 | 1.89E-02 | |
| mCAIDE | all | Cog. Impair. | 0.57 | 1.09E-02 | |
| mCAIDE | all | Executive Function | | | 1.11E-02 |
| mCAIDE | all | Memory | | | 1.59E-02 |
| mCAIDE | all | Verbal Ability | | | 1.82E-02 |
| mCAIDE | all | MMSE | | | 2.16E-01 |
| mCAIDE | all | CDR | | | 8.99E-03 |
| mCAIDE | all | Aβ42/Aβ40 | | | 4.07E-03 |
| mCAIDE | all | Total Tau | | | 4.24E-02 |
| mCAIDE | all | pTau | | | 1.19E-01 |
| mCAIDE | all | NfL | | | 1.66E-01 |
| mCAIDE | all | Cortical Thickness | | | 3.55E-02 |
| mCAIDE | all | Hippo. Volume | | | 1.43E-01 |
| mCAIDE (sensitivity) | all | MCI | 0.54 | 3.73E-03 | |

| | | | | | |
|---|---|---|---|---|---|
| mCAIDE (sensitivity) | all | Dementia | 0.54 | 3.55E-03 | |
| mCAIDE (sensitivity) | all | Cog. Impair. | 0.54 | 4.05E-03 | |
| mCAIDE (sensitivity) | all | Executive Function | | | 4.94E-03 |
| mCAIDE (sensitivity) | all | Memory | | | 2.37E-03 |
| mCAIDE (sensitivity) | all | Verbal Ability | | | 1.00E-02 |
| mCAIDE (sensitivity) | all | MMSE | | | 1.96E-01 |
| mCAIDE (sensitivity) | all | CDR | | | 2.51E-03 |
| mCAIDE (sensitivity) | all | Aβ42/Aβ40 | | | 1.98E-03 |
| mCAIDE (sensitivity) | all | Total Tau | | | 4.94E-02 |
| mCAIDE (sensitivity) | all | pTau | | | 1.17E-01 |
| mCAIDE (sensitivity) | all | NfL | | | 1.56E-01 |
| mCAIDE (sensitivity) | all | Cortical Thickness | | | 5.14E-03 |
| mCAIDE (sensitivity) | all | Hippo. Volume | | | 1.13E-01 |
| WHICAP | all | MCI | 0.57 | 1.09E-02 | |
| WHICAP | all | Dementia | 0.63 | 3.45E-02 | |
| WHICAP | all | Cog. Impair. | 0.59 | 1.72E-02 | |
| WHICAP | all | Executive Function | | | 3.44E-02 |
| WHICAP | all | Memory | | | 2.42E-02 |
| WHICAP | all | Verbal Ability | | | 3.06E-02 |
| WHICAP | all | MMSE | | | 2.65E-01 |
| WHICAP | all | CDR | | | 2.41E-02 |
| WHICAP | all | Aβ42/Aβ40 | | | 1.14E-02 |
| WHICAP | all | Total Tau | | | 4.84E-02 |
| WHICAP | all | pTau | | | 1.18E-01 |
| WHICAP | all | NfL | | | 1.84E-01 |
| WHICAP | all | Cortical Thickness | | | 4.38E-02 |
| WHICAP | all | Hippo. Volume | | | 1.47E-01 |
| WHICAP (sensitivity) | all | MCI | 0.59 | 1.75E-02 | |
| WHICAP (sensitivity) | all | Dementia | 0.55 | 2.65E-03 | |
| WHICAP (sensitivity) | all | Cog. Impair. | 0.58 | 1.31E-02 | |
| WHICAP (sensitivity) | all | Executive Function | | | 1.74E-02 |
| WHICAP (sensitivity) | all | Memory | | | 1.10E-02 |
| WHICAP (sensitivity) | all | Verbal Ability | | | 1.46E-02 |
| WHICAP (sensitivity) | all | MMSE | | | 2.03E-01 |
| WHICAP (sensitivity) | all | CDR | | | 4.86E-03 |
| WHICAP (sensitivity) | all | Aβ42/Aβ40 | | | 1.87E-03 |

| | | | | | |
|---|---|---|---|---|---|
| WHICAP (sensitivity) | all | Total Tau | | | 4.84E-02 |
| WHICAP (sensitivity) | all | pTau | | | 1.33E-01 |
| WHICAP (sensitivity) | all | NfL | | | 1.71E-01 |
| WHICAP (sensitivity) | all | Cortical Thickness | | | 3.78E-03 |
| WHICAP (sensitivity) | all | Hippo. Volume | | | 1.09E-01 |
